# Predictions from standard epidemiological models of consequences of segregating and isolating vulnerable people into care facilities

**DOI:** 10.1101/2023.02.05.23285490

**Authors:** Joseph Hickey, Denis G. Rancourt

## Abstract

**Objectives:** Since the declaration of the COVID-19 pandemic, many governments have imposed policies to reduce contacts between people who are presumed to be particularly vulnerable to dying from respiratory illnesses and the rest of the population. These policies typically address vulnerable individuals concentrated in centralized care facilities and entail limiting social contacts with visitors, staff members, and other care home residents. We use a standard epidemiological model to investigate the impact of such circumstances on the predicted infectious disease attack rates, for interacting robust and vulnerable populations.

**Design:** We implement a general susceptible-infectious-recovered (SIR) compartmental model with two populations: robust and vulnerable. The key model parameters are the per-individual frequencies of within-group (robust-robust and vulnerable-vulnerable) and between-group (robust-vulnerable and vulnerable-robust) infectious-susceptible contacts and the recovery times of individuals in the two groups, which can be significantly longer for vulnerable people.

**Results:** Across a large range of possible model parameters including degrees of segregation versus intermingling of vulnerable and robust individuals, we find that concentrating the most vulnerable into centralized care facilities virtually always increases the infectious disease attack rate in the vulnerable group, without significant benefit to the resistant group.

**Conclusions:** Isolated care homes of vulnerable residents are predicted to be the worst possible mixing circumstances for reducing harm in epidemic or pandemic conditions.

**Strengths and limitations of this study:**

- We implement a simplest-possible sufficiently-realistic SIR model for an infectious respiratory disease with two interacting populations: robust and vulnerable.
- We investigate the predicted attack rates for a large range of parameters representing different degrees of segregation or isolation of the minority vulnerable population.
- We make broad-ranging conclusions about the consequences of segregation and isolation of vulnerable people, which apply to any epidemic model based on the SIR foundational assumptions.
- Large-parameter-range exploration is needed because the actual parameter values, especially the frequencies of infectious contacts, are not well delimited by empirical measurements and are often essentially unknown.

## Introduction

During the COVID era (from the World Health Organization (WHO) 11 March 2020 COVID-19 pandemic declaration to present), many governments have imposed policies isolating or segregating people deemed highly vulnerable to respiratory disease, including by restricting movement into and out of long-term care homes where elderly and physically or mentally disabled people reside and reducing contacts between care home residents and staff (WHO, 2020a; WHO, 2020b, pp. 5, 22; WHO, 2020c, p. 10; Low et al., 2021).

Although it was known that isolation and loneliness can have serious negative health consequences for segregated vulnerable people (Armitage & Nellums, 2020; Holt-Lunstad et al., 2015; Valtorta et al., 2016), and although it was known that residents concentrated in care homes are particularly vulnerable to infectious diseases (Strausbaugh et al., 2003; Meyer, 2004; Monto et al., 2004; Gozalo et al., 2012; Lansbury et al., 2017), and although data from the spring of 2020 showed disproportionately large all-cause mortality increases in long-term care homes that were positively correlated with the number of care home residents (Amore et al., 2021; Sundaram et al., 2021), governments continued to implement policies confining vulnerable people into care homes and reducing social contacts with visitors and staff more than one year after the WHO’s 11 March 2020 COVID-19 pandemic declaration.

Non-pharmaceutical interventions such as travel restrictions, workplace closures, and age-specific enforced social distancing or quarantining have been justified during the COVID era using theoretical infectious disease models based on the paradigm of spread by close-proximity pairwise contacts (Ferguson et al., 2020; Kreps & Kriner, 2020; Chang et al., 2020; Moss et al., 2020; Ogden et al., 2020). None of these models have been used to investigate the impact of segregation of the vulnerable into care homes.

Since policies isolating the vulnerable from contact with the majority of society have been widely applied, and since models are the main predictive tool used by governments to justify their public health policies, it is important to investigate model predictions for a large range of possible epidemiological parameters, rather than solely for limited ranges of parameters, which are not well constrained by empirical studies and which may be subject to political or institutional bias.

Large-range exploration of the parameters is needed because the actual parameter values are not well delimited by empirical measurements and are often essentially unknown; and because unexpected effects or magnitudes of effects can occur in different otherwise unexplored and relevant regions of the parameter space.

In order to appreciate the spectrum of outcomes that are possible in a given theoretical model, and its limitations and sensitivity to assumptions, it is crucial to base the model on the simplest-possible sufficiently realistic conceptual foundation and only add extensions incrementally (Garnett & Anderson, 1996; Siegenfeld et al., 2020). This approach optimizes relevance and minimizes confounding the results with complexity and intangible propagation of error. Focusing on only the core model ingredients limits the dimensionality of the model, permitting the needed examination of the model’s outcomes across a comprehensive range of parameter values. To the extent that this approach is not adopted, the model becomes more removed from reality, because each additional complexity or sophisticated model element introduces new mechanisms, and therefore new assumptions about how those mechanisms function and new uncertainties about the values of their associated parameters.

At their core, the baseline epidemiological models on which essentially all more sophisticated models are built, have two main parameters determining whether an infectious disease epidemic emerges and, if it does, its magnitude and duration. These two parameters are: the rate at which individuals experience pairwise contacts with others that could result in transmission of the infection, and the rate at which infected individuals recover and become immune.

We construct a simple susceptible-infectious-recovered (SIR) epidemic model consisting of two interacting populations, one representing the relatively robust majority of society and the other the vulnerable minority. The different health states of individuals in the two populations are represented by their different recovery times upon infection, as is well established for respiratory diseases (Faes et al., 2020; Rhee et al., 2021). We investigate the size and duration of epidemics occurring for a broad range of different within- and between-population contact frequencies representing different segregation or isolation policy-linked behaviours. This approach allows us to make broad-ranging conclusions about the consequences of segregation of vulnerable people that apply to all epidemic models based on the SIR foundational assumptions.

## Model

We implement a susceptible-infectious-recovered (SIR) model for two populations, indexed as population “*a*” and population “*b*”. The total number of *a* individuals is *N_a_* and the total number of *b* individuals is *N_b_*.

Throughout this paper, we assign the *a* population to be the majority population of robust individuals, and the *b* population to be the minority population of vulnerable individuals.

Following the usual SIR model structure, a person can be in one of three states: susceptible to infection (S), infectious (I), or recovered and immune (R). If a susceptible person comes into contact with an infectious person, the susceptible person can become infectious, and infectious people eventually recover. The numbers of susceptible, infectious, and recovered people in group *i* at time *t* are therefore *S_i_*(*t*), *I_i_*(*t*), and *R_i_*(*t*), respectively, and *N_i_* = *S_i_*(*t*) + *I_i_*(*t*) + *R_i_*(*t*).

The number of individuals in each of the three epidemiological compartments, in each of group *a* and *b*, evolve according to the following equations:

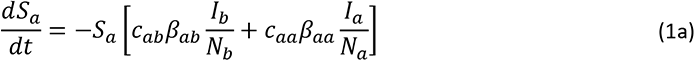

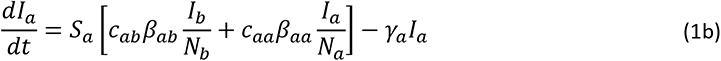

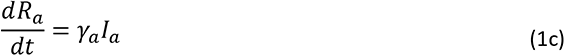

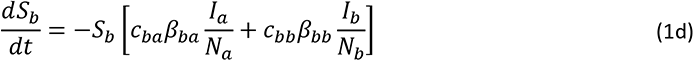

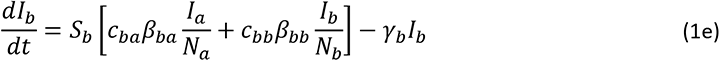

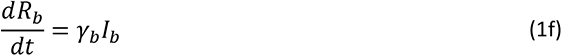

Equations 1a-f involve three sets of parameters, described below.

The parameters γ*_a_* and γ*_b_* represent the rates at which *a* and *b* individuals (robust and vulnerable individuals, respectively) recover from infection. Since the *b* population represents the minority, vulnerable population: *N_b_* ≤ *N_a_*. Since they are more vulnerable than *a* individuals, *b* individuals take a longer time to recover from infection, such that γ*_b_* ≤ γ*_a_*.

We use a value of γ*_a_* = 75 yrs^-1^ corresponding to a recovery time of approximately 5 days for healthy individuals (Wolfel et al., 2020; CDC, 2022), and we consider three values of γ*_b_*, equal to γ*_a_*, γ*_a_*/2, and γ*_a_*/4, corresponding to recovery times of approximately 5, 10, and 20 days for the *b* individuals (Faes et al., 2020; Rhee et al., 2021).

The other two sets of parameters, *c_ij_* and β*_ij_*, are intrinsically dependent, such that one set is actually redundant, which can be understood as follows. β*_ij_* represents the probability that a contact between a susceptible *i* (*a* or *b*) person and an infectious *j* person results in infection of the susceptible *i* person, whereas *c_ij_* represents the frequency (number per unit time) of contacts between an *i* person and a *j* person. Therefore, we are free to make the following simplification. Without loss of generality, in this paper we set β*_aa_* = β*_bb_* = β*_ab_* = β*_ba_* = 1. This means that the only contacts considered and counted are by definition contacts that are guaranteed to result in transmission when the contact involves a susceptible *i* person and an infectious *j* person.

There is no reason or advantage to considering other definitions of *c_ij_* having associated smaller values of β*_ij_*; and it would make no difference in the calculated results arising from Eqns. 1a-f. Under this notational and conceptual simplification, the *c_ij_* are the dominant control parameters in the model, along with the recovery rates γ*_a_* and γ*_b_*. We apply this interpretation of *c_ij_* (arising from setting all the β parameters equal to 1) throughout the remainder of the paper.

The within-group contact frequencies, *c_aa_* and *c_bb_* are independent of one another. The between-group contact frequencies *c_ab_* and *c_ba_* are also independent. However, we impose the following relationship between *c*_ab_ and *c_ba_*, modulated by the coefficient λ:

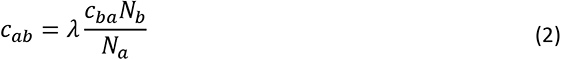

A value of λ = 1 corresponds to a strict proportionality between *c_ab_* and *c_ba_* determined purely by the relative sizes of the populations of the two groups, as would be common to impose in the sliding definition of contact in which β*_ij_* are undetermined (Garnett & Anderson, 1996).

In the present paper, λ = 1 effectively means that pairwise contact events that are of a physical proximity and duration sufficient to guarantee infection of a susceptible *b* person by an infectious *a* person are also sufficient to guarantee infection of a susceptible *a* person by an infectious *b* person. However, in principle, λ can take values less than 1, due to the more resistant health status of *a* individuals compared to *b* individuals. Since, given the relative sizes of the populations *N_a_* and *N_b_*, *c_ab_* is much smaller than *c_ba_* and typically much smaller than *c_aa_* in our analyses, we use a value of λ = 1 in the main text of this paper. In the Appendices, we show that our results are robust against smaller values of λ.

We also define *c_a_* = *c_aa_* + *c_ab_* and *c_b_* = *c_bb_* + *c_ba_* to be the total contact frequencies of *a* and *b* people, respectively. The majority, robust (*a*) population is typically younger and more socially active than the minority, vulnerable (*b*) population, such that the frequency of all person-to-person contacts is generally higher in the *a* group than the *b* group (Prem et al., 2017). However, when *c_a_* and *c_b_* represent the frequency of only those types of contacts that are guaranteed to result in infection of a susceptible individual (as per our simplifying assumption that β*_aa_* = β*_bb_* = β*_ab_* = β*_ba_* = 1, in the present article), then it is not unreasonable to consider that *c_b_* can be greater or significantly greater than *c_a_*, due to the frailer health status of the *b* individuals.

## Results

We examine the epidemic outcomes for the robust (*a*) and vulnerable (*b*) populations for a large range of possible contact frequencies and recovery rates. For specificity, we use a total population of *N* = 10^7^ individuals, with *N_a_*/*N* = *P_a_* = 0.95, such that the *a* population constitutes 95% of the entire society, and the *b* population 5%. The simulations are “seeded” with 100 infectious individuals inserted proportionally into each of the two groups, such that *I_a_*(*t*=0) = 95 and *I_b_*(*t*=0) = 5.

We verified that the results are the same on varying *P_a_*, λ, and seeding magnitude and distribution, which is shown in the Appendices.

We define the attack rate among population *i* as the proportion of initially-susceptible *i* people who become infected during the epidemic:

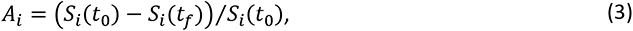

where *S_i_*(*t*_0_) is the number of susceptible *i* people at the beginning of the epidemic and *S_i_*(*t_f_*) is the number of susceptible *i* people remaining once there are no longer any infectious people in either of the two groups (*a* or *b*).

In order to examine the impact of policies that isolate or segregate the *b* individuals from the *a* group, we introduce the index *x* equal to the share of a *b* individual’s contacts that are with *a* people:

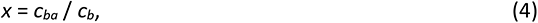

When *x* = 0, *b* individuals only ever have contacts with other *b* individuals, and when *x* = 1, *b* individuals only ever have contacts with *a* people. In this way, *x*, represents the degree of segregation versus intermingling of the *a* and *b* groups. Complete segregation is *x* = 0. Complete *a-b* intermingling, while avoiding all *b-b* contacts, is *x* = 1.

Fig. 1 shows the evolution of the epidemic (number of new cases per day, over time) in the *a* and *b* groups, for different values of *x*. In this example, *c_a_* is slightly larger than γ*_a_* (in order that *c_a_* / γ*_a_* (*“R_0_*”) ≈ 1.1 > 1 such that an epidemic would occur in the *a* group if it were completely isolated from the *b* group) and *c_b_* is 25% larger than *c_a_*. γ*_b_* = γ*_a_*/4, such that *b* people take four times as long to recover from infection as *a* people.

**Figure 1:**
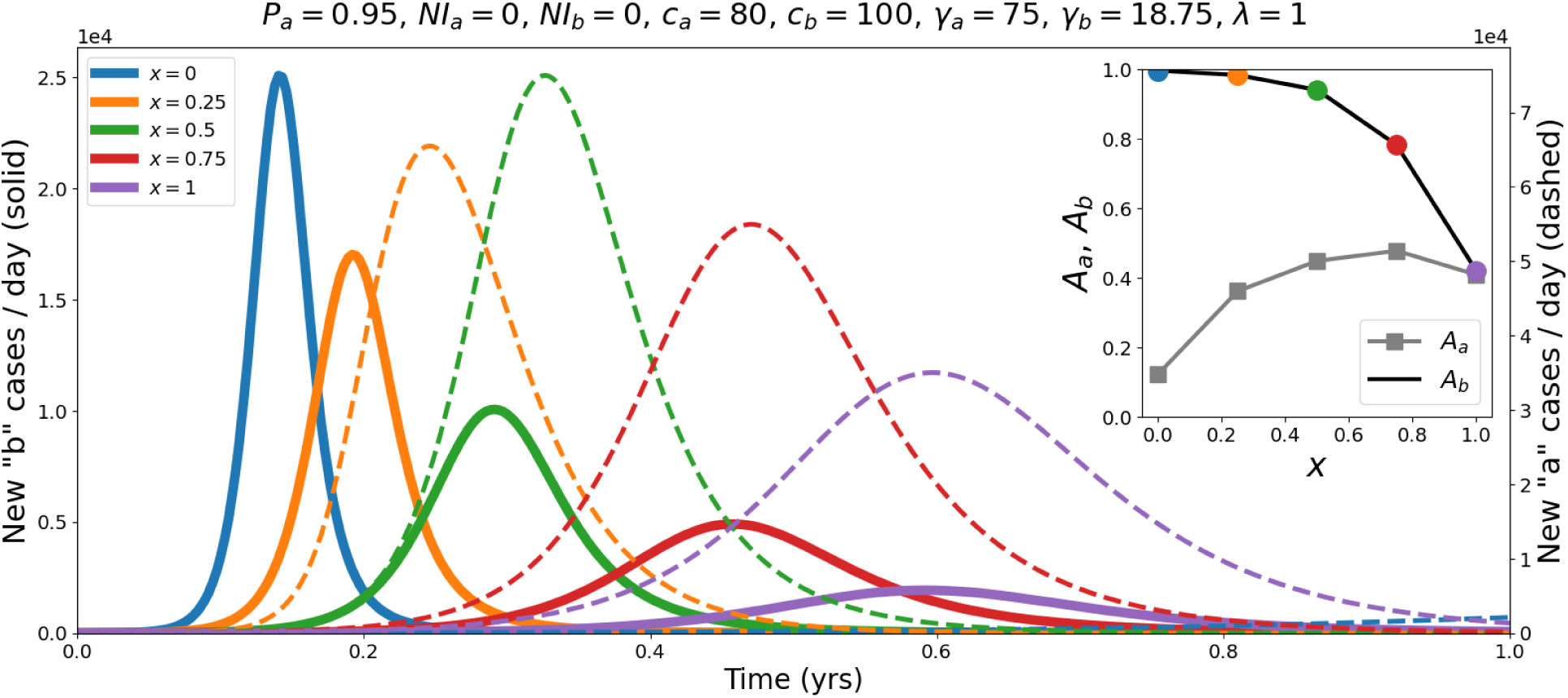
Epidemic curves showing the number of new cases per day in population *b* (vulnerable, minority group, solid lines, left y-axis) population *a* (robust, majority group, dashed lines, right y-axis), for different values of *x*, and for the fixed model parameters indicated above the figure. Inset: attack-rates *A_a_* and *A_b_* as functions of *x* (coloured circles indicate the *x* values listed in the main figure legend).

As can be seen in Fig. 1, *x* (the degree of separation or intermingling) has a large effect on the size and duration of the epidemics occurring in both the *a* (robust, majority) and *b* (vulnerable, minority) groups.

When *x* = 0, *b* individuals only ever come into contact with other *b*’s, and the number of new cases per day in the *b* group rapidly surges, peaks, and decays, and essentially all of the *b* population becomes infected (*A_b_* ≈ 1, inset of Fig. 1). An epidemic also occurs in the *a* group, but the attack rate is smaller (*A_a_*, inset) and it takes significantly longer for the epidemic to transpire (see the dashed blue line in the extreme lower-right corner of Fig. 1).

In Fig. 1, as *x* is increased above 0, a larger and larger share of *b* contacts are with *a* individuals. In the *b* group, the epidemic size (peak value of new cases per day and attack-rate) decreases with increasing *x* and the duration of the epidemic increases. Going from *x* = 0.5 to *x* = 0.75 and *x* = 1, *A_b_* is significantly decreased, to the point where less than half of the susceptible, vulnerable *b* population becomes infected. On the other hand, increasing *x* above 0 initially increases *A_a_* and significantly shortens the time it takes for the number of new *a* cases per day to surge and decay. When *x* = 1, the epidemic curves for the *a* and *b* populations have their peaks at approximately the same time, and the attack rates become similar for the two groups.

Fig. 1 illustrates the important effect of *x* on the epidemic outcomes in the two populations. In particular, it is apparent that larger *x* (more contacts with robust individuals) can produce significantly better (lower attack rate) results for the minority vulnerable population. This is important if it is a general feature because the vulnerable individuals in the real world have higher risk of dying on being infected (COVID-19 Forecasting Team, 2022), which is the motivation for wanting to protect them.

Next, we present figures showing results across our large range of possible and reasonable *c_a_* and *c_b_* values, for different degrees of segregation vs. intermingling, *x*, between the *a* and *b* groups, and for the different values of γ*_b_* representing different degrees of vulnerability of the *b* population.

Fig. 2 contains a collection of panels showing how the attack-rates *A_a_* and *A_b_* change as *c_a_* and *c_b_* are varied. Each panel corresponds to a choice of *x* _and_ γ*_b_*.

**Figure 2:**
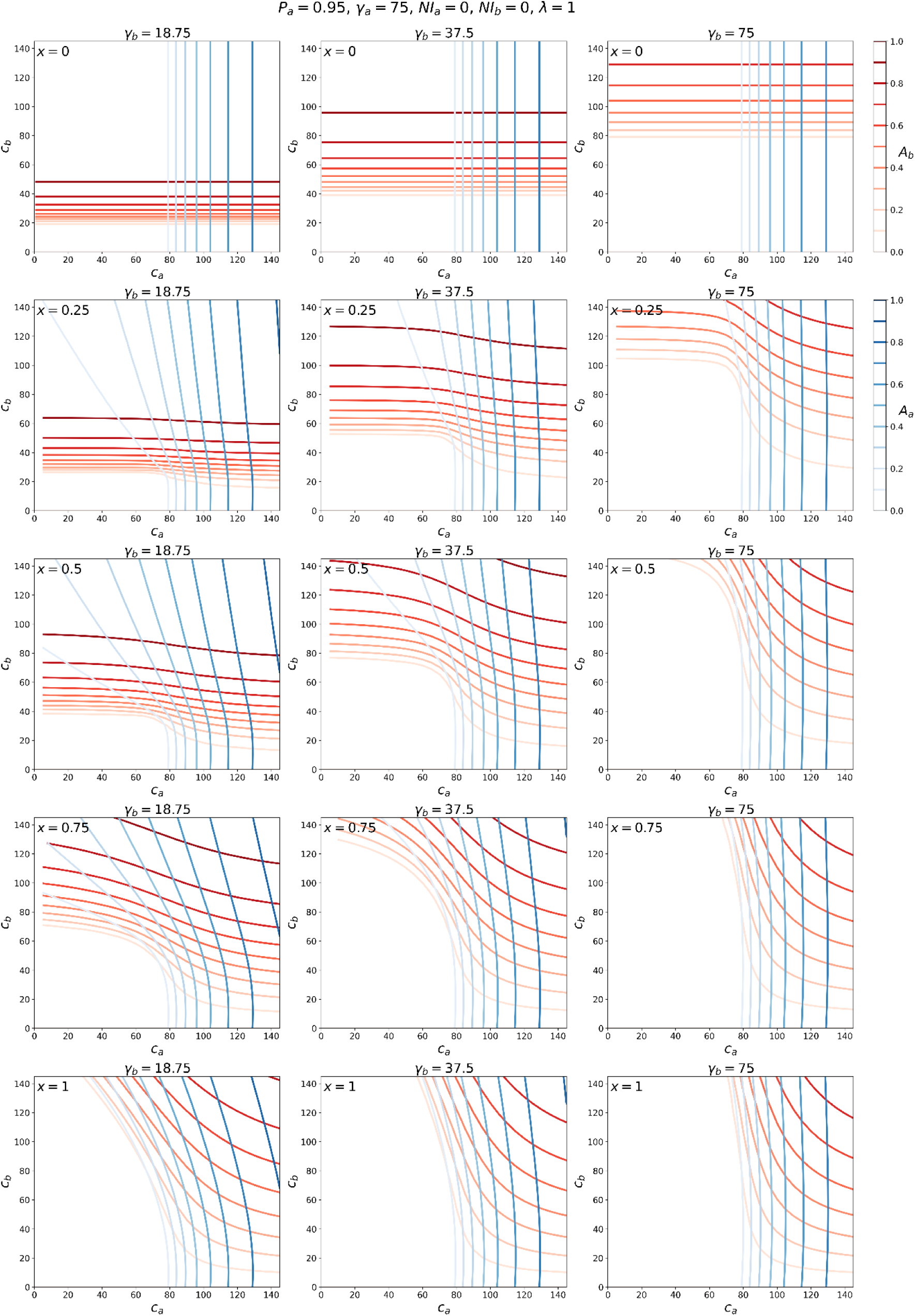
Contour maps of *A_a_* (blue lines, see scale at the upper right) and *A_b_* (red lines, see scale at the upper right) for a range of contact frequencies *c_a_* and *c_b_*. Each column of panels corresponds to a different γ*_b_* and each row to a different *x*, as indicated.

The panel in the upper-left corner of Fig. 2 corresponds to *x* = 0 and γ*_b_* = γ*_a_*/4 = 18.75. Since *x* = 0, there is complete segregation between the *a* and *b* groups. In this case, an epidemic emerges in the *a* group when *c_a_* > γ*_a_* and in the *b* group when *c_b_* > γ*_b_*, and this can be seen by the fact that *A_a_* > 0 when *c_a_* > 75, for all values of *c_b_*, and *A_b_* > 0 when *c_b_* > 18.75, for all values of *c_a_*. Thus, when *x* = 0, we see the usual transition to an epidemic, which occurs in a one-population SIR model when *R* = *c*/γ > 1, in each group.

The panels in the second through fifth rows of Fig. 2 correspond to *x* > 0, progressively increasing up to *x* = 1 (fifth row). For many values of *c_a_*, increasing *x* results in a shift upwards (to higher *c_b_* values) of the red contour lines, indicating a decrease in *A_b_* for fixed *c_b_*.

For example, when γ*_b_* = 18.75 (left column of panels), *c_a_* = 20 and *c_b_* = 40, the attack rate *A_b_* is large when *x* = 0. However, as *x* is increased, the red contour lines shift upward, indicating a lowering of the attack rate at (*c_a_*, *c_b_*) = (20, 40), until *A_b_* = 0 (no epidemic in the *b* population) in the second-last and last panels in the column (*x* = 0.75 and *x* = 1).

The positioning of the blue contour lines (*A_a_*) is generally less affected by changes in *x* than that of the red contours. This is particularly evident for the case of γ*_b_* = γ*_a_* (right column of panels). This is due to the asymmetry in the sizes of the populations of the *a* and *b* groups (*N_b_* being 5% of the total population).

To better appreciate the model results summarized in the contour maps of Fig. 2, it is helpful to simultaneously examine the attack rates for a particular point in the (*c_a_*, *c_b_*) parameter-space as *x* is varied. This is shown in Figs. 3-5.

**Figure 3:**
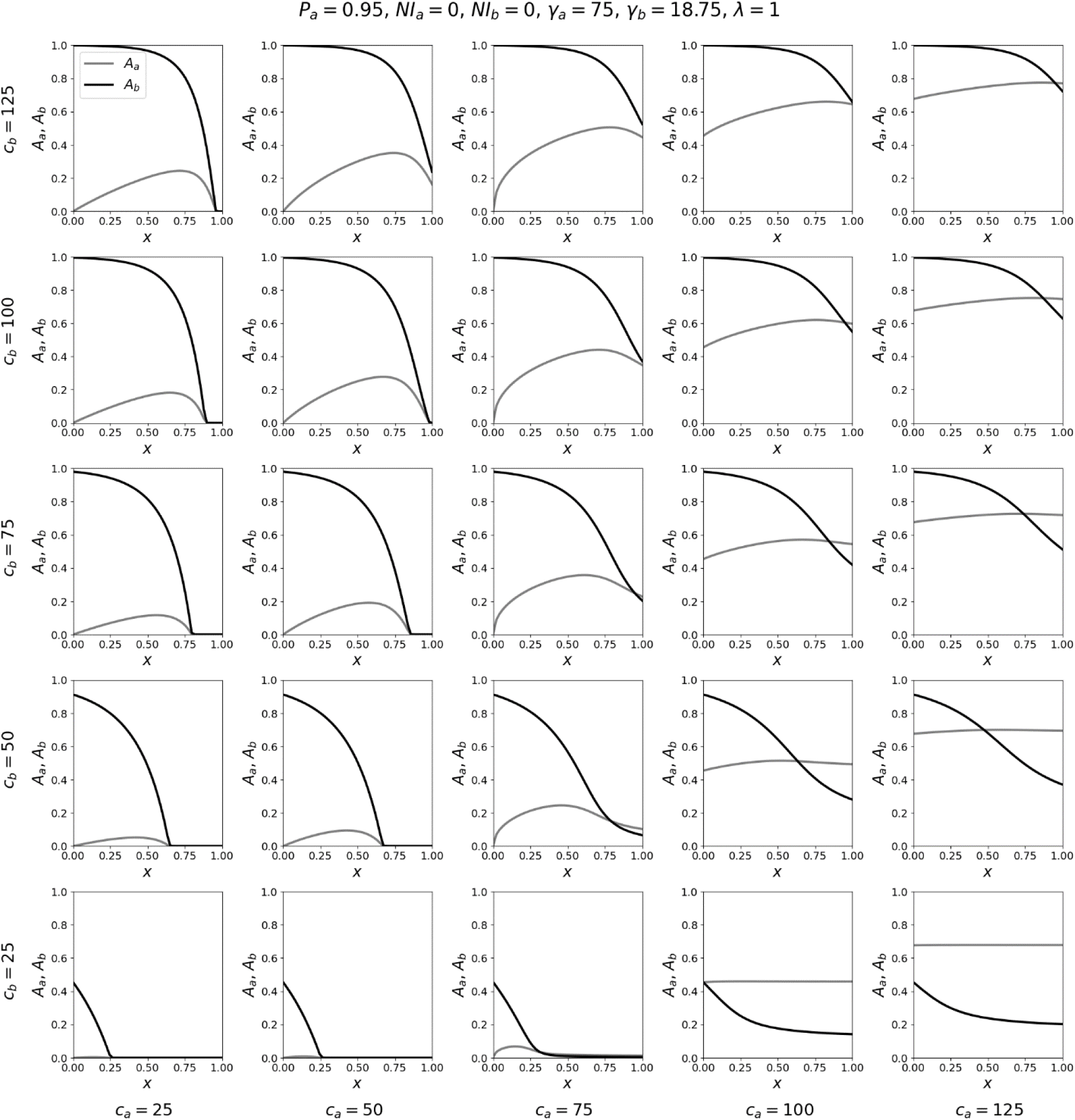
Attack-rates *A_a_* and *A_b_* as functions of *x*, for a range of contact frequencies *c_a_* and *c_b_*, for γ*_b_* = γ*_a_*/4.

Figs. 3-5 show the variation in the attack rates *A_a_* and *A_b_* as functions of *x*, for various (*c_a_*, *c_b_*) coordinates. Each panel is for one pair of the (*c_a_*, *c_b_*) coordinates, with *c_a_* increasing (in columns) from left to right, and *c_b_* decreasing (in rows) from top to bottom. In this way, one can visualize the behaviours of the attack rates with *x*, on the (*c_a_*, *c_b_*) plane, across a range of *c_a_* and *c_b_* values sampled from the phase diagrams shown in Fig. 2.

As can be seen, when γ*_b_* = γ*_a_*/4 (Fig. 3), increasing *x* decreases *A_b_* for all values of (*c_a_*, *c_b_*) shown in the figure. The decrease in *A_b_* can be dramatic, including going from *A_b_* = 1 for small values of *x* to *A_b_* = 0 for large values of *x*. Increasing *x* generally increases *A_a_*, and the increase in *A_a_* is largest for values of *c_a_* ≤ γ*_a_* (such that no epidemic would occur in the *a* group if it were completely isolated from the *b* group) and for intermediate values of *x*. When *c_a_* > γ*_a_*, increasing *x* has a very small effect on *A_a_*, because the *a* group has a much larger population than the *b* group; this is reflected in the small changes in the blue contour lines in Fig. 2 for *c_a_* > γ*_a_* and for increasing *x*.

When γ*_b_* = γ*_a_*/2 (Fig. 4), increasing *x* generally decreases *A_b_*, similar to the results in Fig. 3, and *x* has a smaller effect on *A_a_* compared to the results in Fig. 3. The only parameter values for which *A_b_* increases with *x* are in the extreme lower-right corner of Fig. 4, for which (*c_a_*, *c_b_*) = (100, 25) and (125, 25). For these two pairs of (*c_a_*, *c_b_*) values, *c_b_* < γ*_b_*, such that the contact frequency of *b* individuals is so low that an epidemic would not occur among the vulnerable if they were completely excluded from the majority group. Furthermore, for (*c_a_*, *c_b_*) = (100, 25) and (125, 25), *c_a_* is much greater than *c_b_*, which is unrealistic given our interpretation of *c_ij_* implied by our simplifying assumption β*_aa_* = β*_bb_* = β*_ab_* = β*_ba_* = 1 (see the Model section). We note that a similar, small increase in *A_b_* versus *x* also occurs in the case of γ*_b_* = 18.75 when *c_b_* < γ*_b_* and *c_a_* >> *c_b_*, as can be seen in the left column of panels in Fig. 2, e.g. when *c_b_* ≈ 15 and *c_a_* = 120.

**Figure 4:**
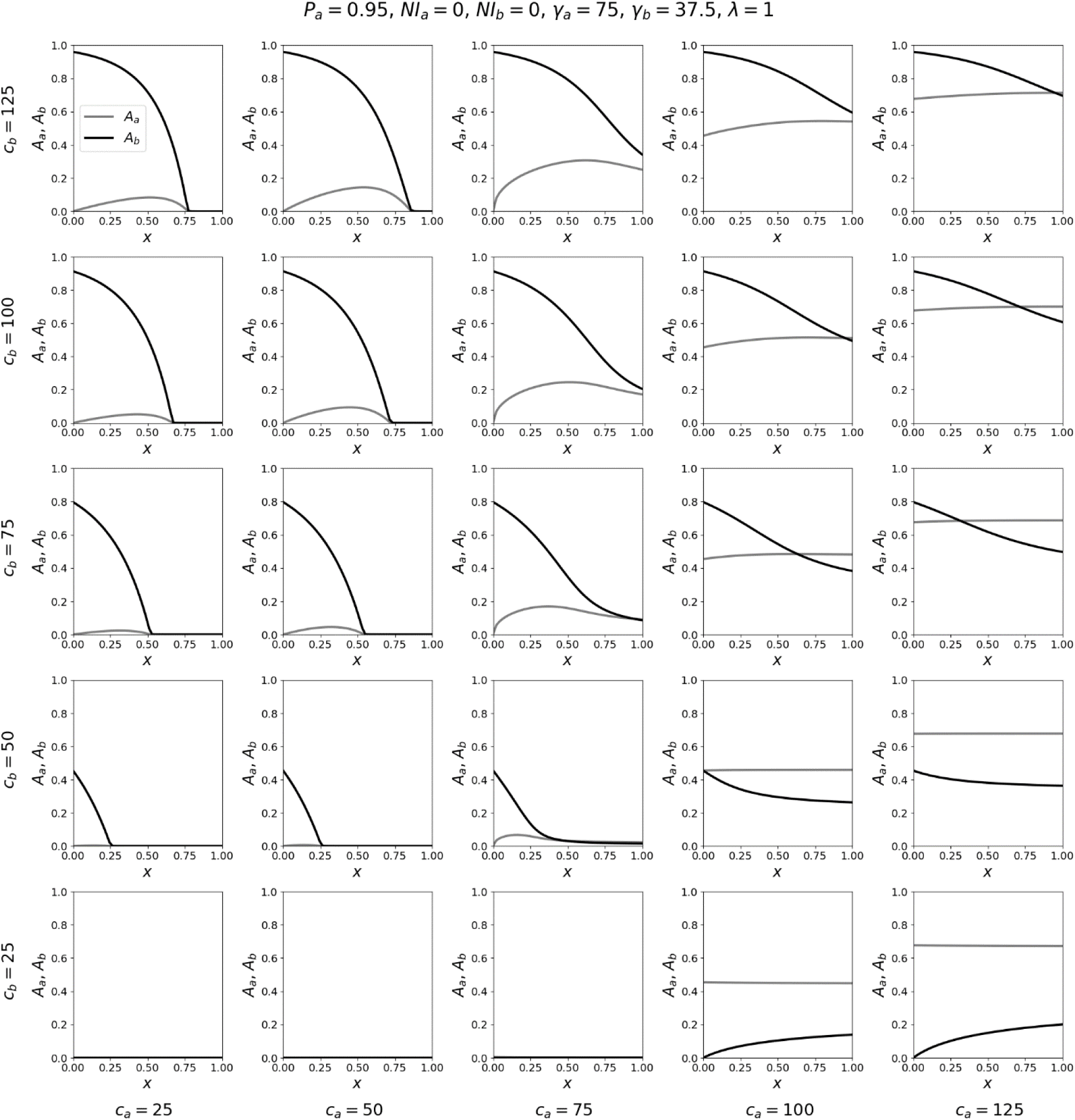
Same as Fig. 3, with γ*_b_* = γ*_a_*/2.

When γ*_b_* = γ*_a_* (Fig. 5), *x* has little effect on *A_a_*, due to the differences in population sizes of the *a* and *b* groups. Increasing *x* can decrease *A_b_* significantly when *c_b_* >> *c_a_* (panels in the upper-left corner of Fig. 5) and can increase *A_b_* significantly when *c_a_* >> *c_b_* (panels in the lower-right corner of Fig. 5). This asymmetry occurs because of the asymmetry in population sizes *N_a_* and *N_b_*, causing *c_ab_* << *c_ba_* (when λ = 1) such that it is much less likely for any given *a* person to come into contact with a *b* person than vice-versa. Similarly, in the right column of panels in Fig. 2, increasing *x* has a large effect on the red (*A_b_*) contour lines and essentially no effect on the blue (*A_a_*) contour lines.

**Figure 5:**
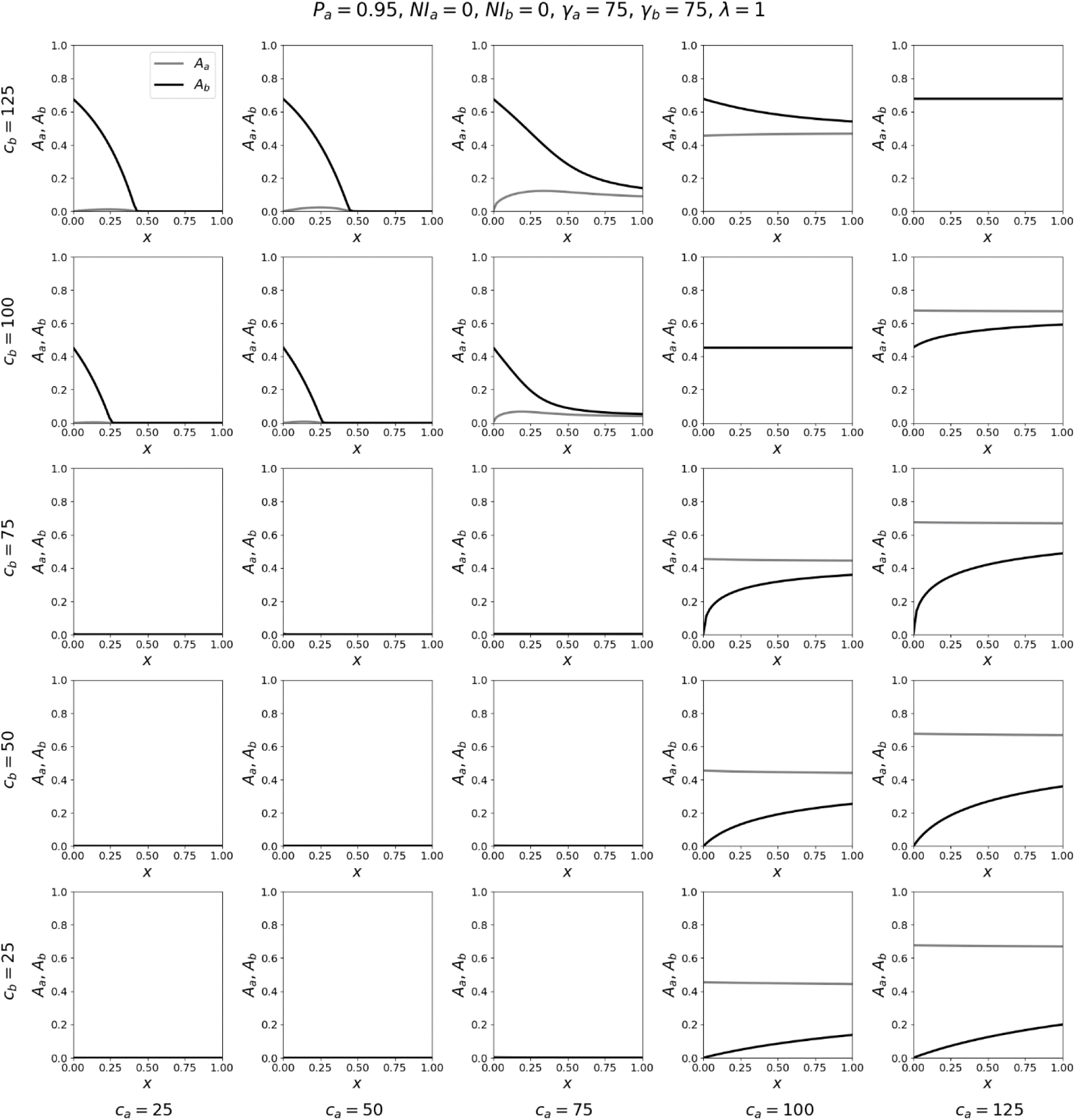
Same as Fig. 3, with γ*_b_* = γ*_a_*.

In summary, increasing *x* for fixed *c_b_* and *c_a_* decreases the attack rate in the vulnerable group across all realistic values of the contact frequencies, when *b* represents a minority vulnerable population (here making up 5% of the total population and having a recovery time twice or four times as long as for the robust majority). This means that the vulnerable population is harmed by isolation from the robust population and benefits from mixing with or dilution within the robust population, in terms of risk of infection during the course of the epidemic or pandemic.

In the Appendices, we show that the same results hold when varying *P_a_*, λ, and the seeding magnitude and distribution.

## Discussion

Using a general two-population epidemic model, we have shown that increasing the degree of intermingling of the minority vulnerable (*b*) population with the majority robust (*a*) population reduces the attack-rate among the vulnerable. The advantage to the vulnerable group of intermingling with the robust group increases as the vulnerability of the minority group increases, that is, as their disease recovery time increases. Increasing the share of a vulnerable person’s interactions that are with other vulnerable people, by confining them together in the same facility, increases the likelihood of infection of the vulnerable person during the course of the epidemic or pandemic, because infected vulnerable people remain infectious for a long time, relative to robust people.

The only exception to this general rule occurs if the contact frequency for vulnerable individuals is so small that no epidemic would occur in the vulnerable group if it were completely segregated from the robust majority of society, while the frequency of guaranteed infection-causing contacts for robust people is large enough to produce an epidemic in that group and is also much higher than that of vulnerable individuals. We expect this exception to be irrelevant in reality because it is unrealistic for *c_a_* >> *c_b_*, given the definition of the contact frequencies *c_ij_* as representing contacts of sufficient physical proximity and duration such that a susceptible *i* person is guaranteed to be infected by an infectious *j* person (see the Model section).

Our analysis focuses on the two dominant and most fundamental features present in all epidemic models: the contact frequencies and recovery rates. On this simplest-possible yet sufficiently realistic foundation, we establish that segregating the vulnerable into care homes virtually always produces negative results in epidemic models. Not surprisingly, therefore, researchers using complex agent-based models have found that segregation of vulnerable individuals produces worse outcomes both for that group and for the society overall (Markovič et al., 2021).

Others have used simple epidemiological models to study segregation of “high-transmission-risk” and “low-transmission-risk” groups (Munday et al., 2018; Yuan et al., 2022; Garnett & Anderson, 1996). However, because such studies are focused on different transmission rates due to different behavioural and contact characteristics of the two groups – such as sexual preferences, cultural lifestyle factors, and willingness to become vaccinated – they do not consider the impact of different recovery rates for the two populations, which is crucial in the context of segregation of vulnerable individuals from the robust majority. Those studies, therefore, do not directly address the problem of society’s vulnerable sector regarding infectious diseases.

Segregation based on vaccination status has also been studied recently using simple models (Hickey & Rancourt, 2022; Fisman et al., 2022; Virk, 2022; Kosinski, 2021). In this application, Hickey and Rancourt found that the effect of the segregation on increasing or decreasing the contact frequencies in the segregated groups is crucial and can cause the predicted epidemic outcomes to be worse for both the vaccinated and unvaccinated, compared to no segregation (Hickey & Rancourt, 2022). This highlights the importance of contact frequencies, which are necessarily impacted by segregation policies, and which again play a pivotal role in the present analysis.

Isolation policies intending to protect the vulnerable reduce their contacts with the outside world, for example by barring visitors from entering care homes and by reducing the frequency of interaction between care home staff and residents. The care home isolation policies are also designed to reduce the number of epidemiological contacts between the care home residents themselves. However, since transmission of respiratory diseases is air-borne via long-lived suspended aerosol particles (Shaman & Kohn, 2009; Shaman et al., 2010) and occurs in indoor environments (Bulfone et al., 2021), confining many vulnerable people in the same facility in-effect increases the per-individual frequency of infectious contacts, because they are breathing the same air and ventilation is imperfect. Indeed, virtually all studied outbreaks of viral respiratory illnesses have occurred in indoor environments (Moser et al., 1979; Loeb et al., 2000; Salgado et al., 2002; Bulfone et al., 2021; Javid et al., 2021) and care homes for the elderly are known to be “ideal environments” for outbreaks of infectious respiratory diseases, due to the susceptibility of the residents living in close quarters (Strausbaugh et al., 2003; Gozalo et al., 2012; Lansbury et al., 2017). A policy that decrease *c_ba_*, for example by barring younger family members from entering care homes to visit their elderly relatives, causes the isolated vulnerable people to spend more time in the care home, breathing the same air as the other residents. This in-effect increases *c_bb_*.

For constant *c_a_*, decreasing *c_b_* reduces the attack rate in the vulnerable group, regardless of the value of *x*, as can be seen from Fig. 2. However, the sought decreasing of *c_b_* is imposed by isolating the vulnerable (from society, loved ones and each other), which has important negative health consequences (Cohen et al., 1991; Cohen et al., 1997; Cohen, 2004; Holt-Lunstad et al., 2010; Holt-Lunstad et al., 2015; Valtorta et al., 2016). Psychosocial factors, including depression, lack of social support, and loneliness are known to play key roles in the negative health effects of isolation (Hemingway & Marmot, 1999; K.A. Matthews et al., 2010; Elovainio et al., 2017; Groarke et al., 2020; Spring et al., 2020). Proposed psychosocial factors uncovered by participatory qualitative research include dissonance between expectations and reality (Wang et al., 2020; Tarlov, 1996), which could be significant for vulnerable elderly patients with no prior life experience relevant to the isolation measures applied during the COVID era, which had no historical precedent.

Whereas governments used theoretical epidemic models to justify most public health policies during the COVID era, within a tunnel vision of reducing risk of infection with a particular virus, they appear not to have considered what those same models predict about infection rates under conditions of care home segregation; and they appear to have disregarded the exponential increase of infection fatality rate with age (COVID-19 Forecasting Team, 2022). Care home segregation policies may have been responsible for many deaths attributed to COVID-19 in Western countries.

We conclude that segregation and isolation of the vulnerable into care homes as a strategy to reduce the risk of infection during the course of an epidemic or pandemic is contrary to the most relevant immediate considerations from epidemiological models, in realistic conditions in which vulnerable people are highly susceptible and take longer to recover. The model parameter space, within possible parameter values, is one where it is virtually never epidemiologically advantageous to segregate and isolate frail people.

## Data Availability

Theoretical, mathematical modeling study. The mathematical model is described in the manuscript text.

## Appendices

### Table of Symbols

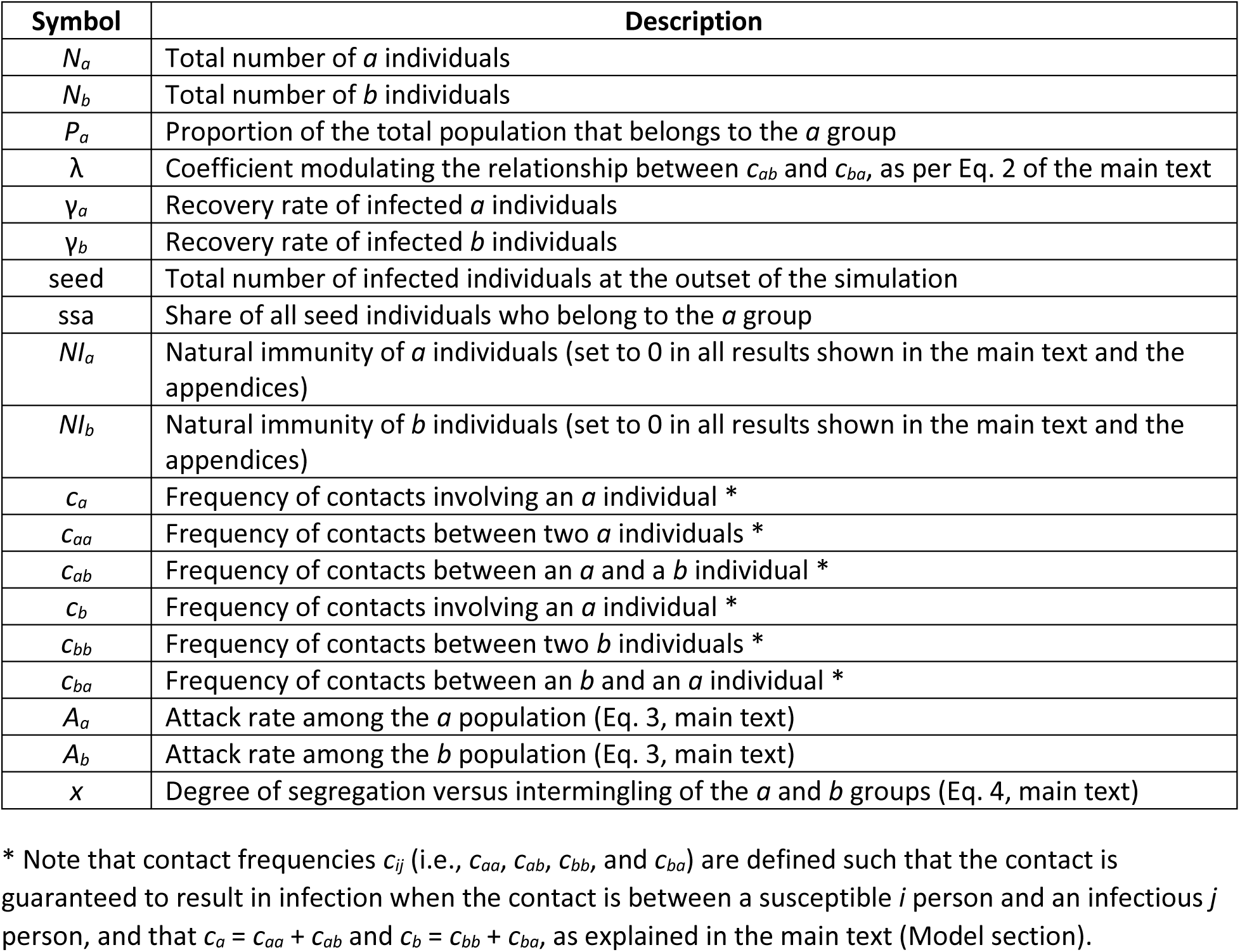

| Symbol | Description |
| --- | --- |
| $N_a$ | Total number of $a$ individuals |
| $N_b$ | Total number of $b$ individuals |
| $P_a$ | Proportion of the total population that belongs to the $a$ group |
| $\lambda$ | Coefficient modulating the relationship between $c_{ab}$ and $c_{ba}$, as per Eq. 2 of the main text |
| $\gamma_a$ | Recovery rate of infected $a$ individuals |
| $\gamma_b$ | Recovery rate of infected $b$ individuals |
| seed | Total number of infected individuals at the outset of the simulation |
| ssa | Share of all seed individuals who belong to the $a$ group |
| $NI_a$ | Natural immunity of $a$ individuals (set to 0 in all results shown in the main text and the appendices) |
| $NI_b$ | Natural immunity of $b$ individuals (set to 0 in all results shown in the main text and the appendices) |
| $c_a$ | Frequency of contacts involving an $a$ individual * |
| $c_{aa}$ | Frequency of contacts between two $a$ individuals * |
| $c_{ab}$ | Frequency of contacts between an $a$ and a $b$ individual * |
| $c_b$ | Frequency of contacts involving an $a$ individual * |
| $c_{bb}$ | Frequency of contacts between two $b$ individuals * |
| $c_{ba}$ | Frequency of contacts between an $b$ and an $a$ individual * |
| $A_a$ | Attack rate among the $a$ population (Eq. 3, main text) |
| $A_b$ | Attack rate among the $b$ population (Eq. 3, main text) |
| $x$ | Degree of segregation versus intermingling of the $a$ and $b$ groups (Eq. 4, main text) |

\* Note that contact frequencies $c_{ij}$ (i.e., $c_{aa}$ , $c_{ab}$ , $c_{bb}$ , and $c_{ba}$ ) are defined such that the contact is guaranteed to result in infection when the contact is between a susceptible $i$ person and an infectious $j$ person, and that $c_a = c_{aa} + c_{ab}$ and $c_b = c_{bb} + c_{ba}$ , as explained in the main text (Model section).

### Appendix A: Additional results for *P_a_* = 0.95

#### A.1: Attack-rate contour maps for different values of λ

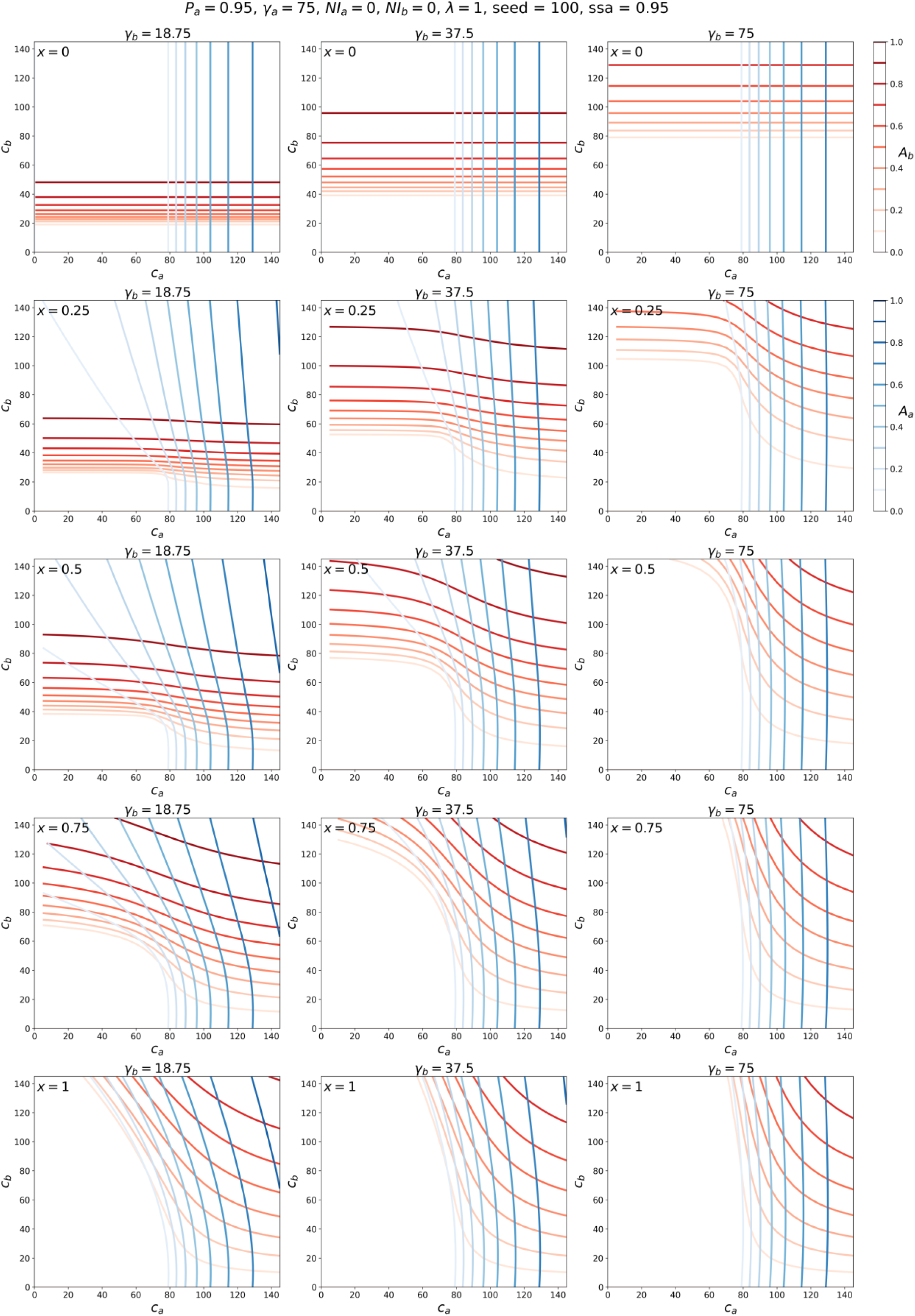
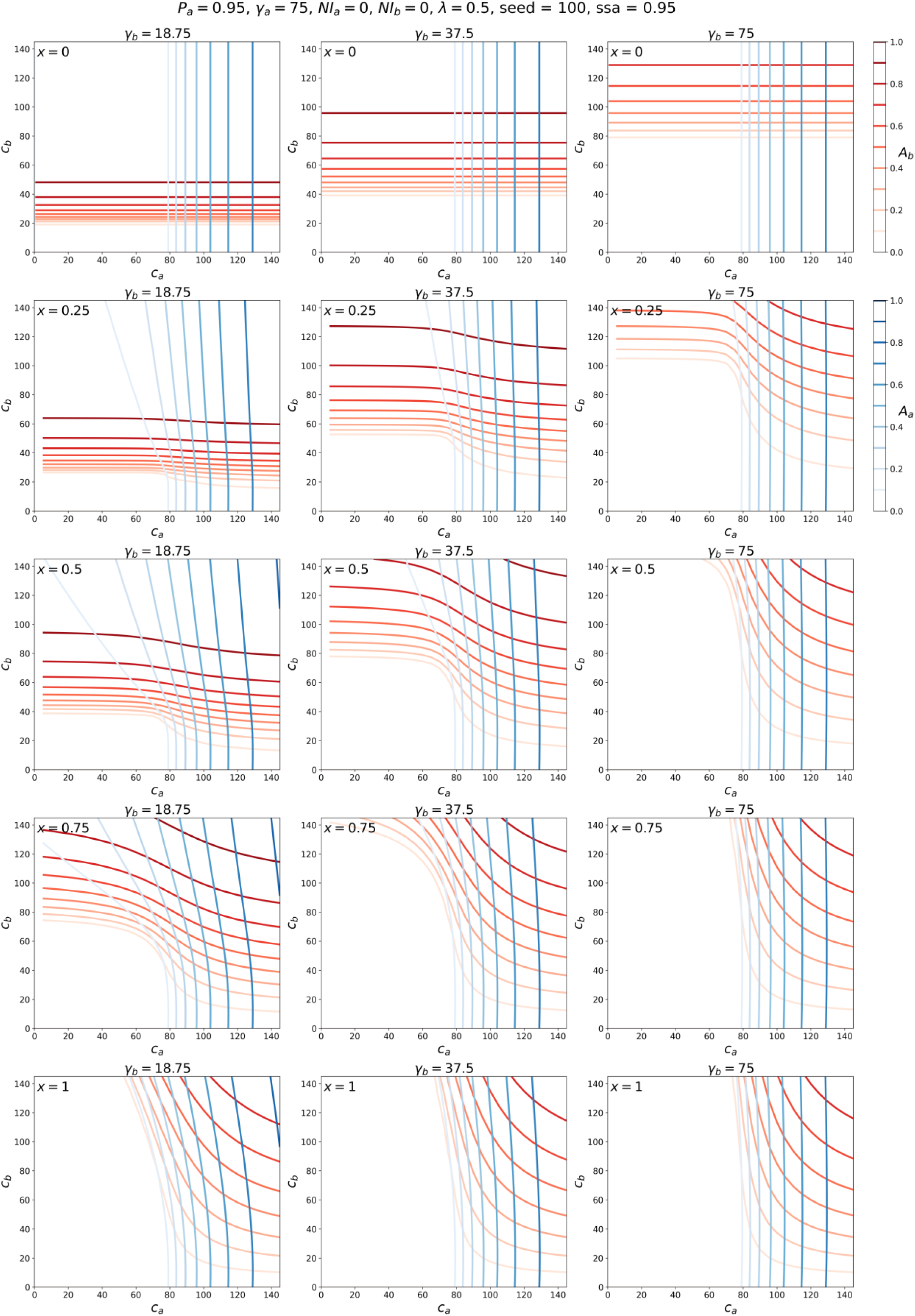
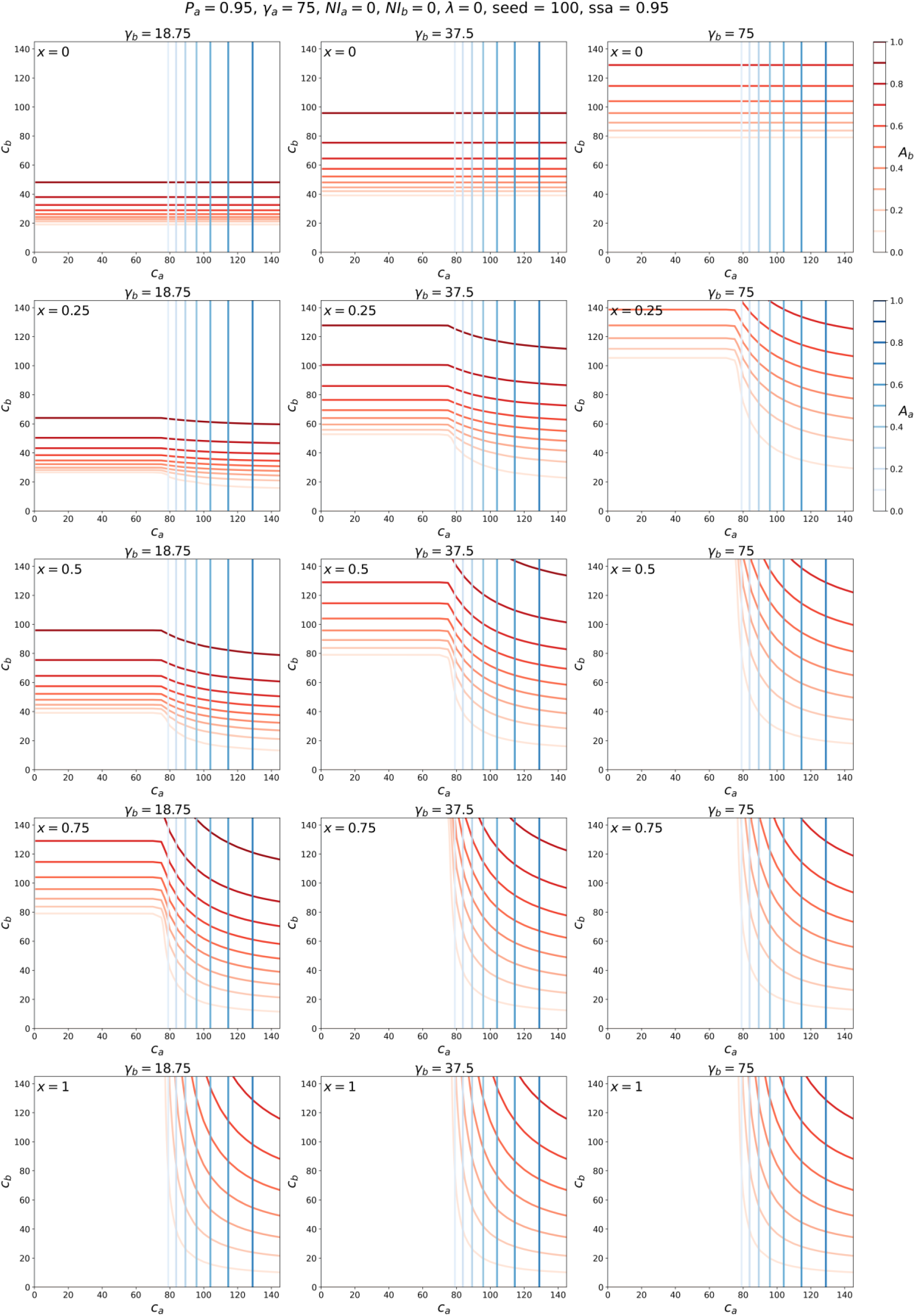

#### A.2: Attack-rate vs. x composite plots, for different values of λ

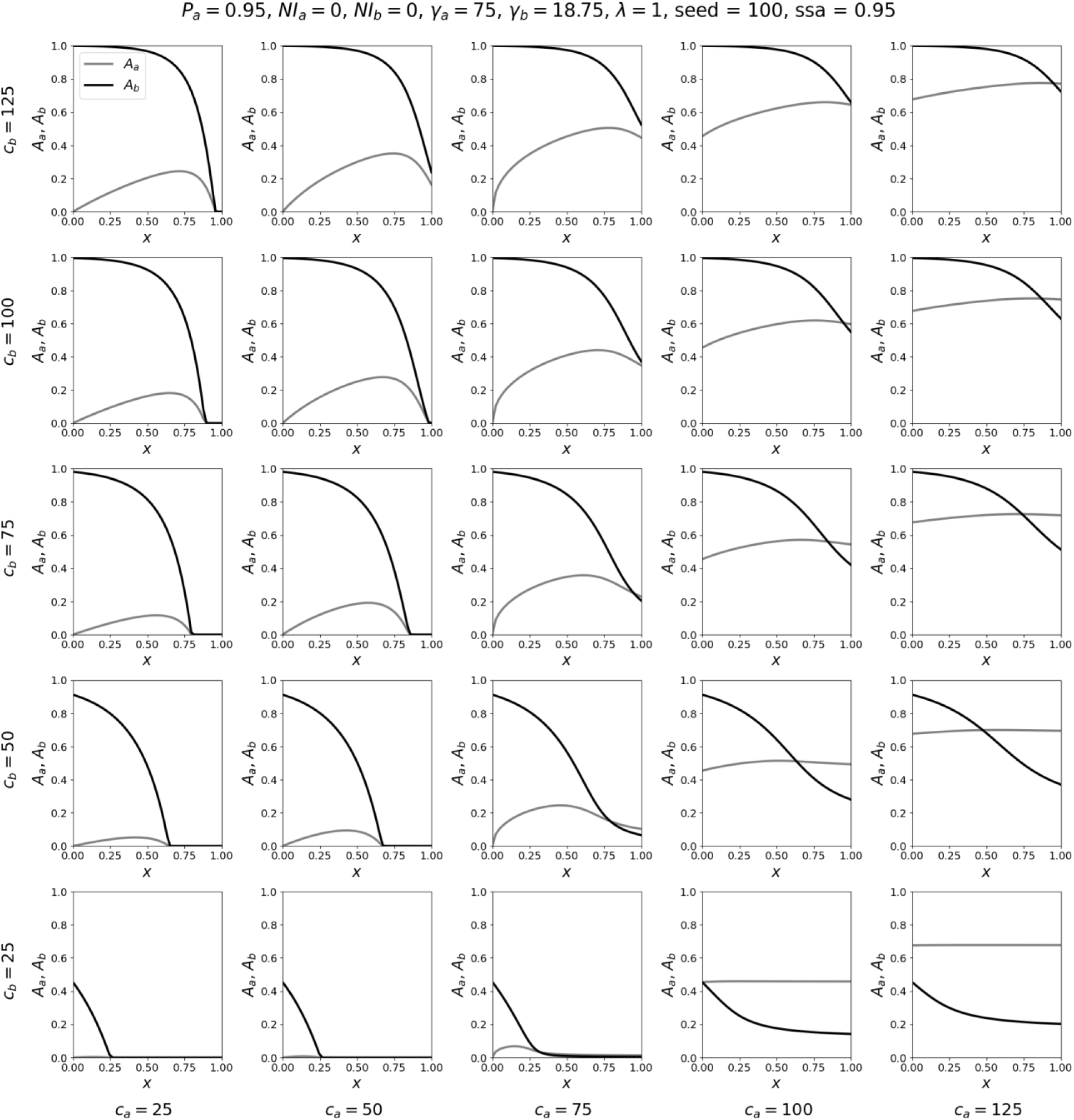
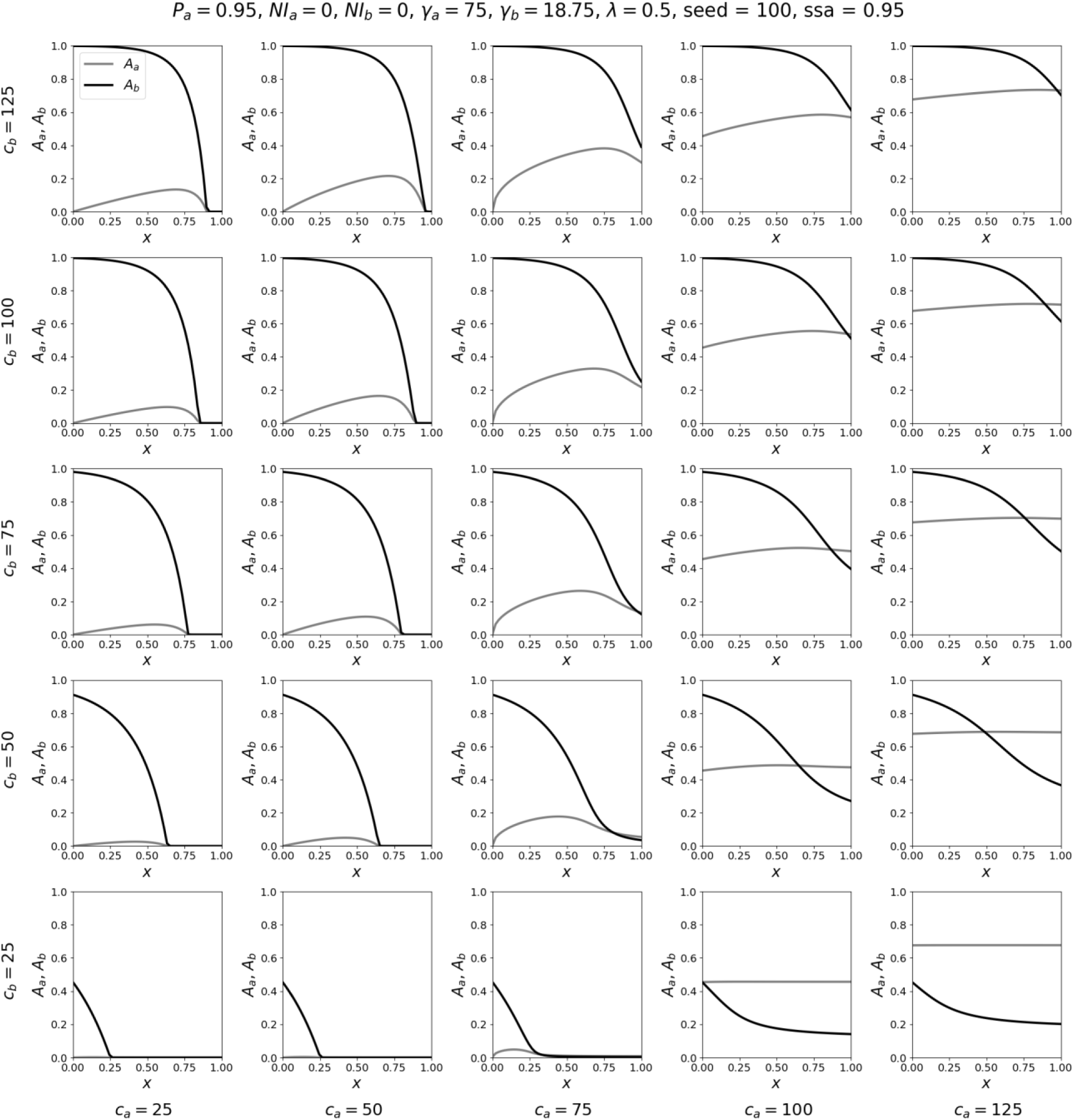
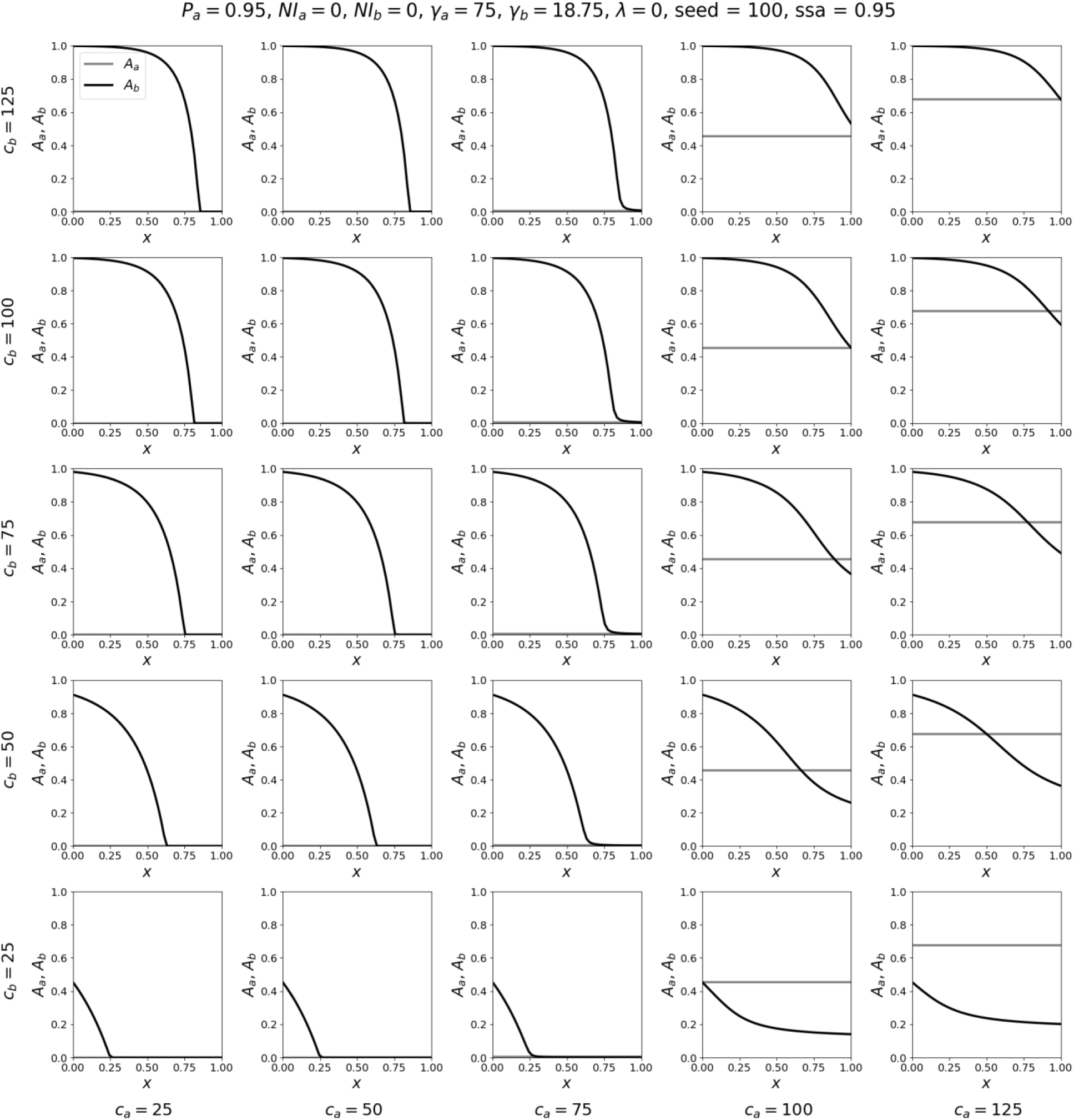
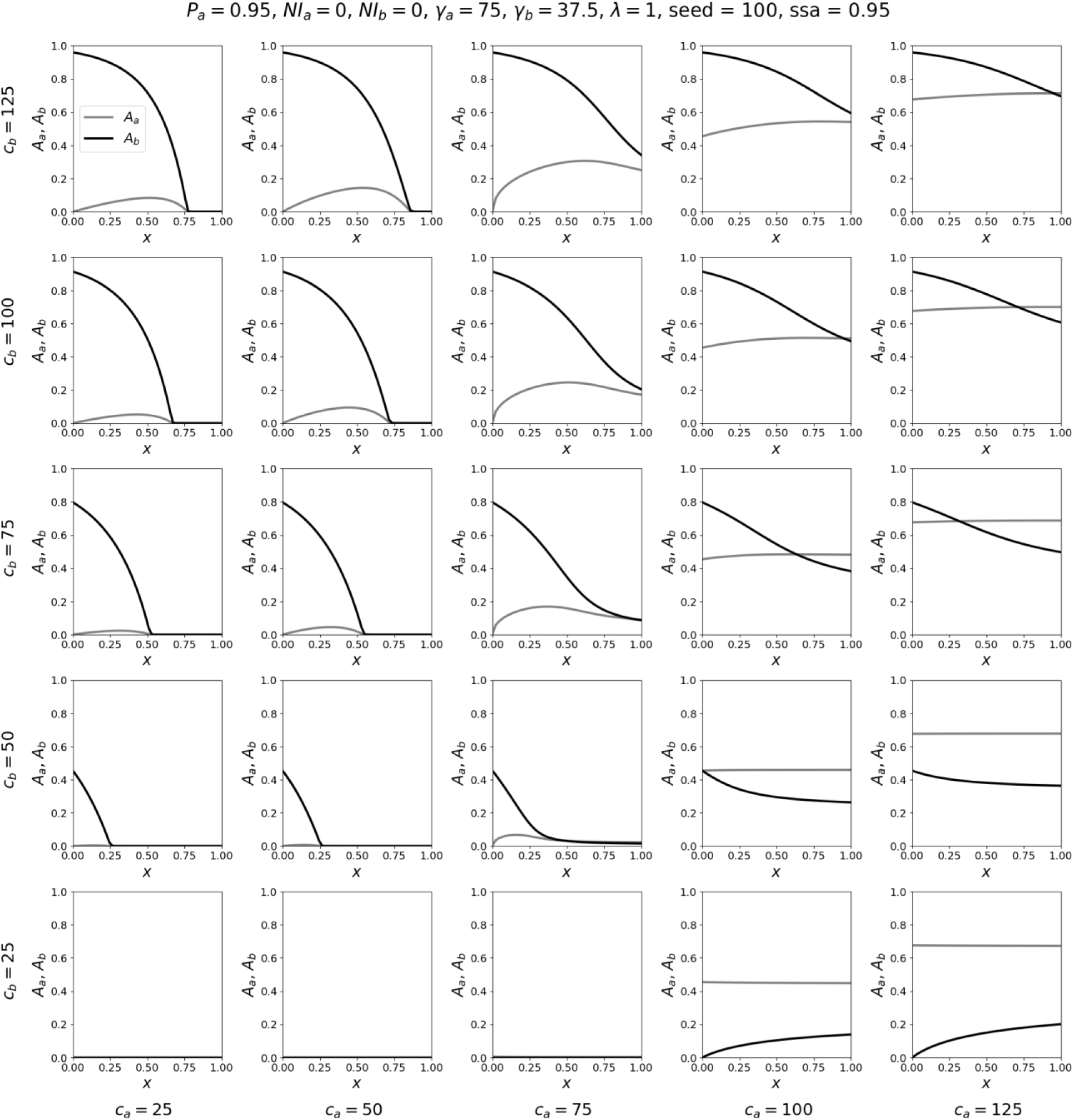
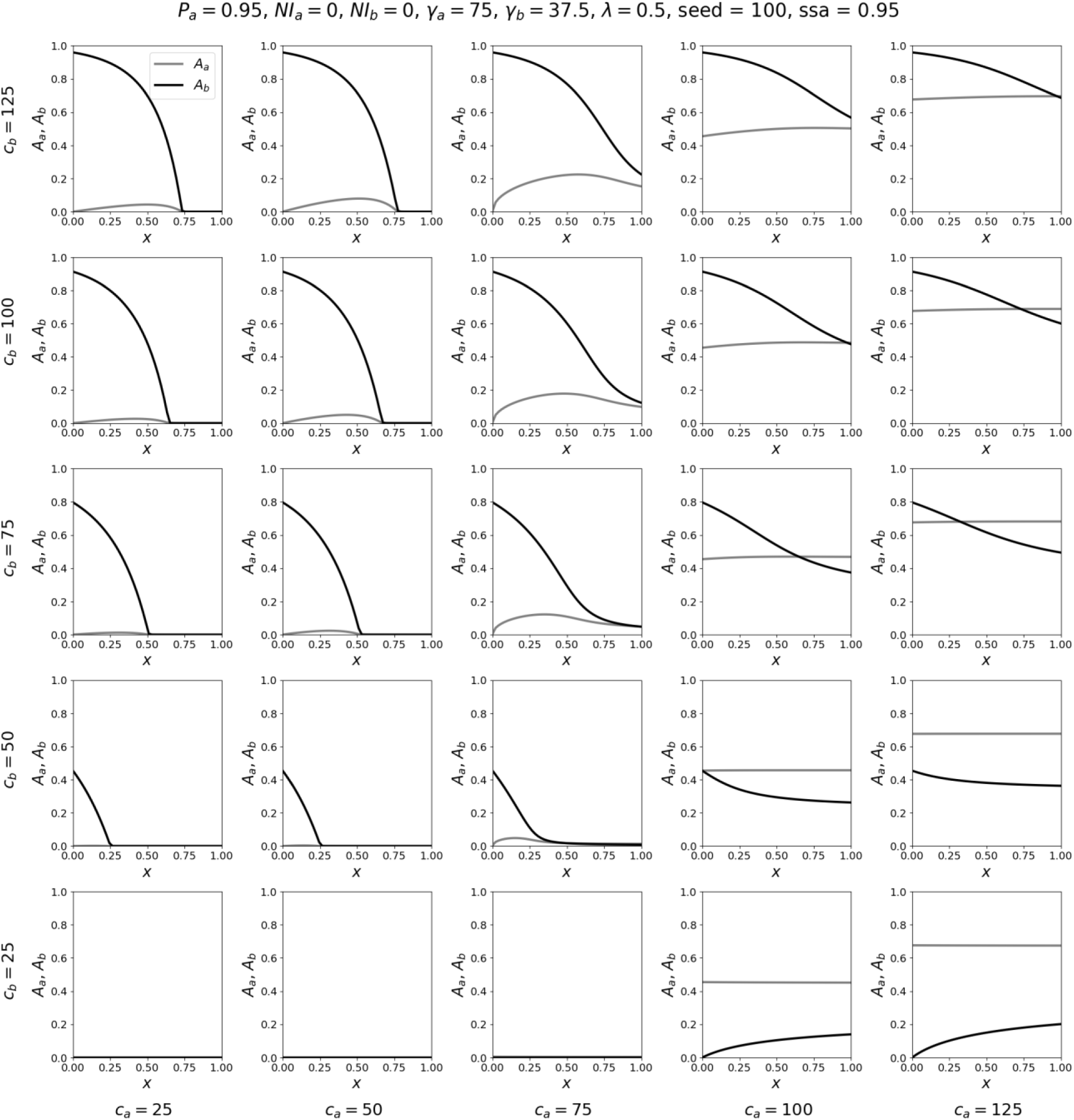
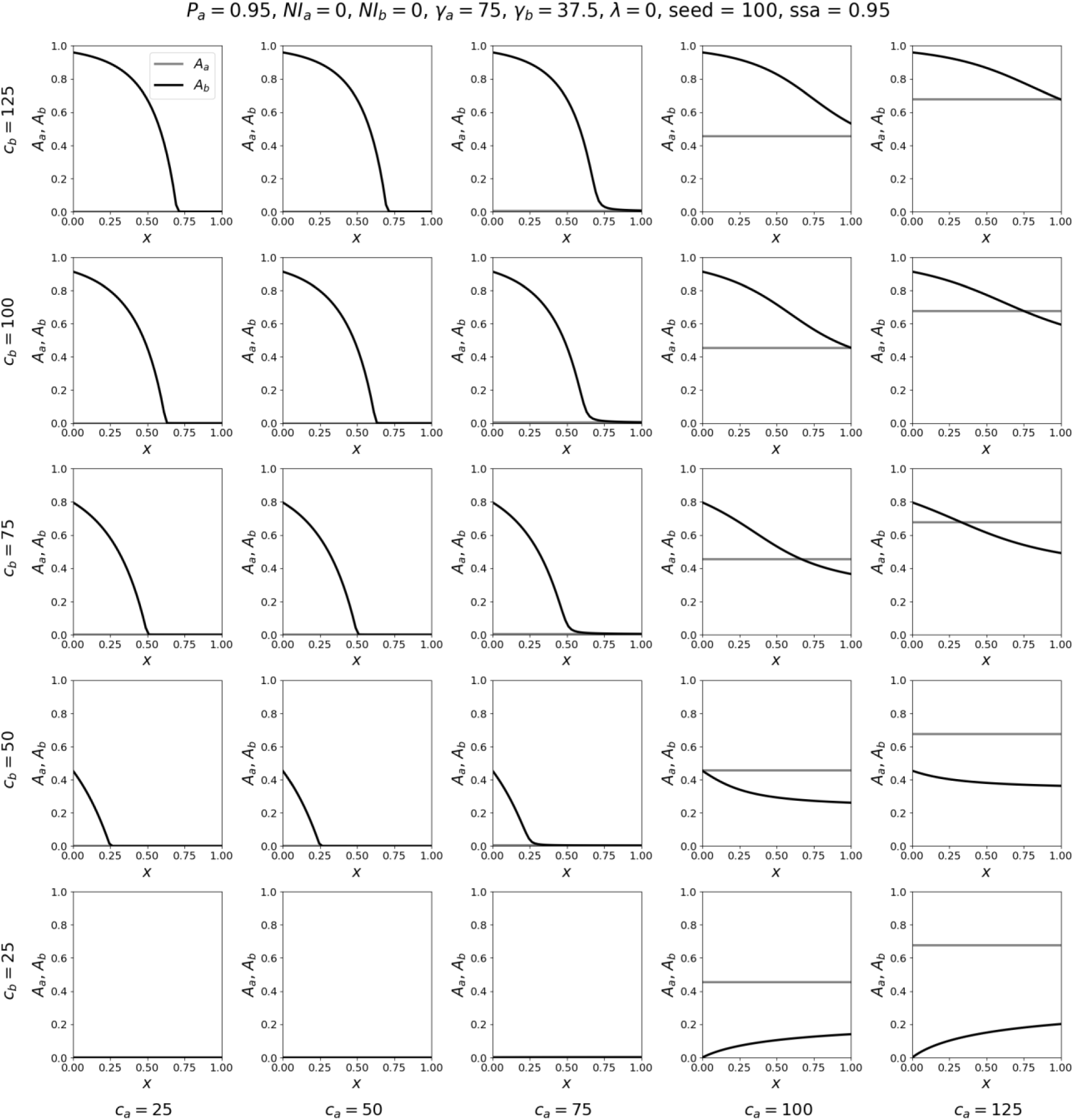
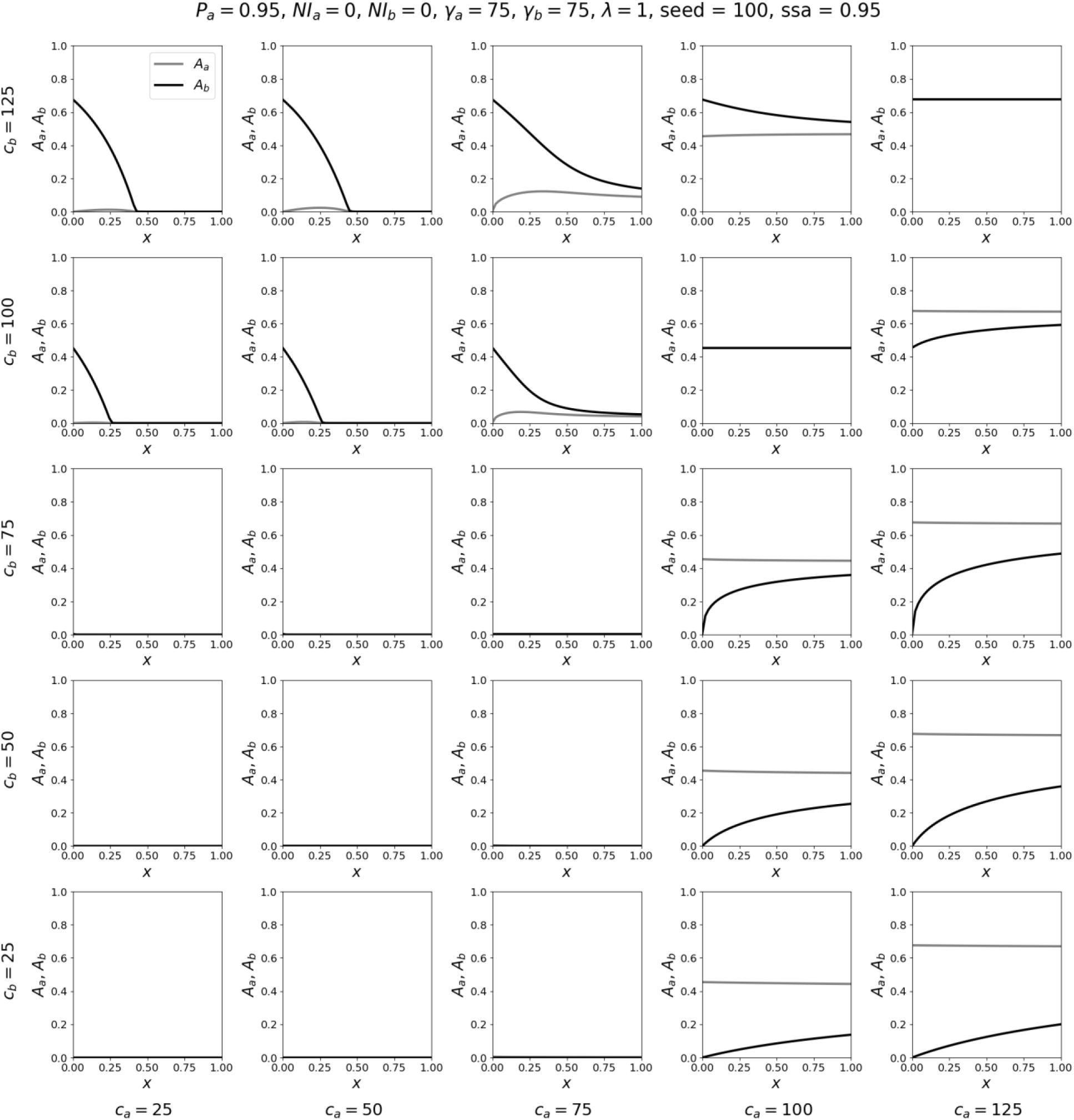
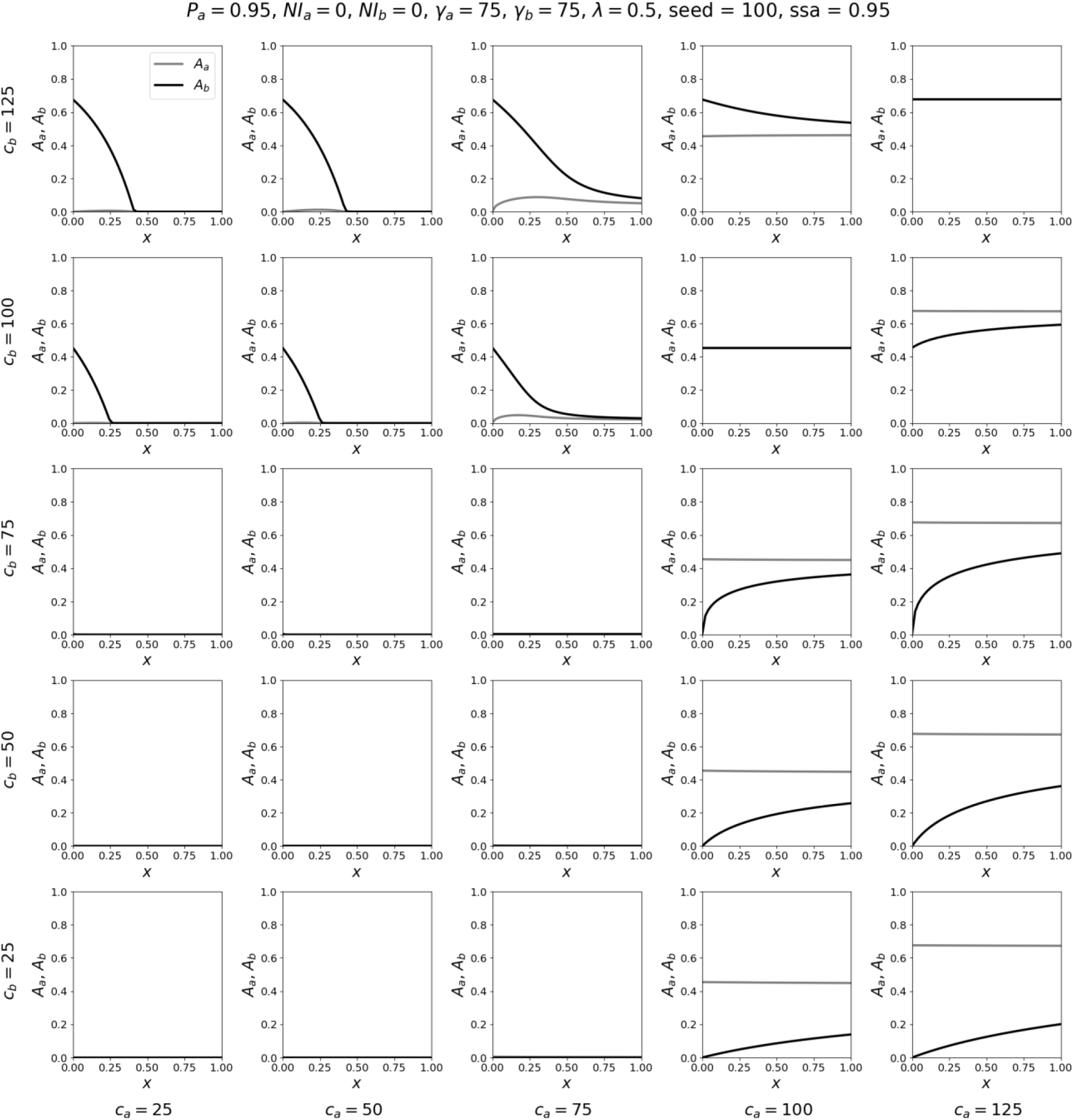
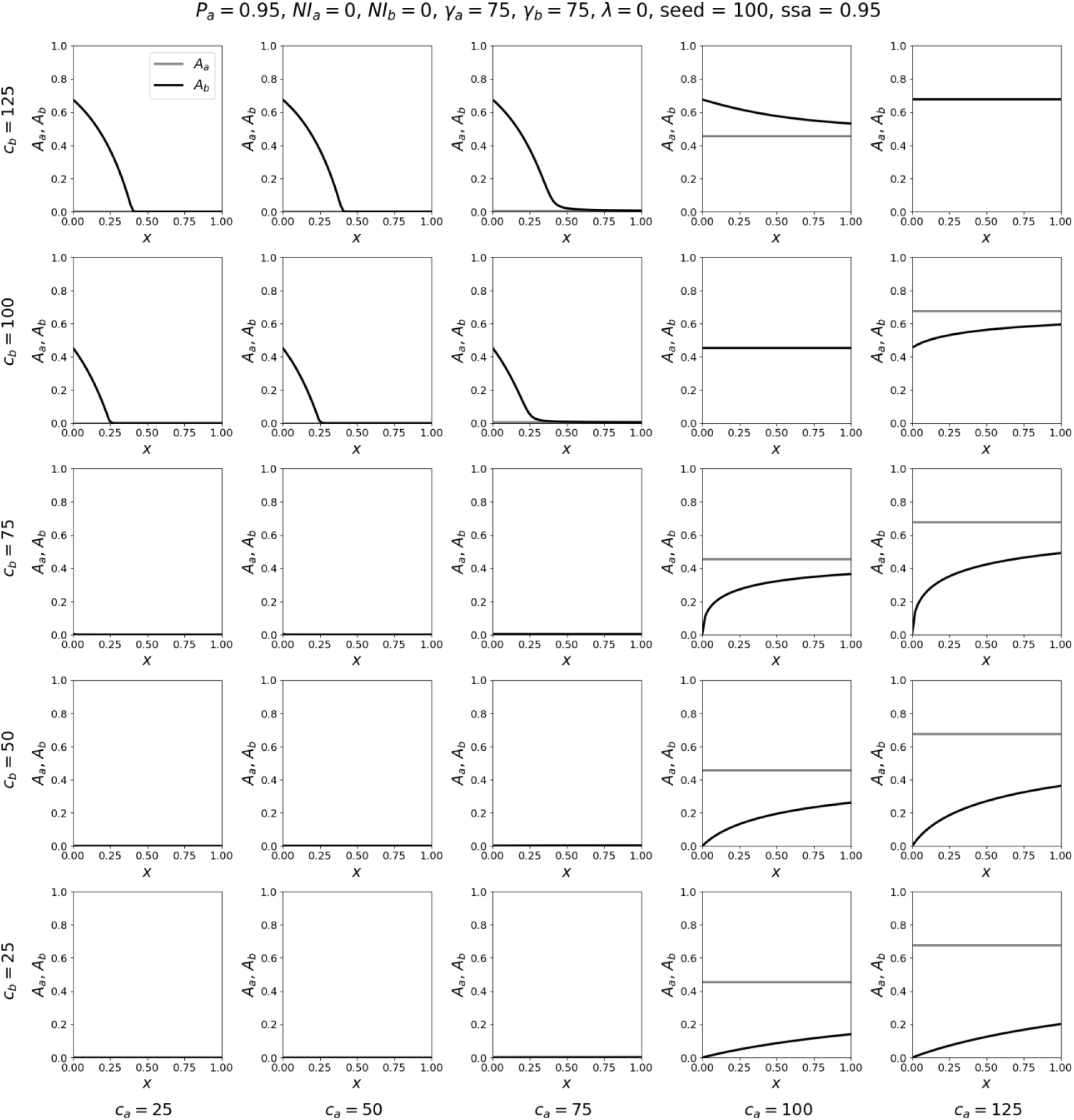

#### A. 3: Epidemic curves for different values of x, for λ = 1

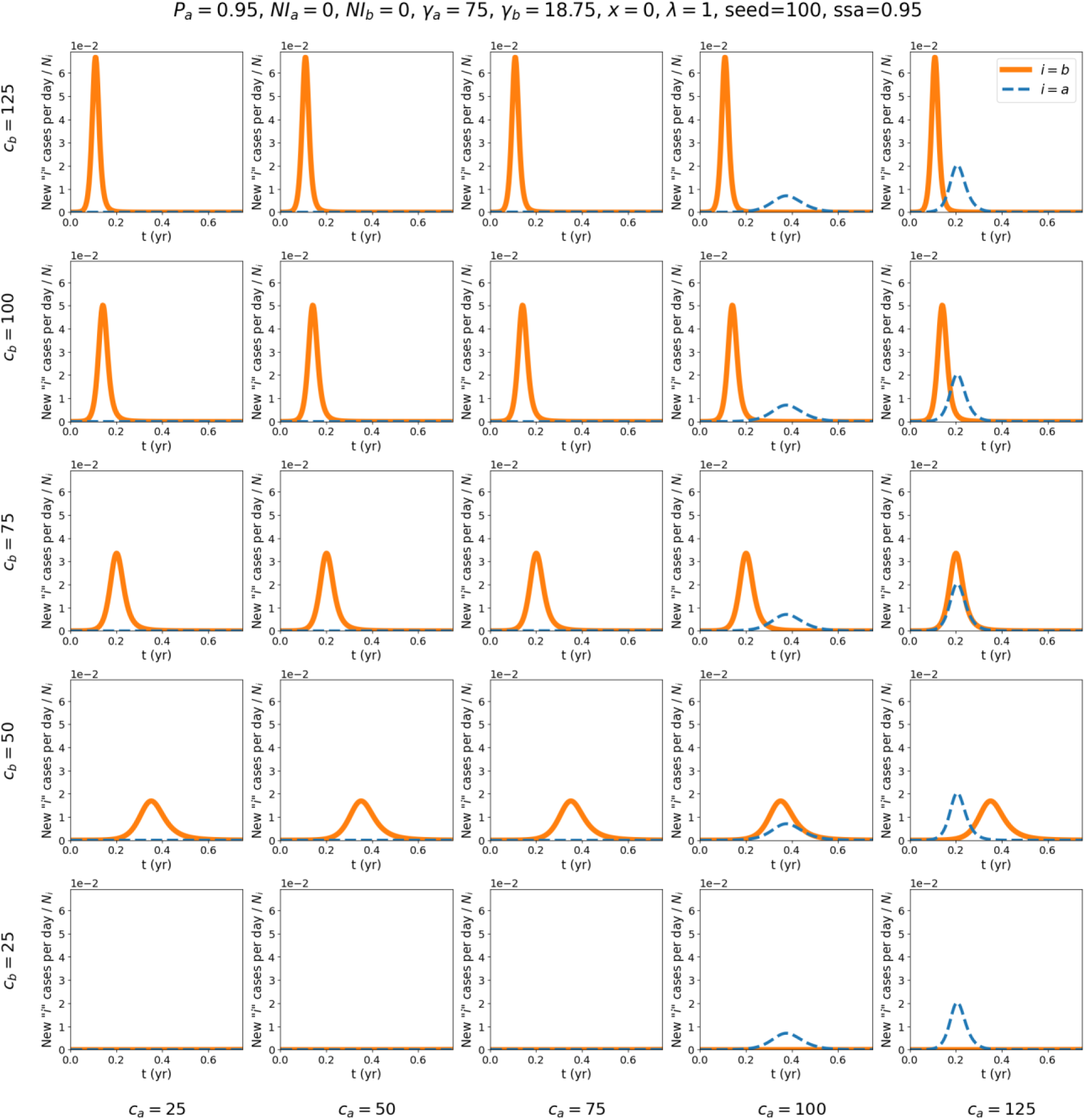
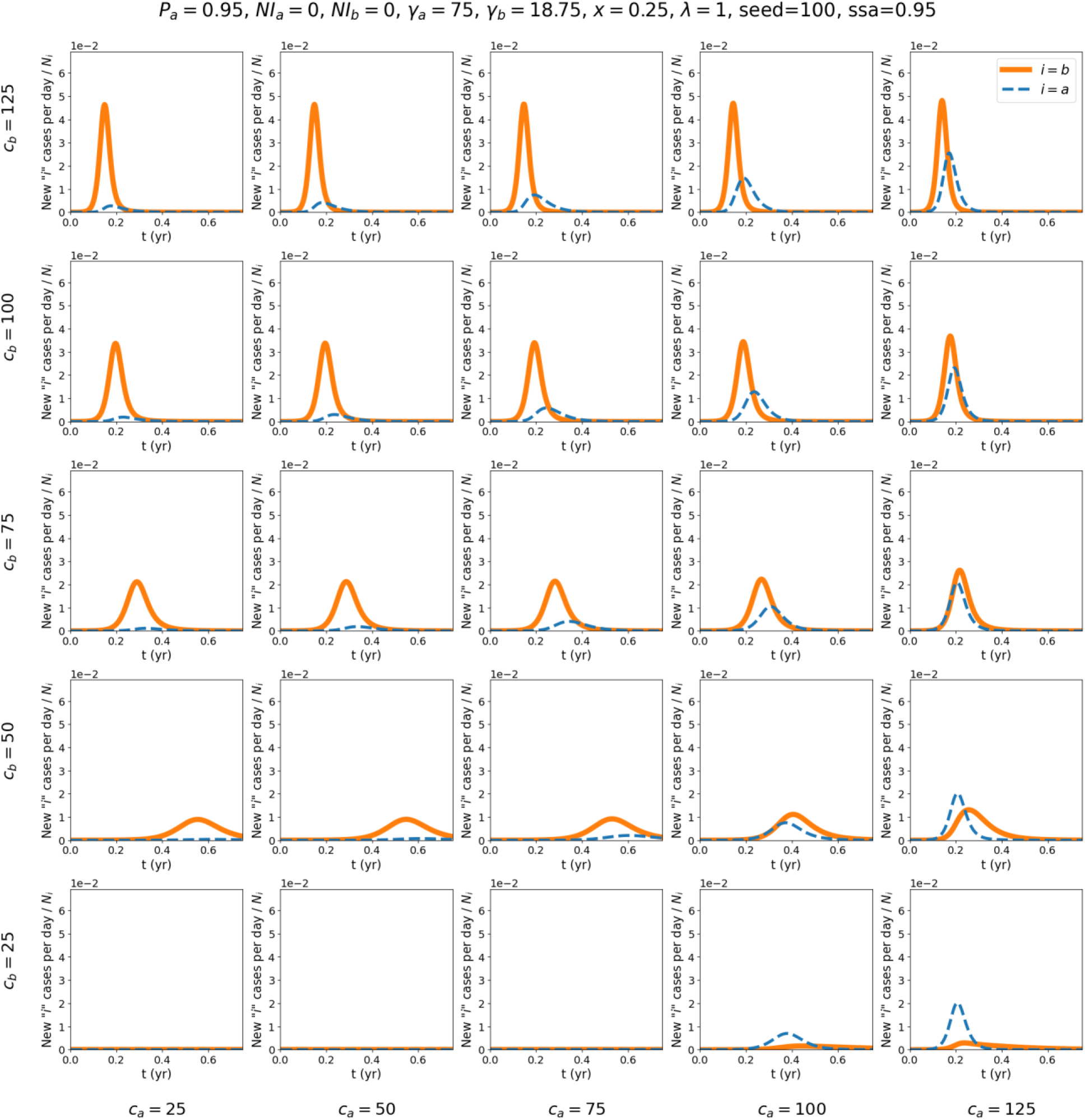
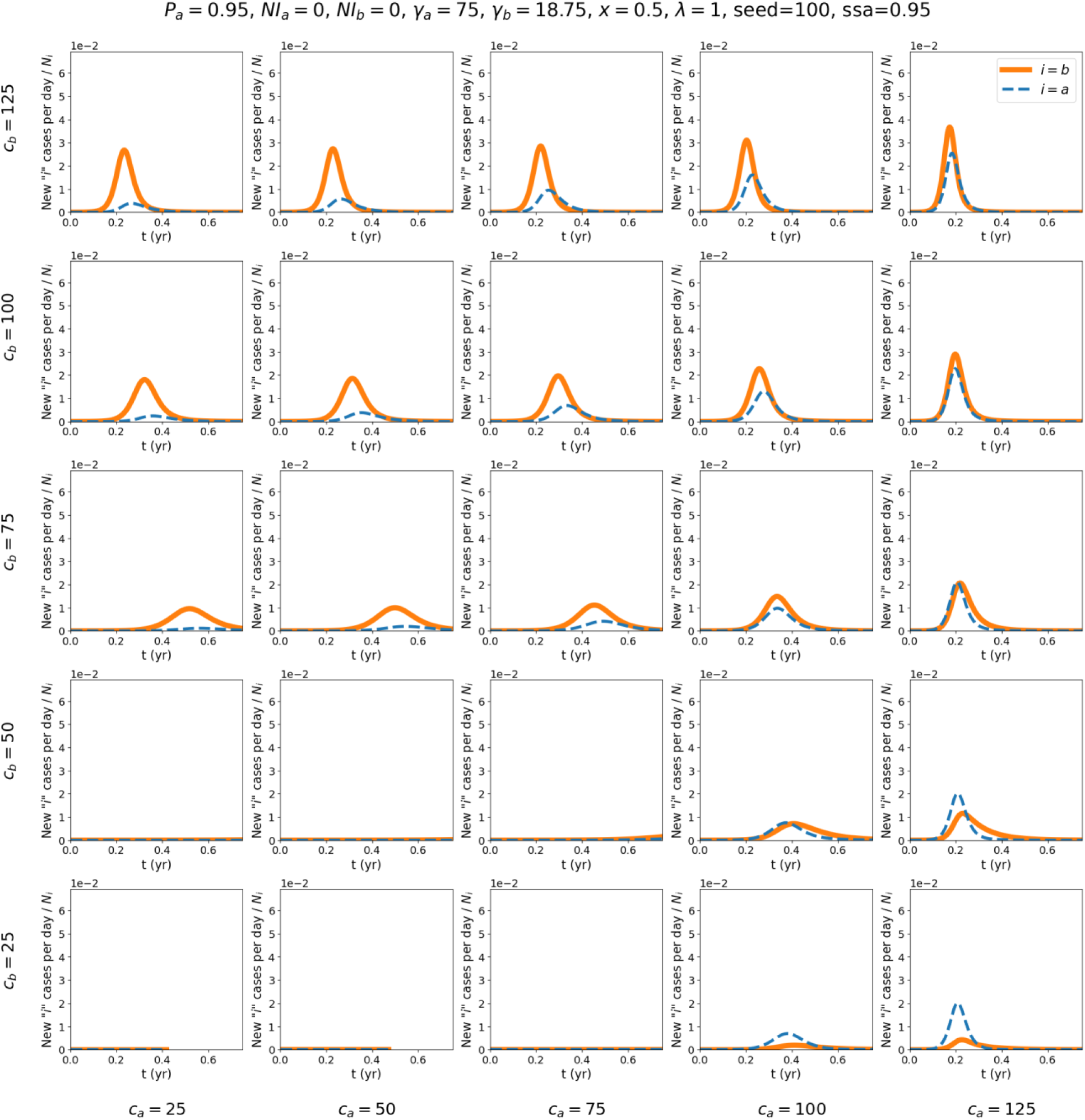
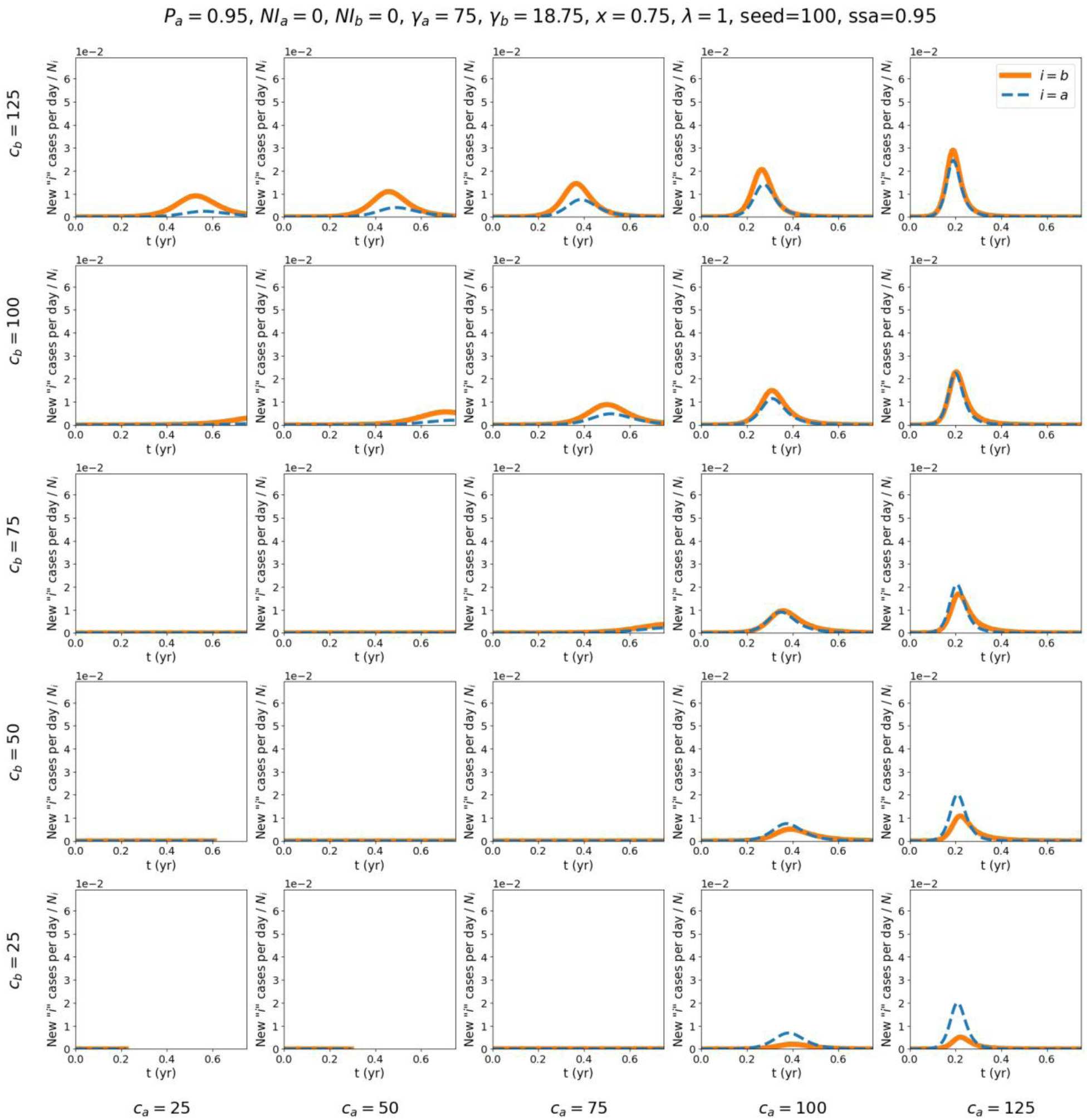
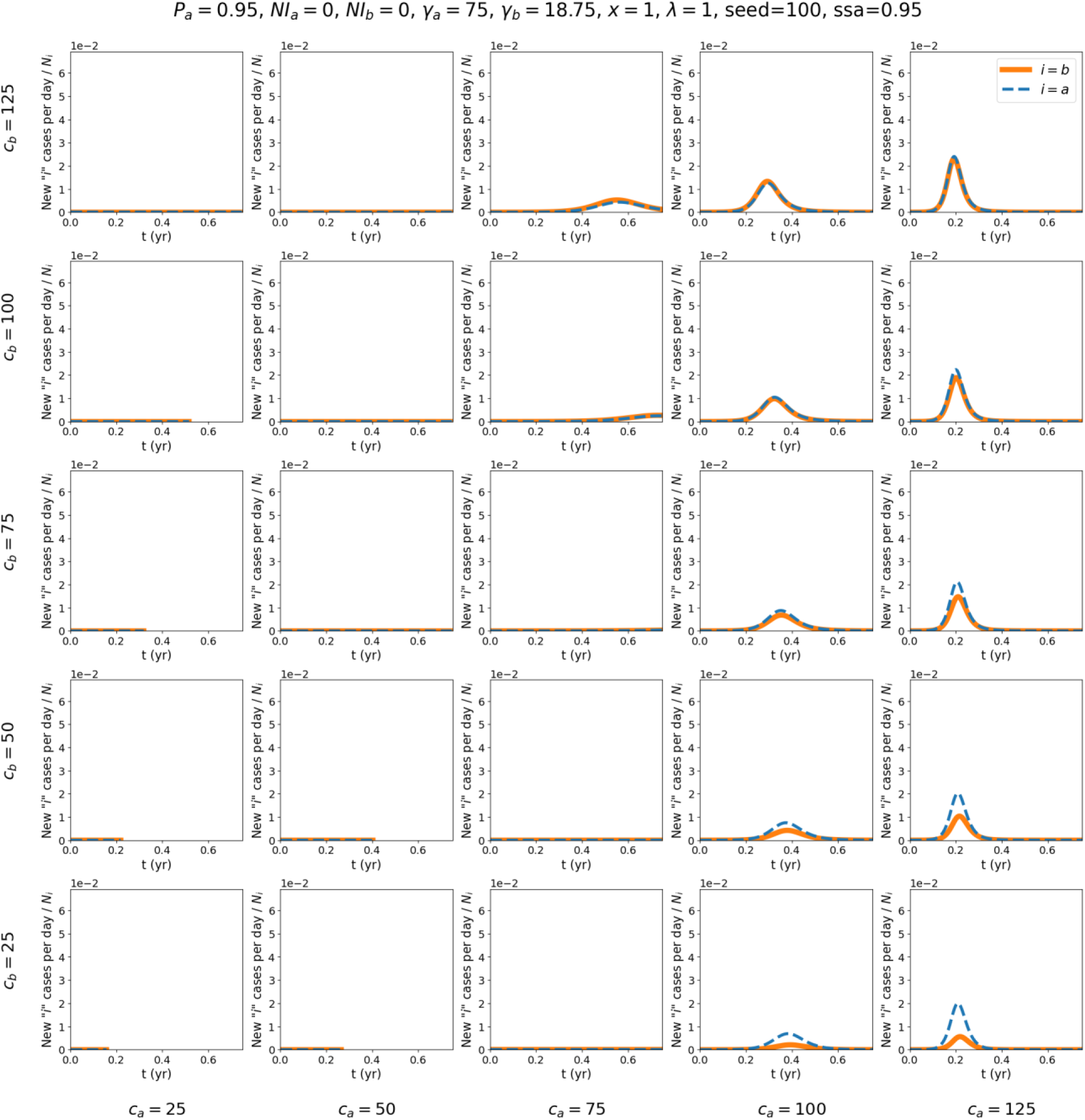
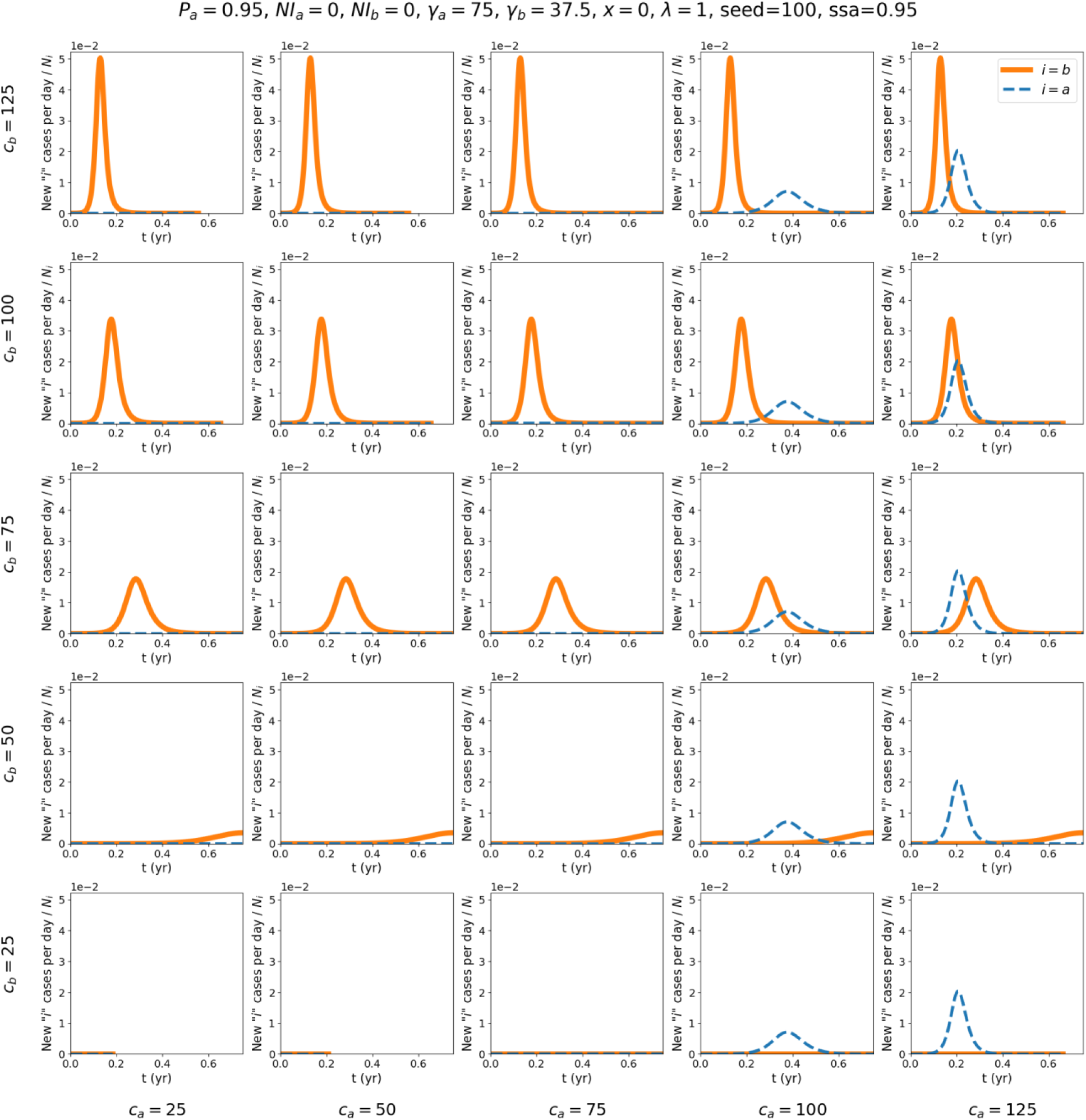
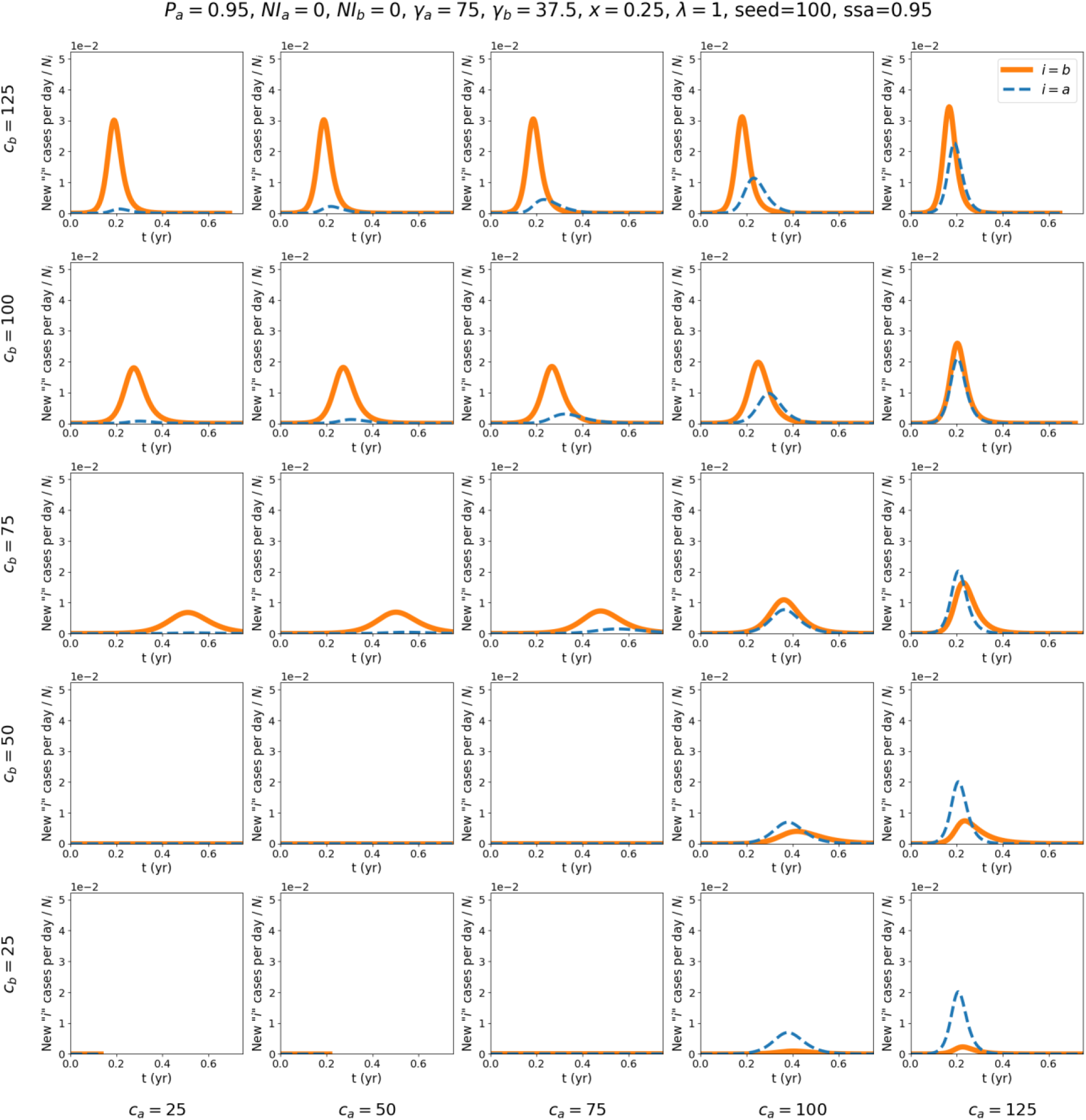
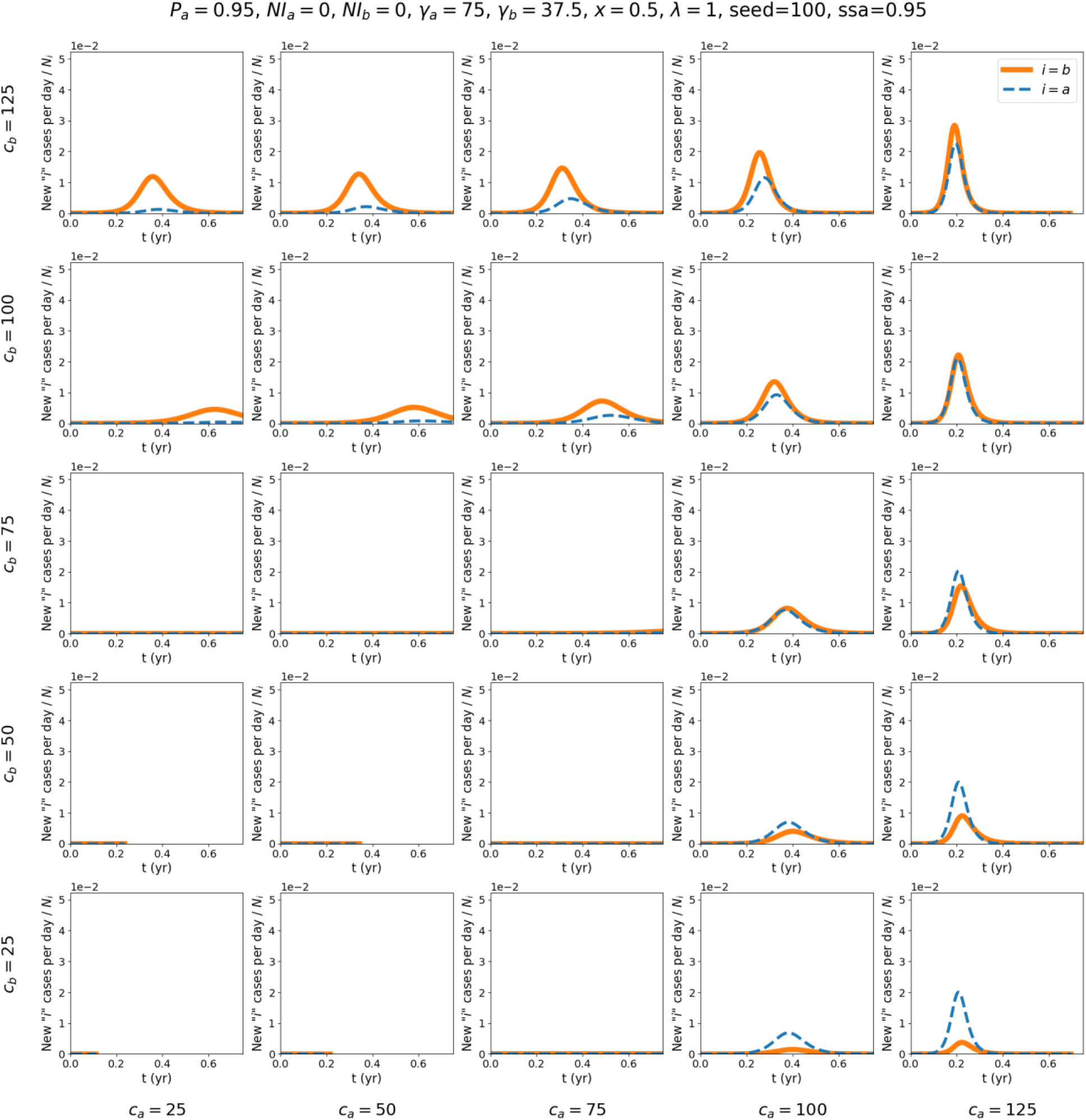
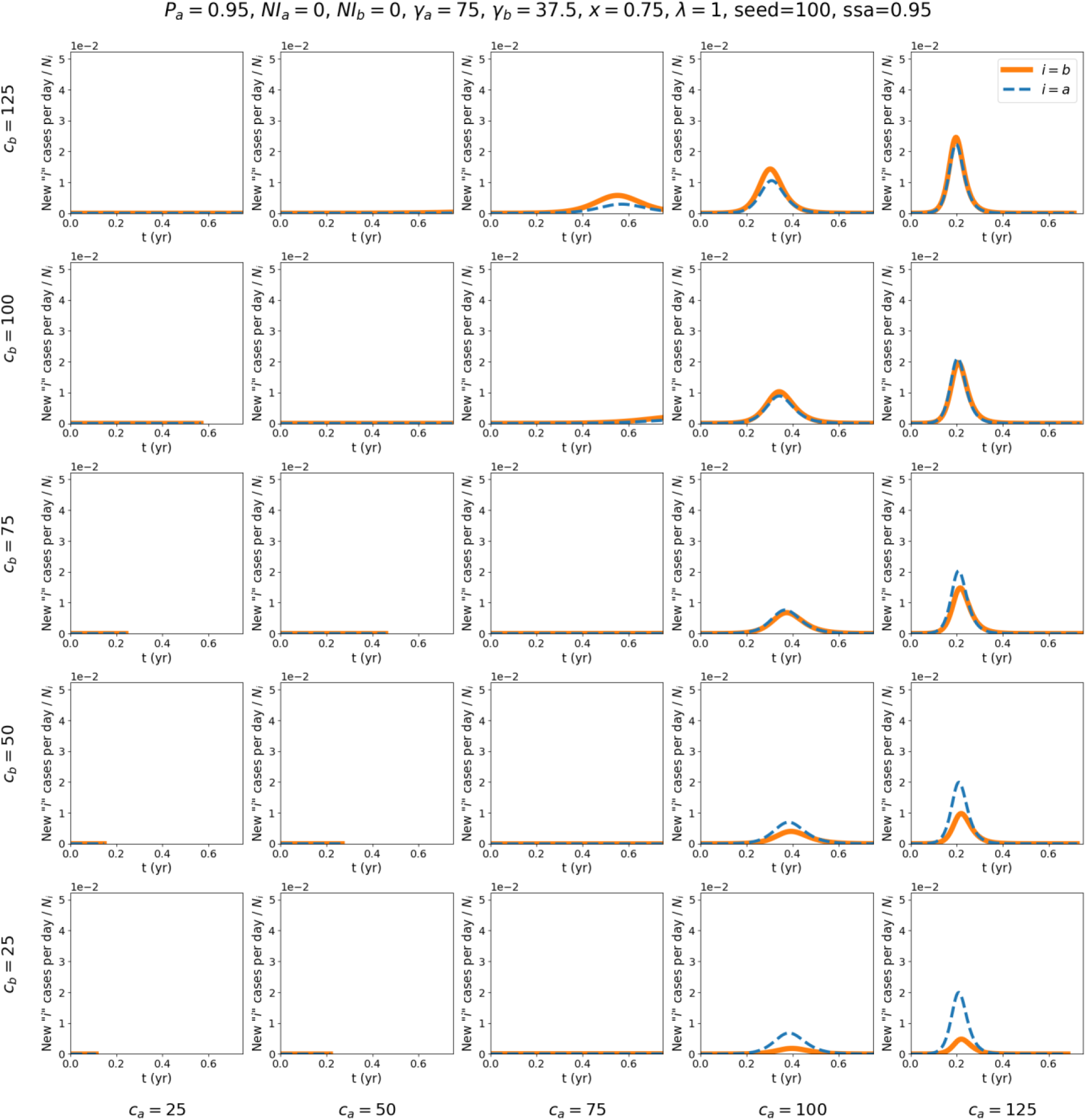
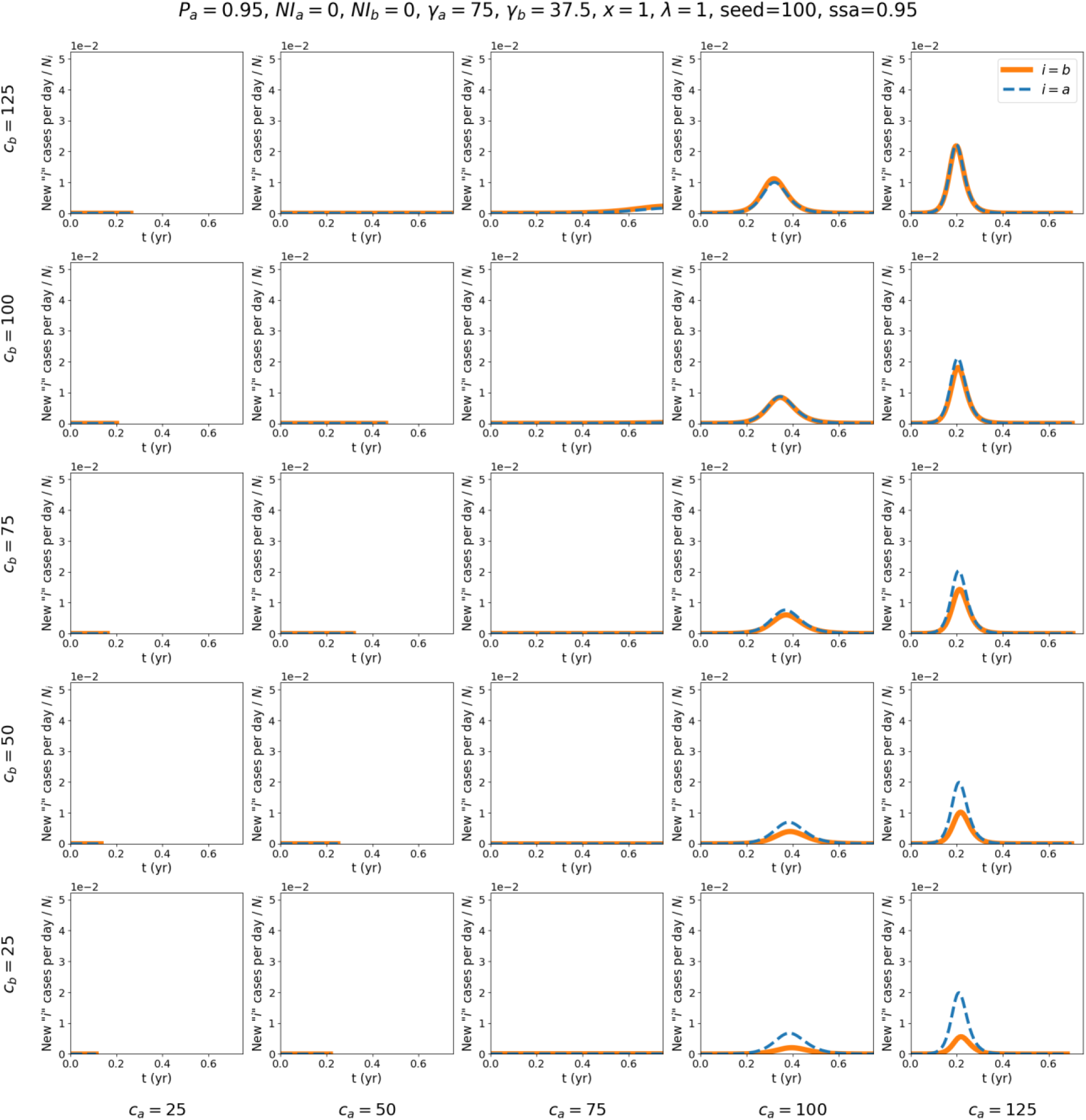
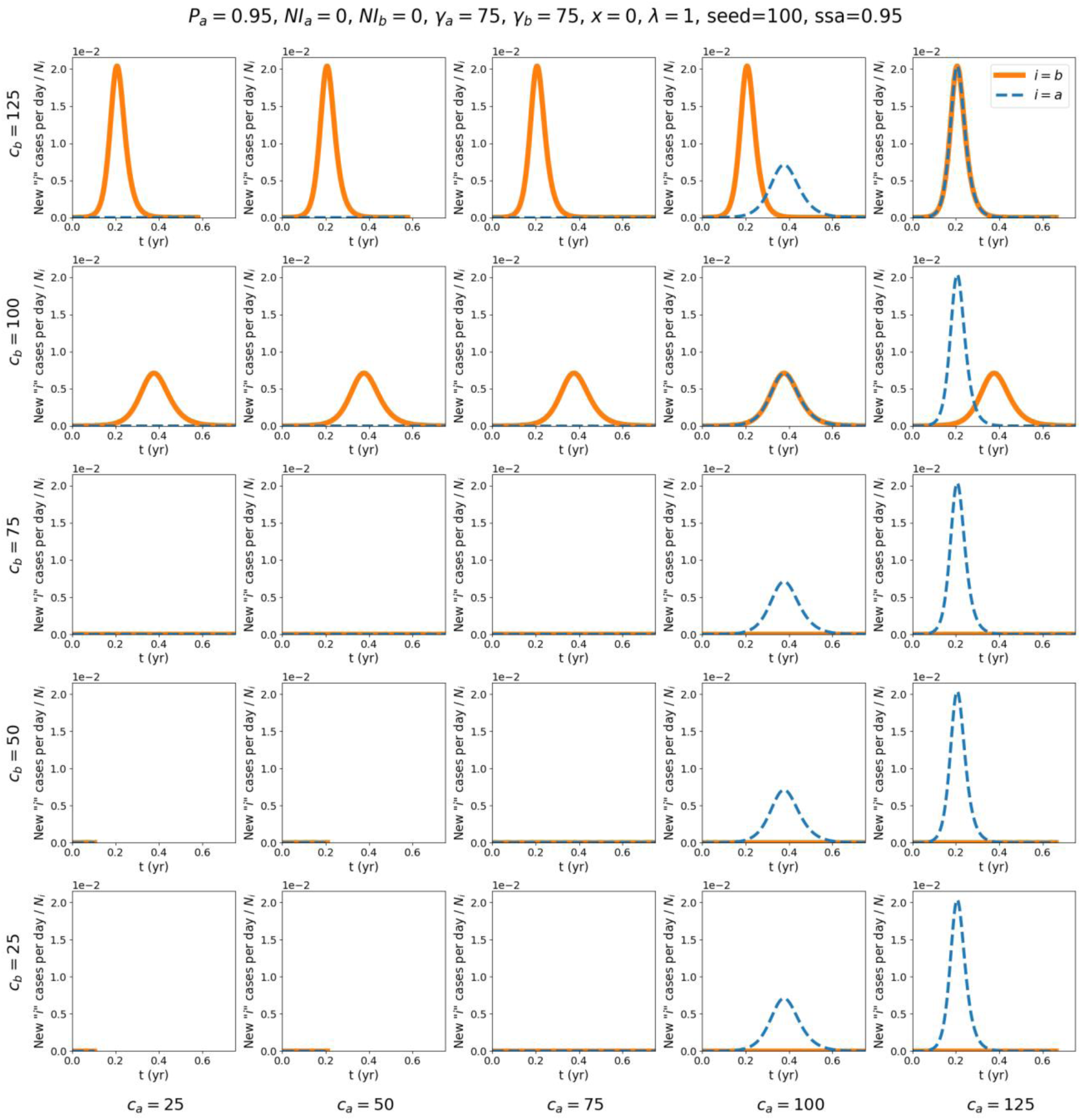
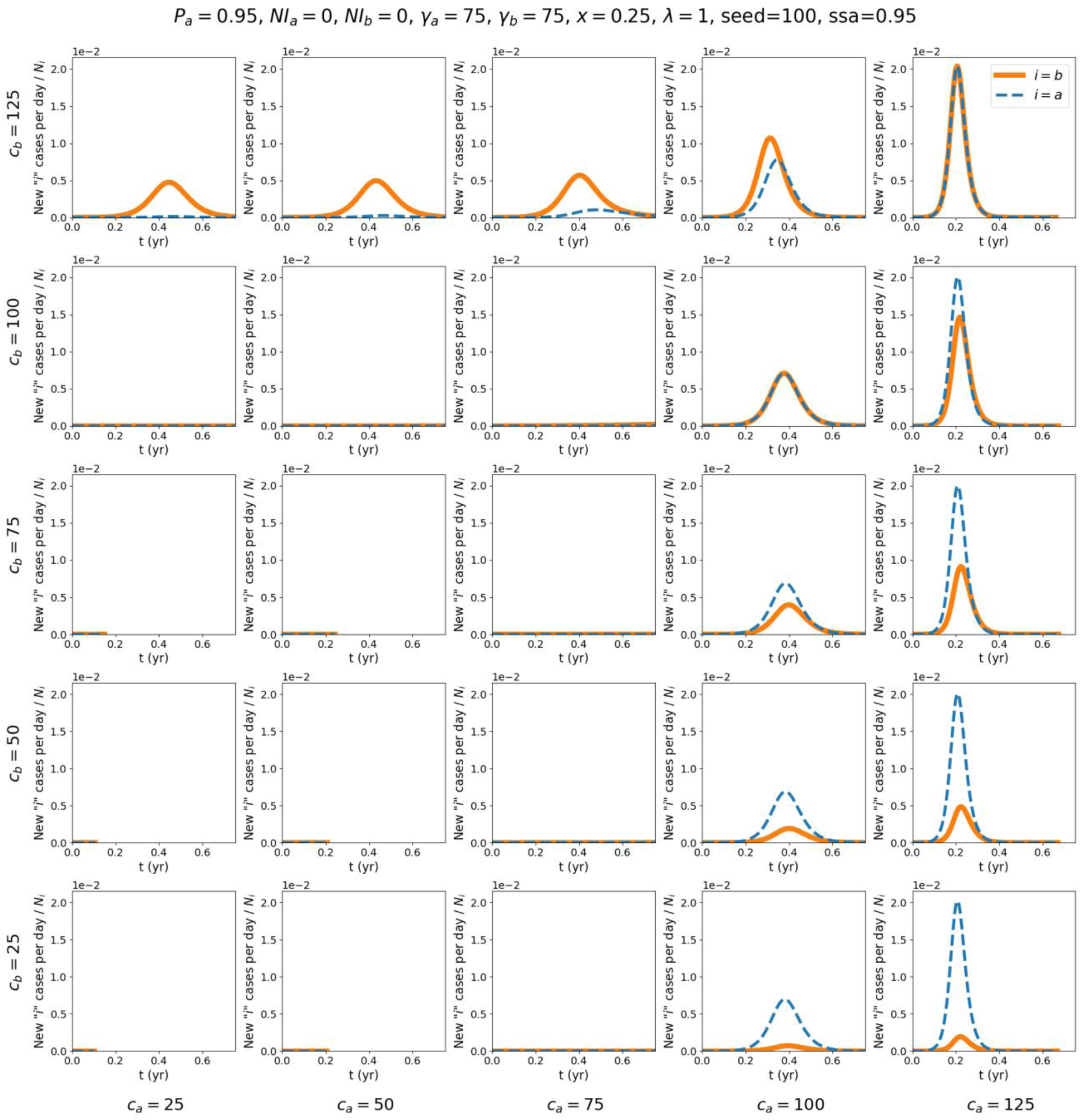
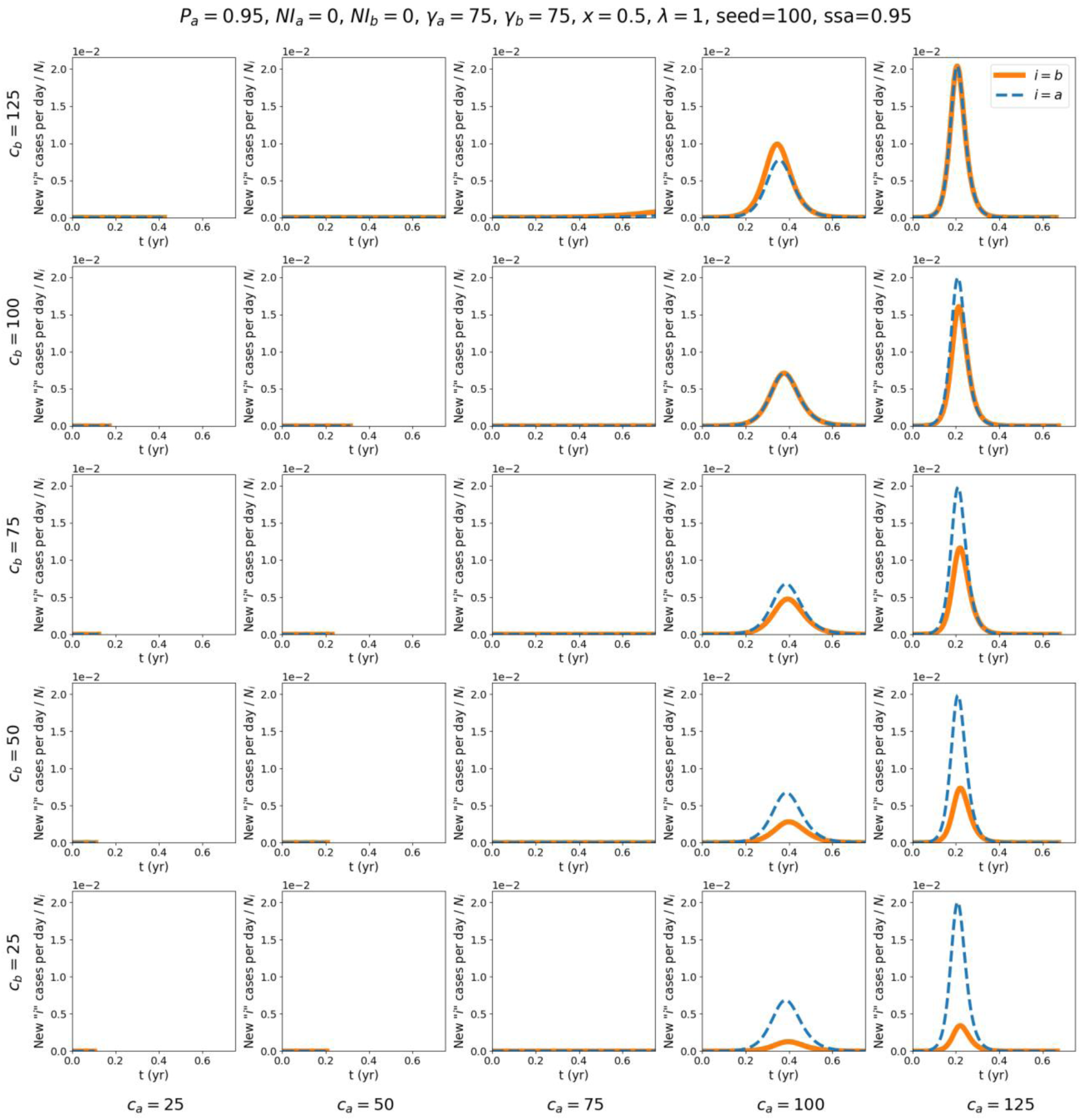
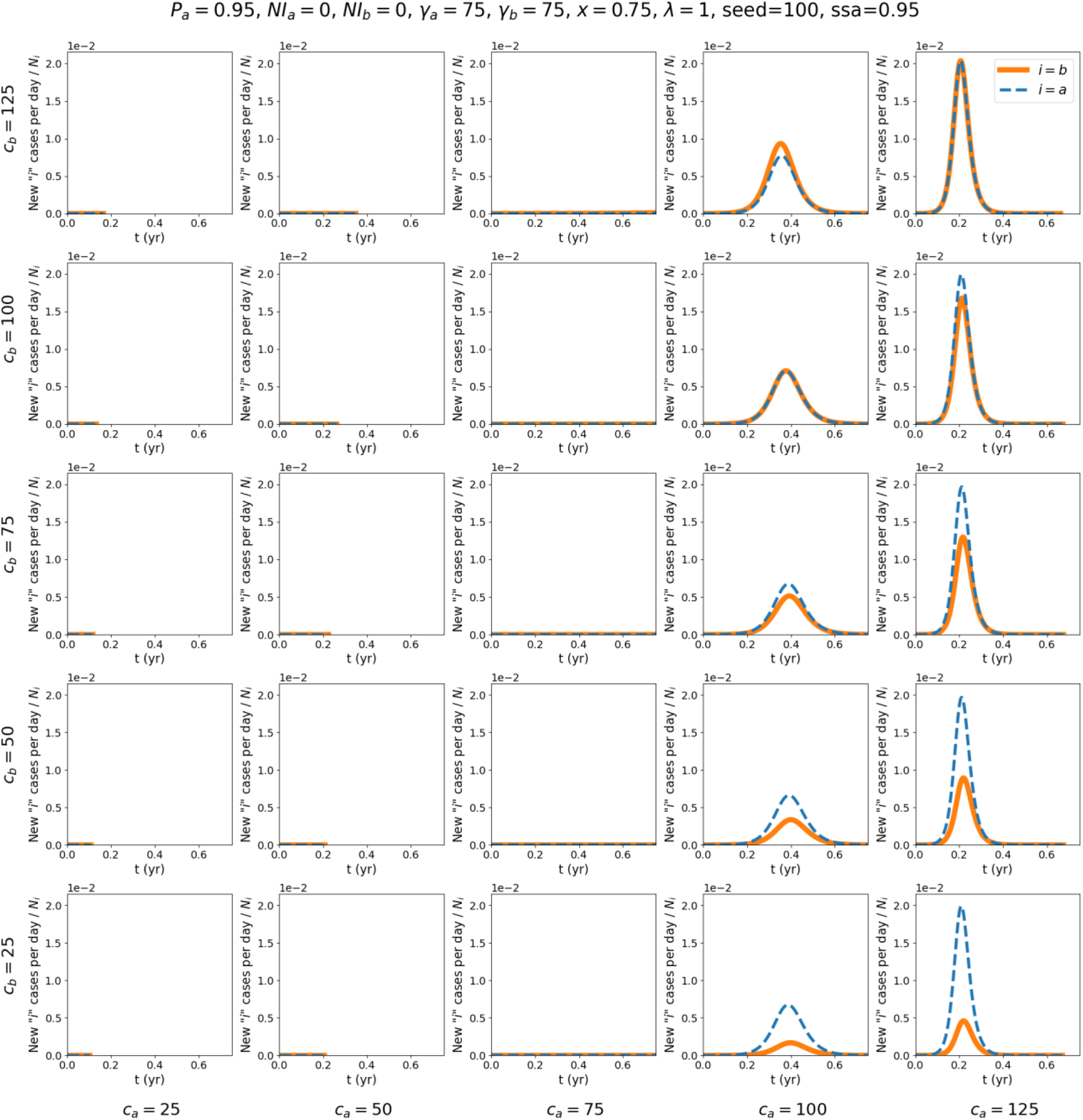
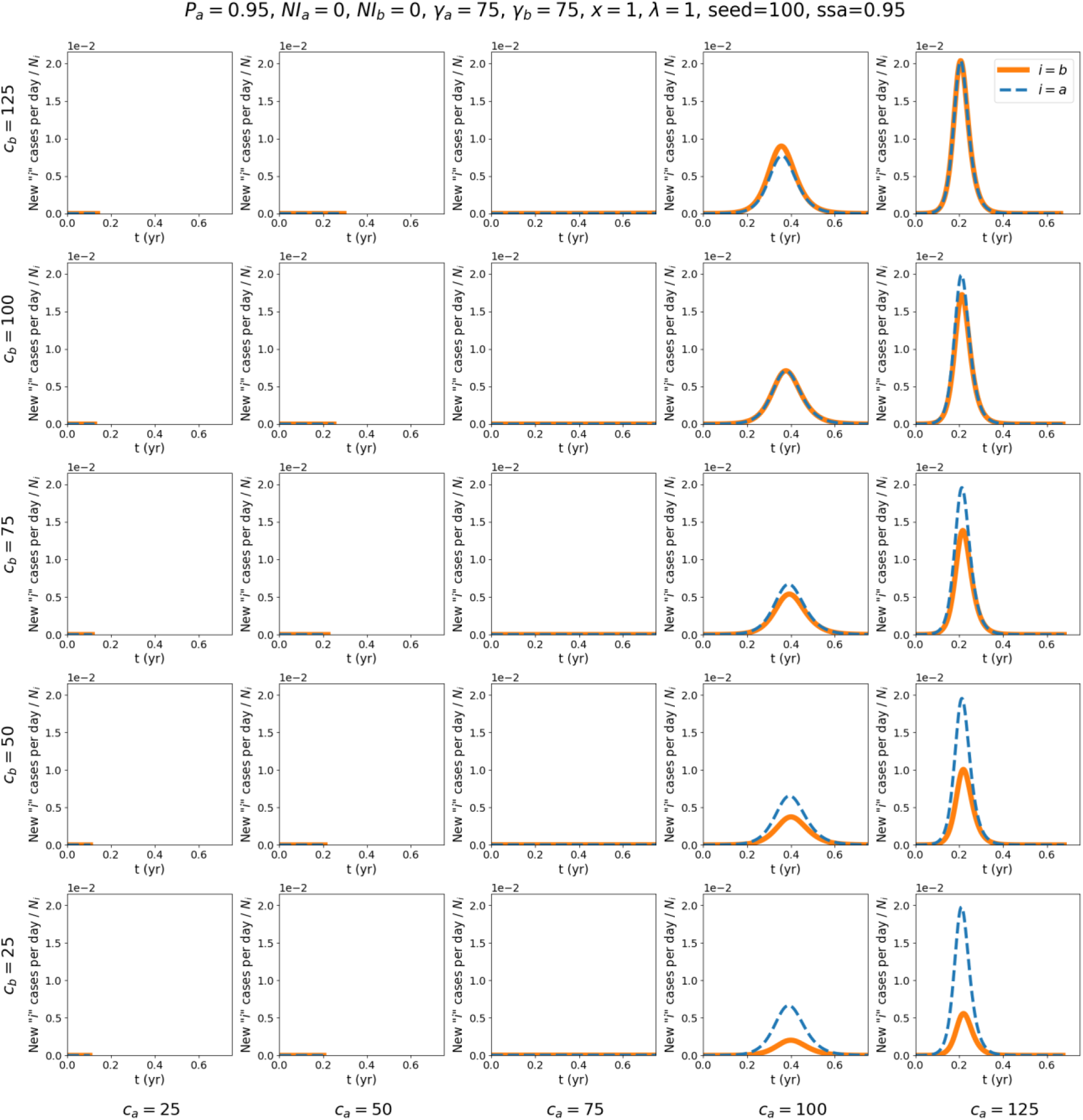

### Appendix B: Results for *P_a_* = 0.8 and *P_a_* = 0.6

This Appendix shows attack-rate contour maps for two different values of *P_a_*, for the same values of *c_a_*, *c_b_*, and *x* used in the main text and elsewhere in the Appendices.

Note that Eqs. 2 and 4 of the main text impose constraints on the *c_ij_*. In some of the contour maps shown in Appendix B and Appendix C, the contour lines end abruptly at points in the (*c_a_*, *c_b_*) plane where these constraints are reached.

For example, for *P_a_* = 0.8, *x* = 1, and γ*_b_* = 18.75, the point (*c_a_* = 20, *c_b_* = 100) is unphysical, because *x* = 1 implies that *c_ba_* = *c_b_* = 100 (see Eq. 4, main text), and λ = 1, *P_a_* = 0.8, and *c_ba_* = 100 are such (from Eq. 2, main text) that *c_ab_* = λ*c_ba_*(1-*P_a_*)/*P_a_* = 25 > *c_a_* which is unphysical, since *c_a_* = *c_aa_* + *c_ab_* and both *c_aa_* ≥ 0 and *c_ab_* ≥ 0. Accordingly, the contour lines in the contour map for *P_a_* = 0.8, *x* = 1, and γ*_b_* = 18.75 (lower-left panel in the first figure in section B.1, below) end before the unphysical point (*c_a_* = 20, *c_b_* = 100).

#### B. 1: Attack-rate contour maps for different values of λ and for P_a_ = 0.8

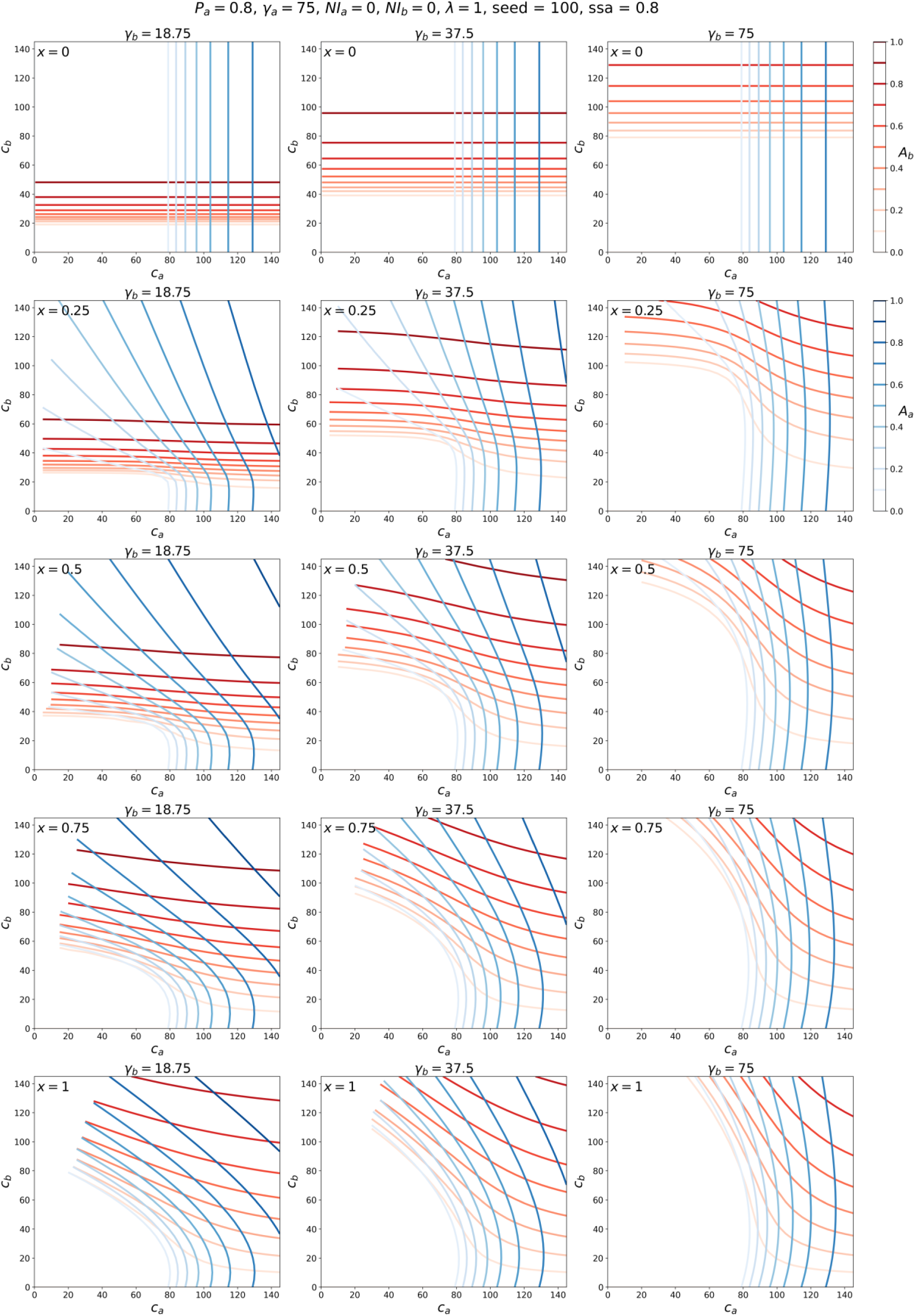
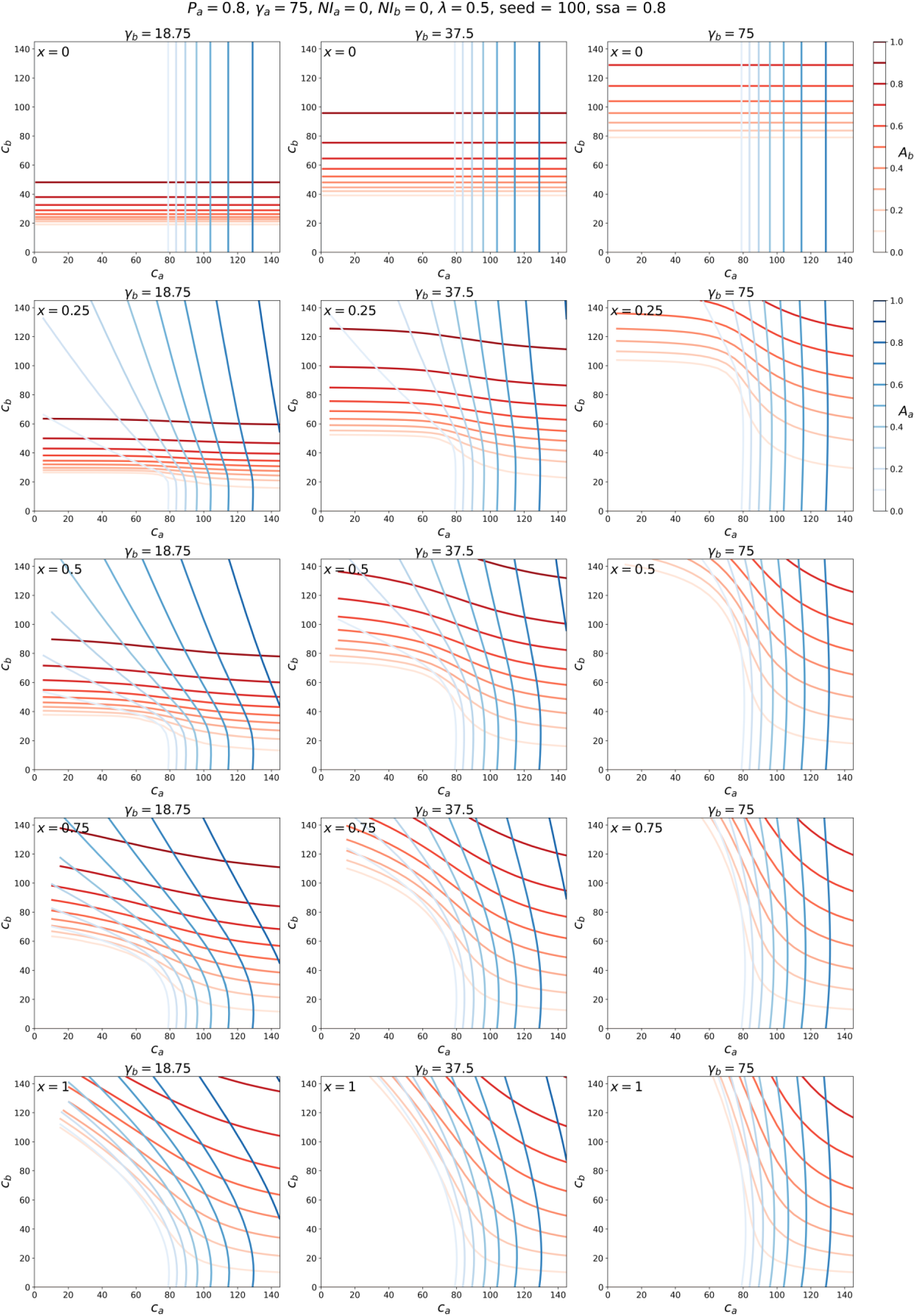
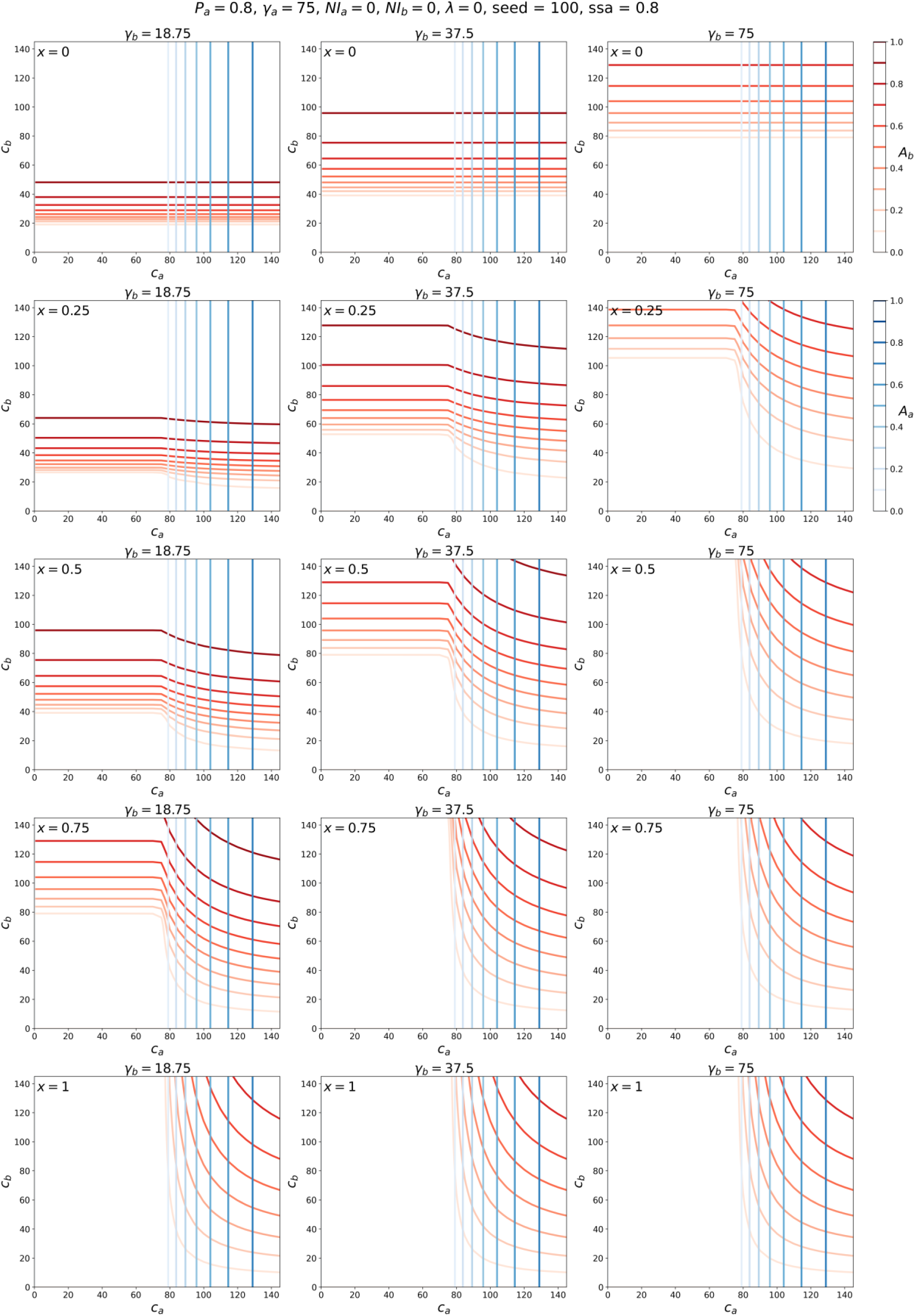

#### B. 2: Attack-rate contour maps for different values of λ and for P_a_ = 0.6

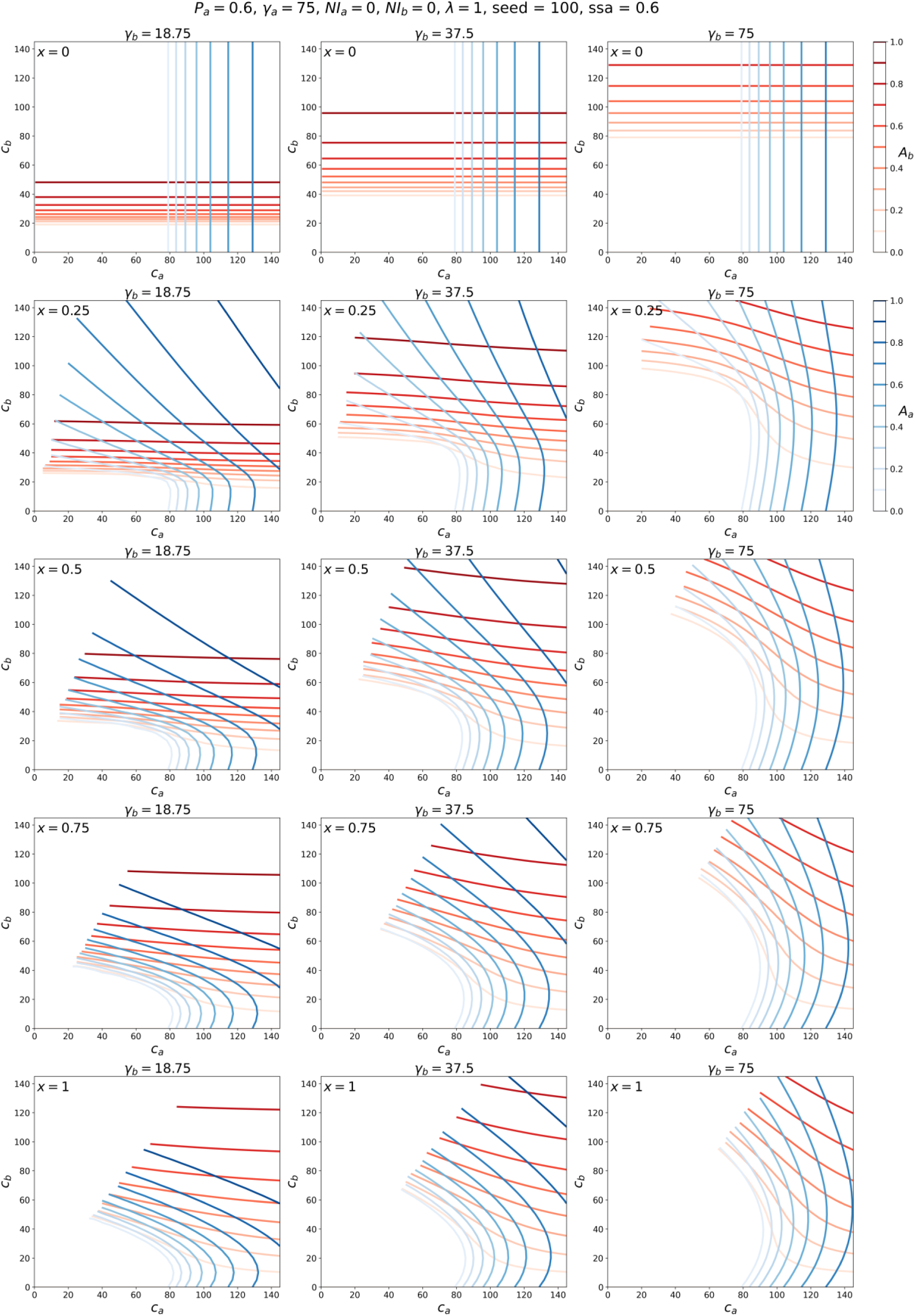
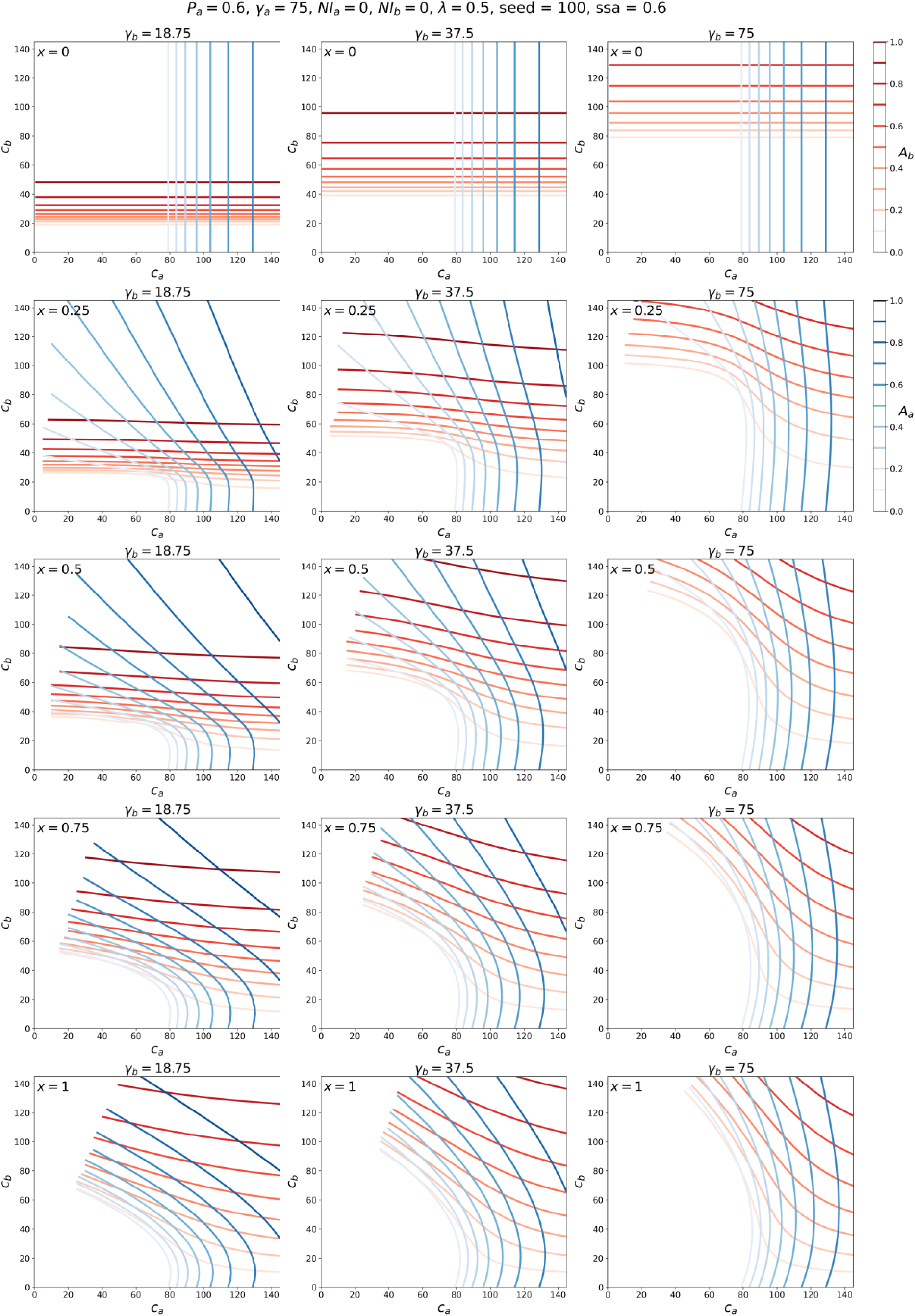
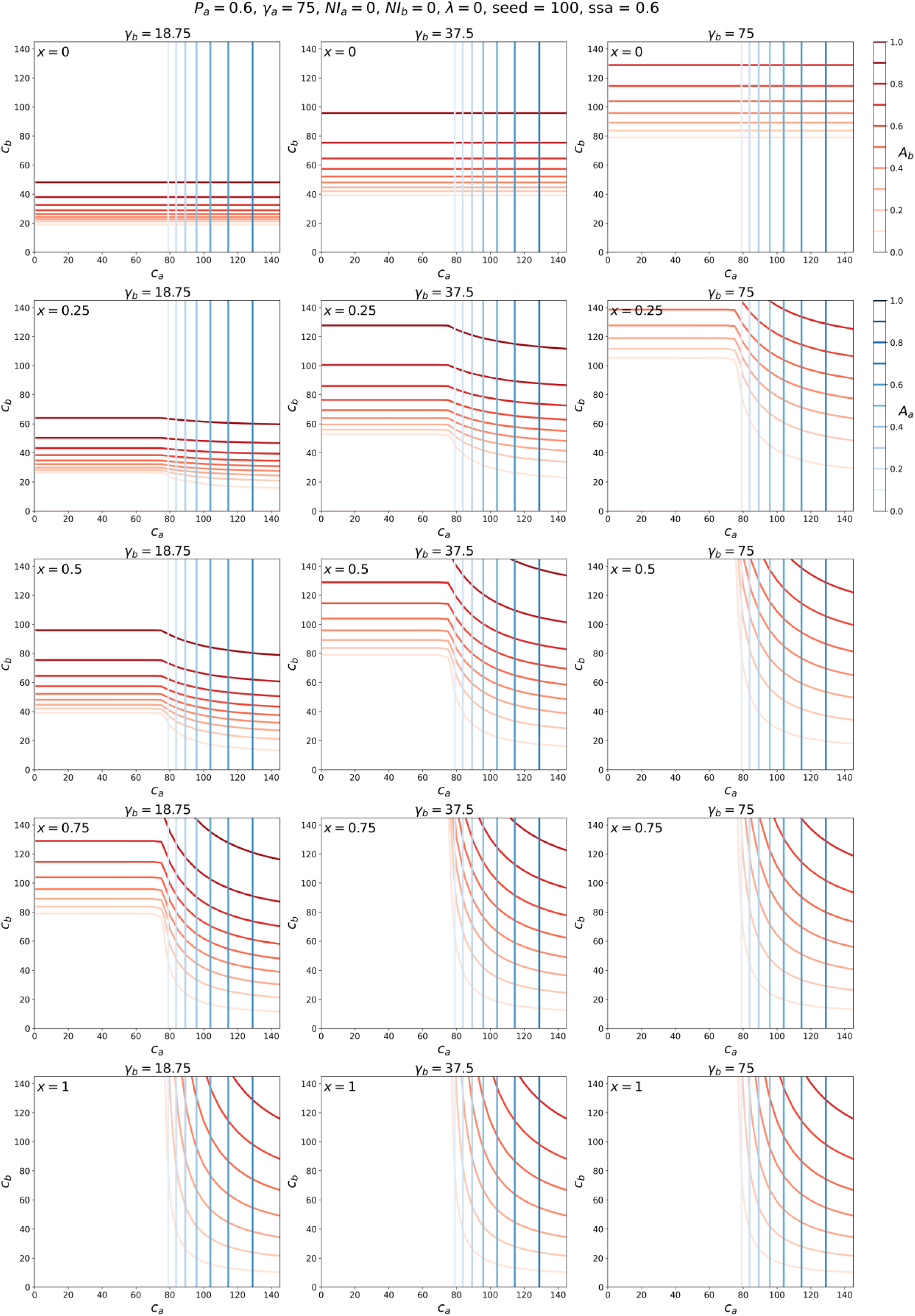

### Appendix C: Varying seed distribution

The figures in Appendix C show contour maps for *P_a_* = 0.95, *P_a_* = 0.8, and *P_a_* = 0.6, with the initial 100 infected “seed” individuals placed entirely in the *a* population (ssa = 1). That is, none of the *b* individuals are initially infected, in the simulations shown in Appendix C.

As can be seen from the figures below, placing all “seed” individuals in the *a* group has no effect on the resulting attack rates as functions of *c_a_*, *c_b_*, and *x*, except for the trivial case of *x* = 0, in which it is impossible for any *b* person to become infected, since *x* = 0 means that *c_ba_* = 0.

#### C. 1: Attack-rate contour maps for different values of λ, for P_a_ = 0.95, seed = 100, and ssa = 1

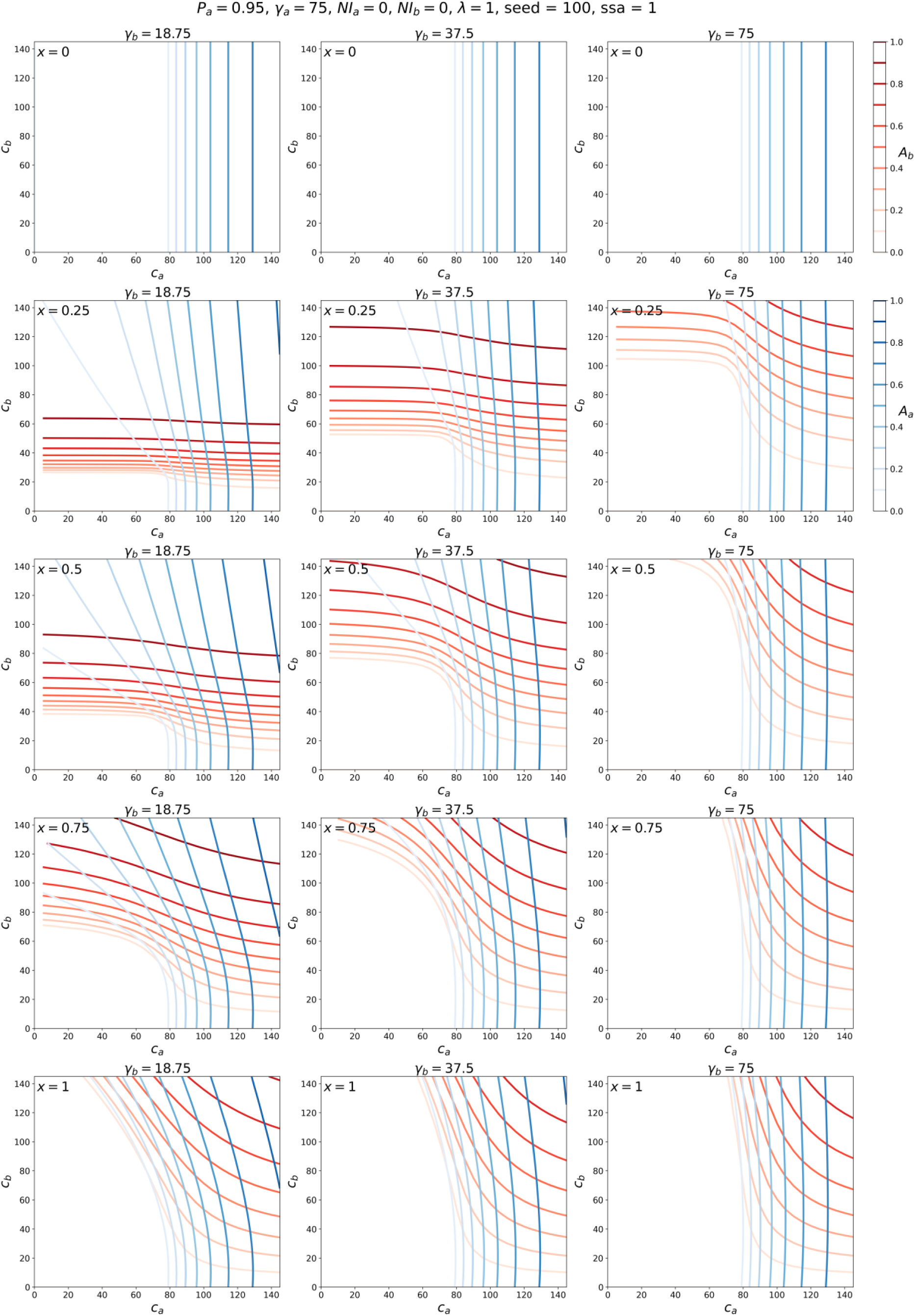
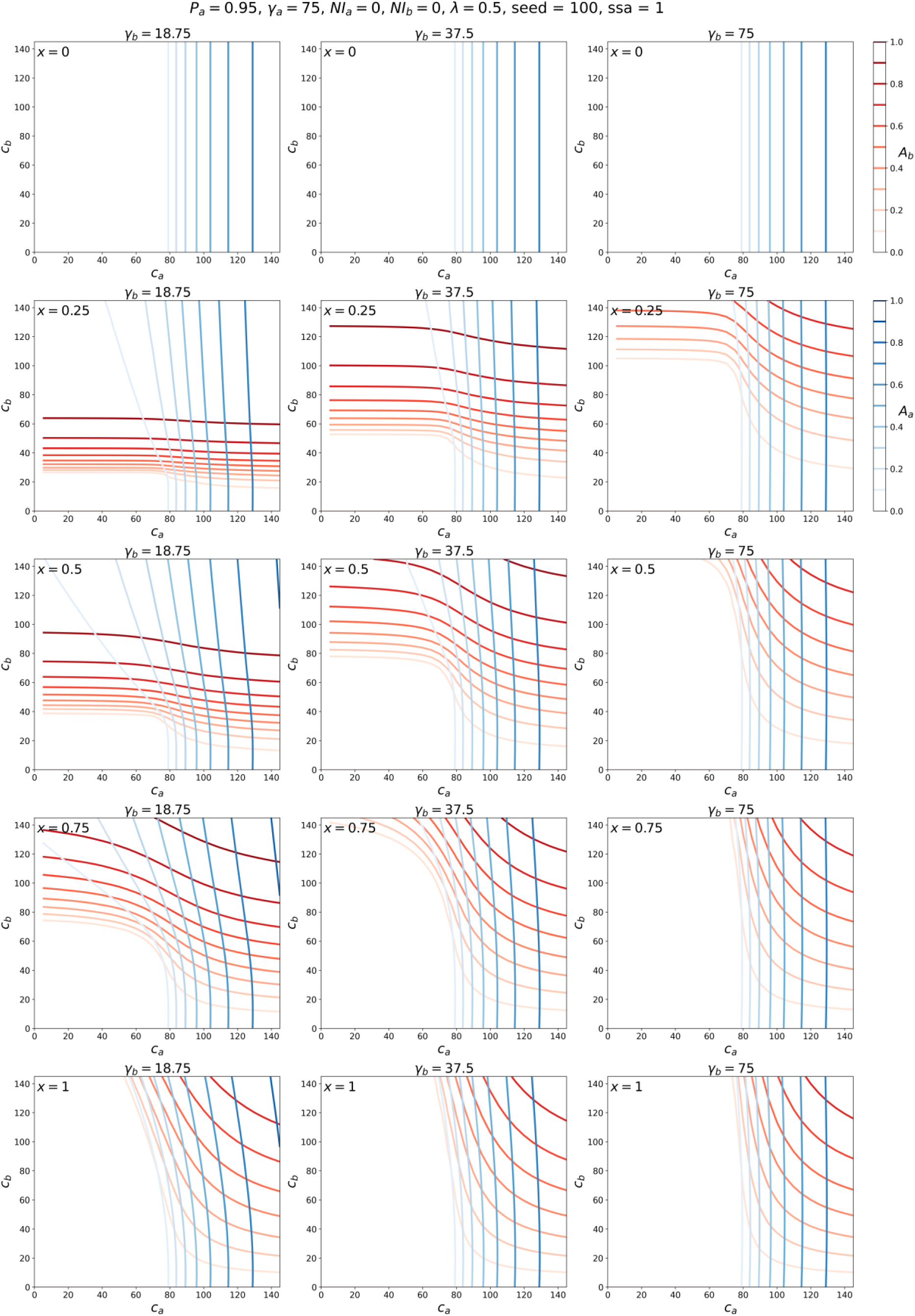
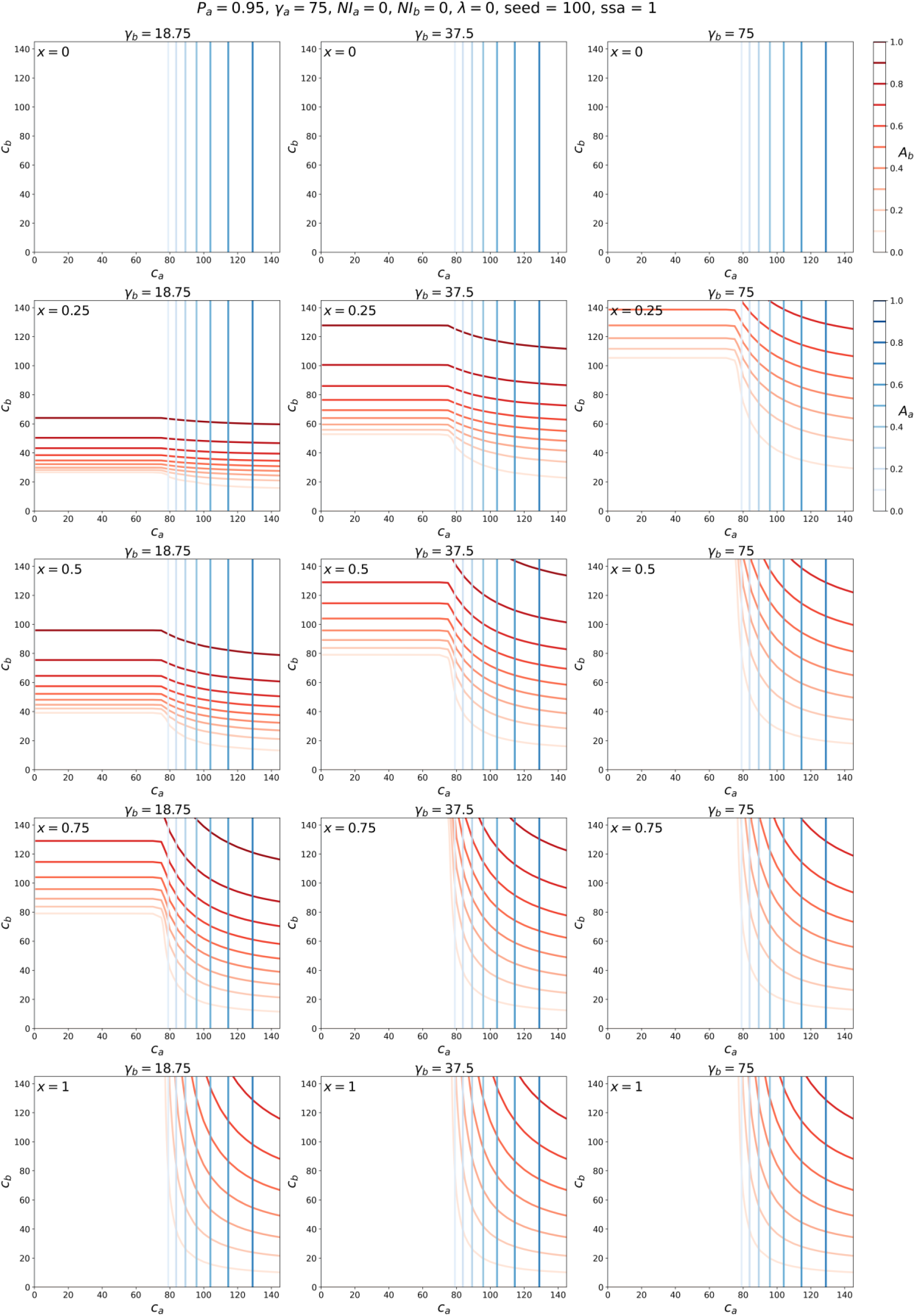

#### C.2: Attack-rate contour maps for different values of λ, for P_a_ = 0.8, seed = 100, and ssa = 1

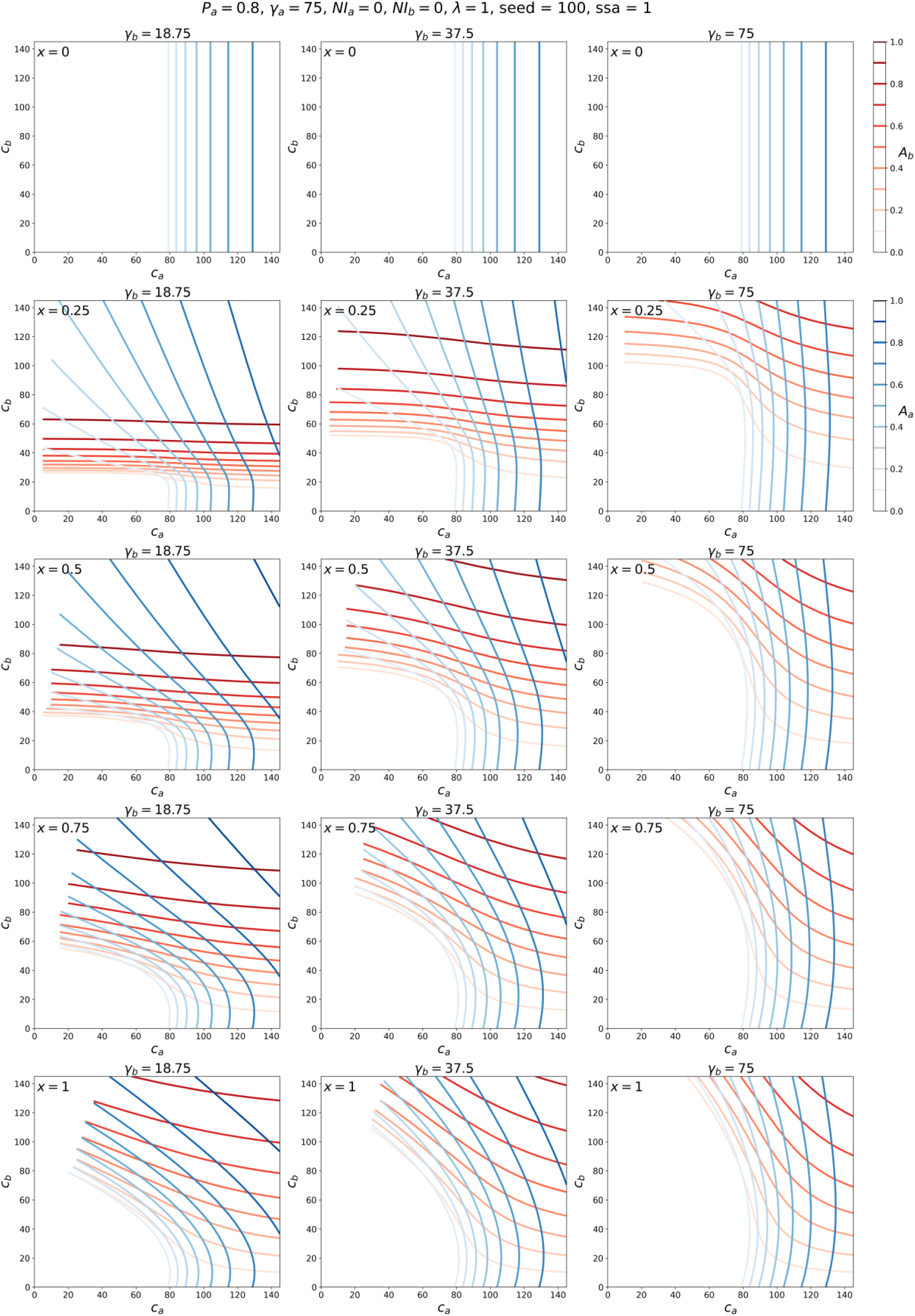
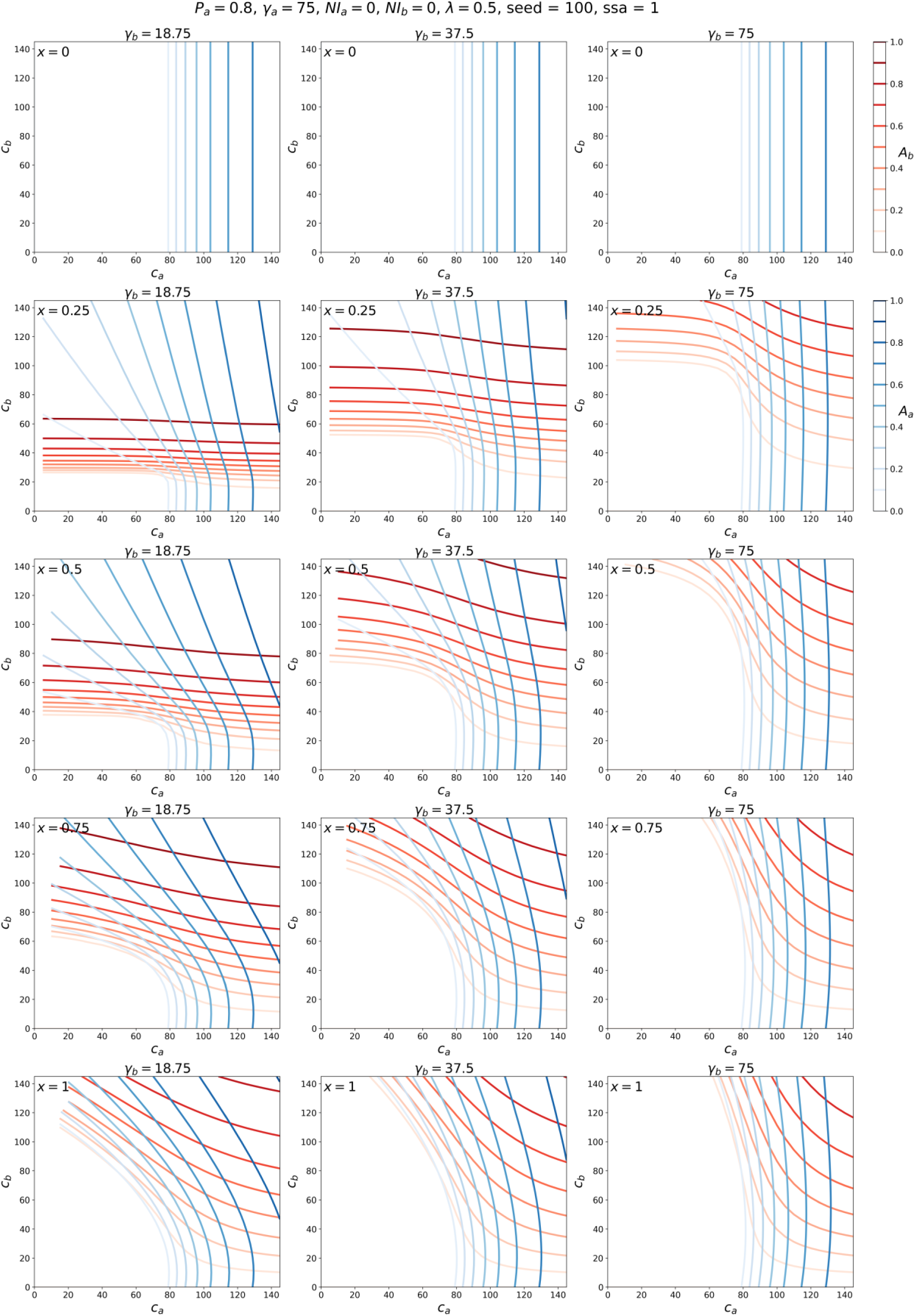
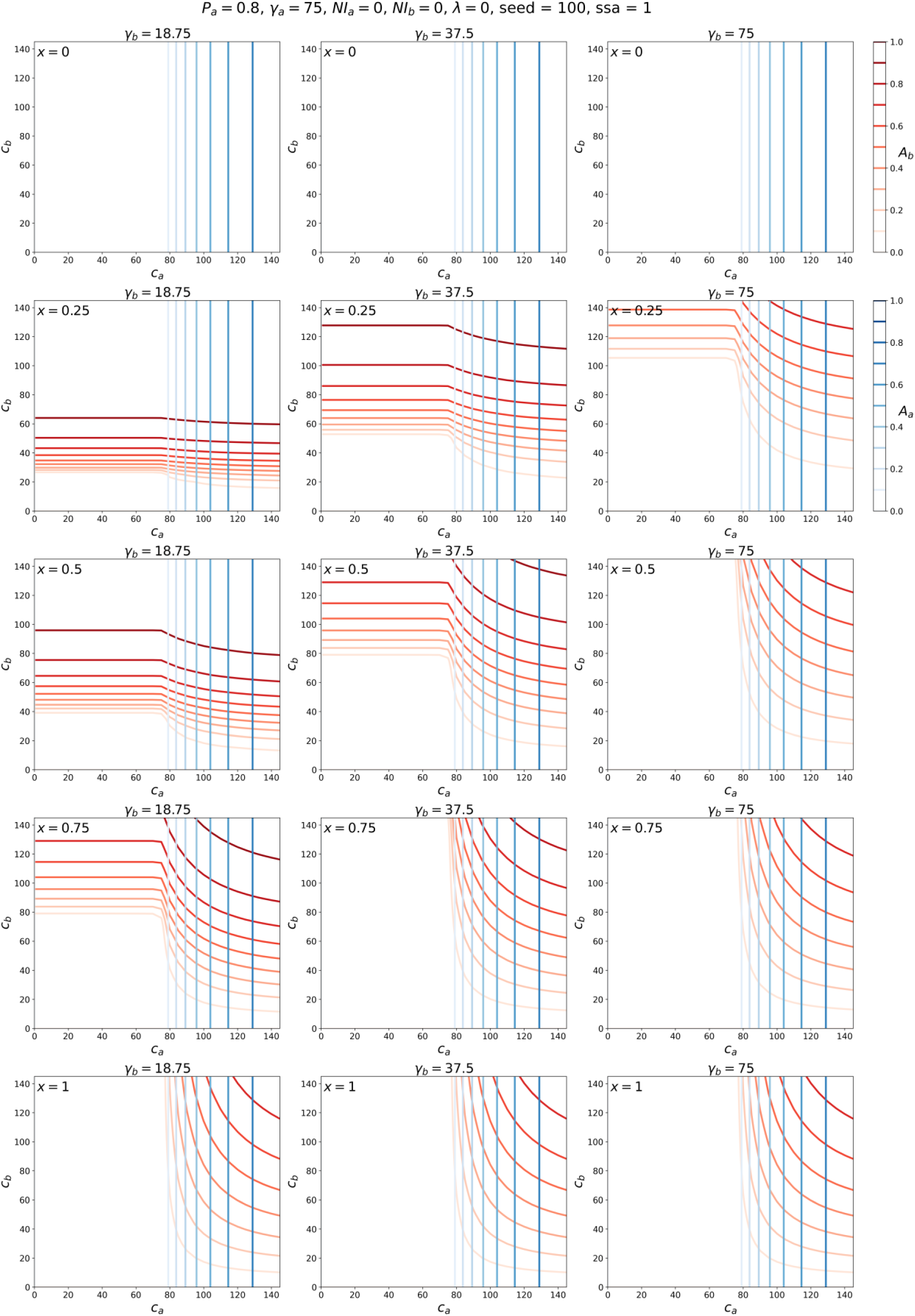

#### C.3 : Attack-rate contour maps for different values of λ, for P_a_ = 0.6, seed = 100, and ssa = 1

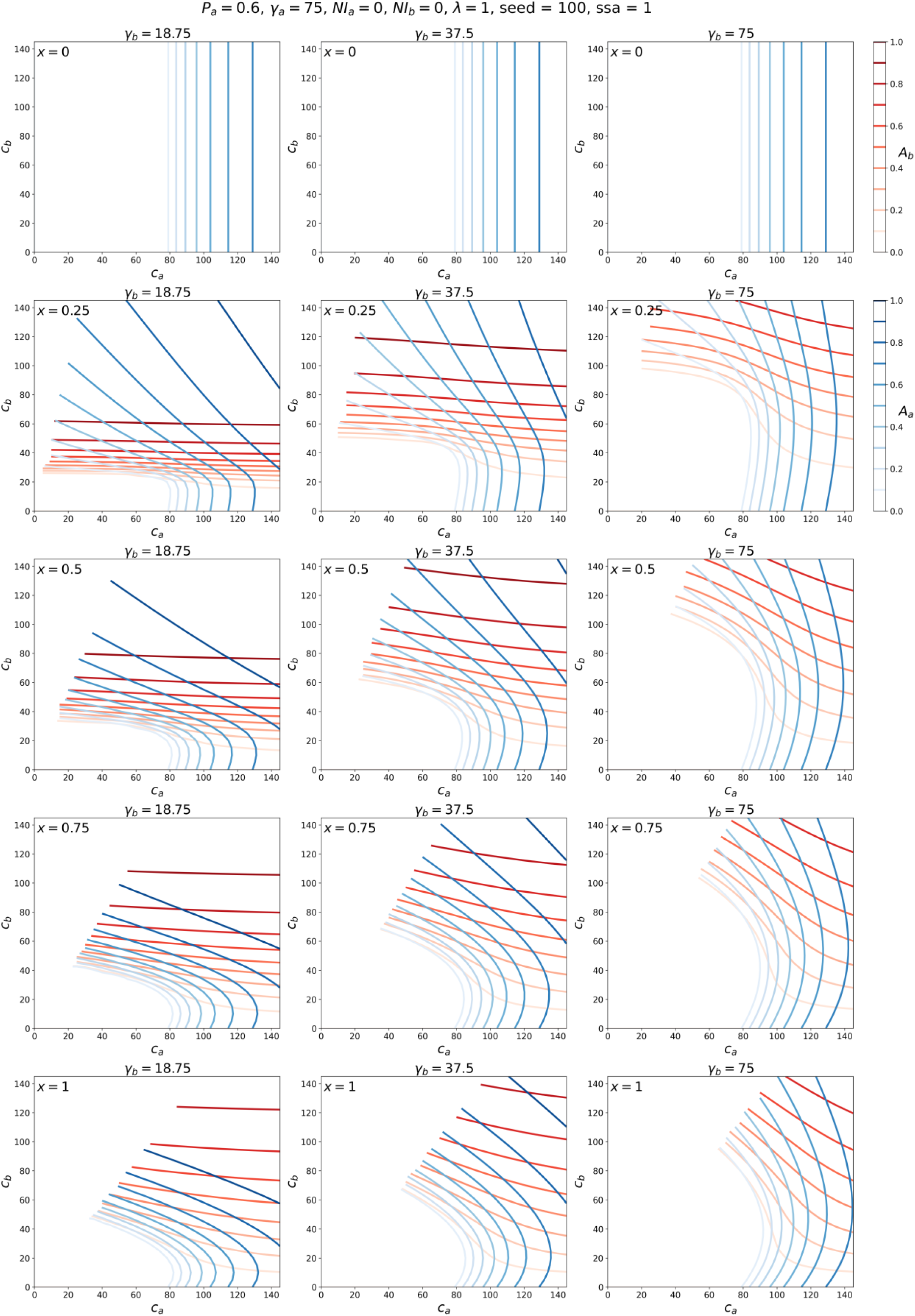
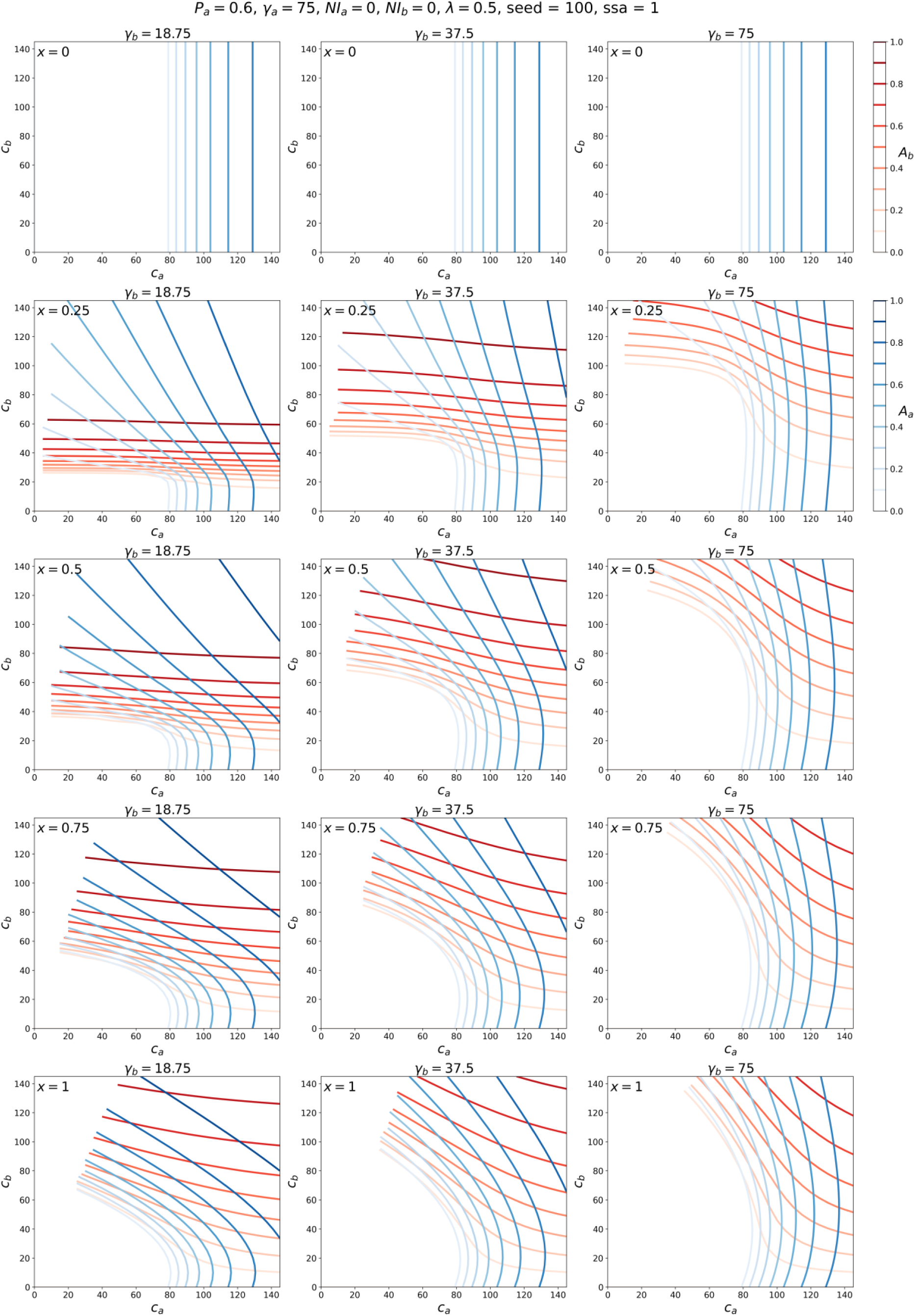
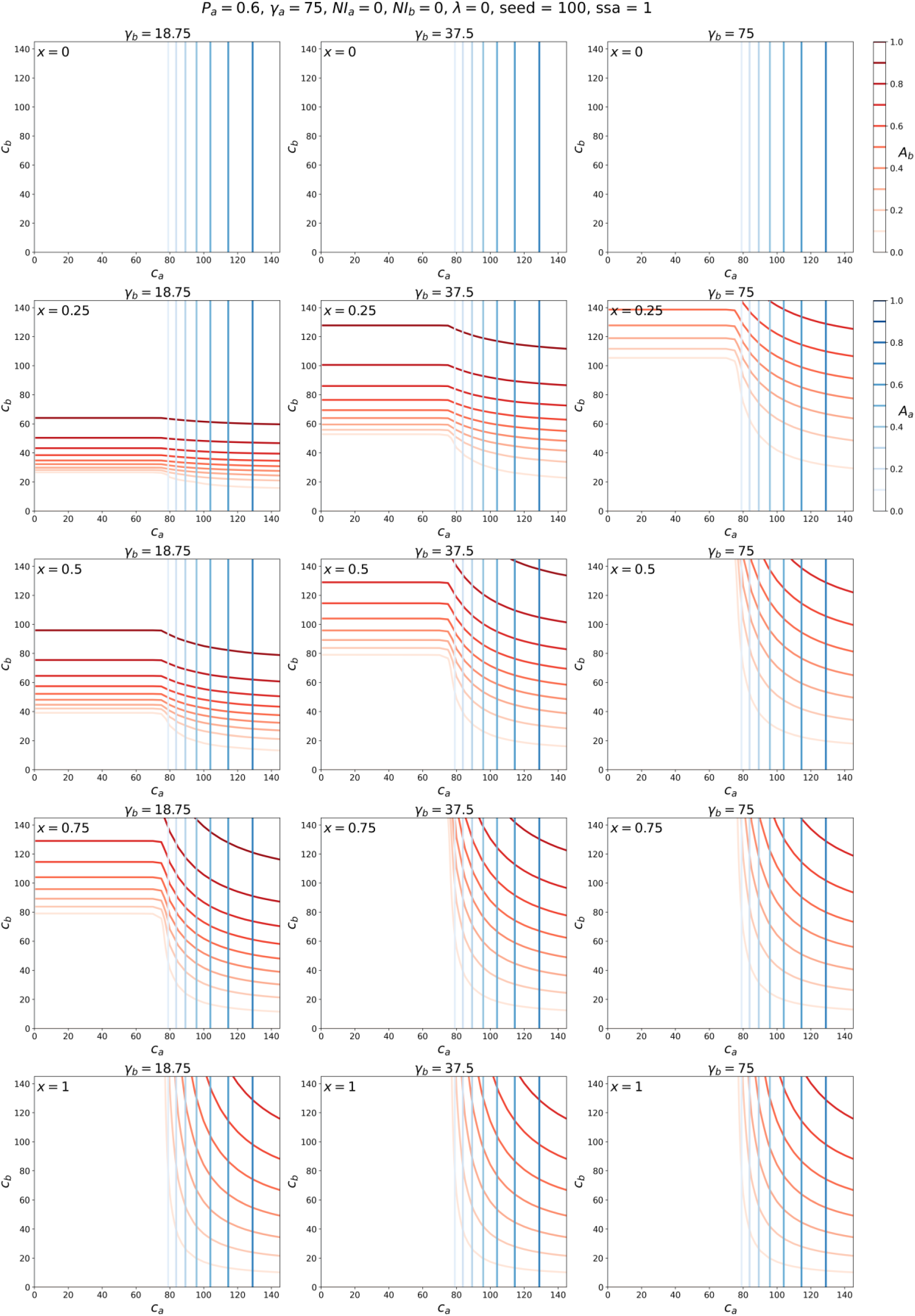

### Appendix D: Varying seed magnitude

This Appendix contains attack-rate contour maps for the parameters used in the main text figures, but for different magnitudes of the initial seed number of infected individuals (parameter “seed”). As can be seen, changing the seed magnitude does not change the attack-rate results.

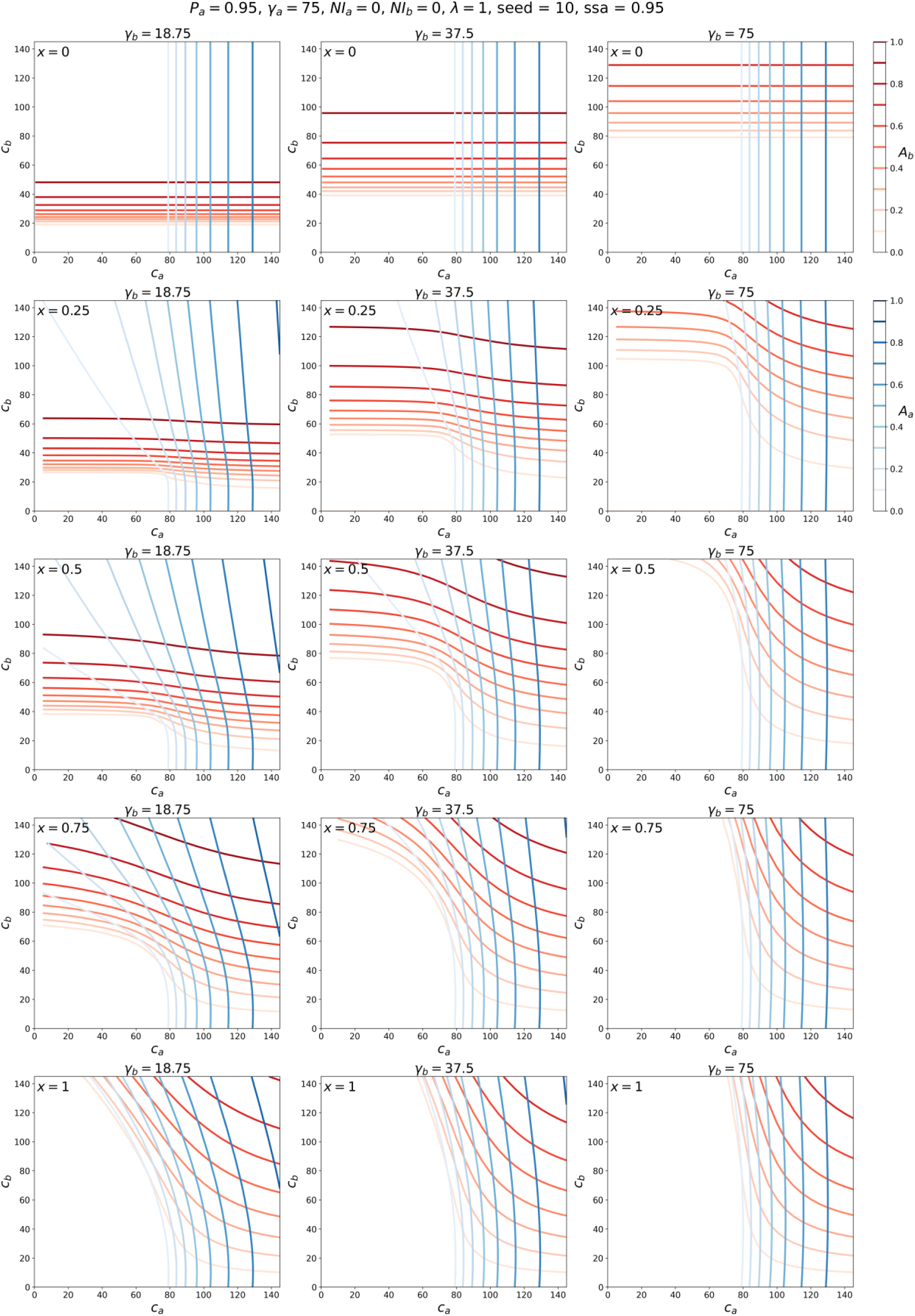
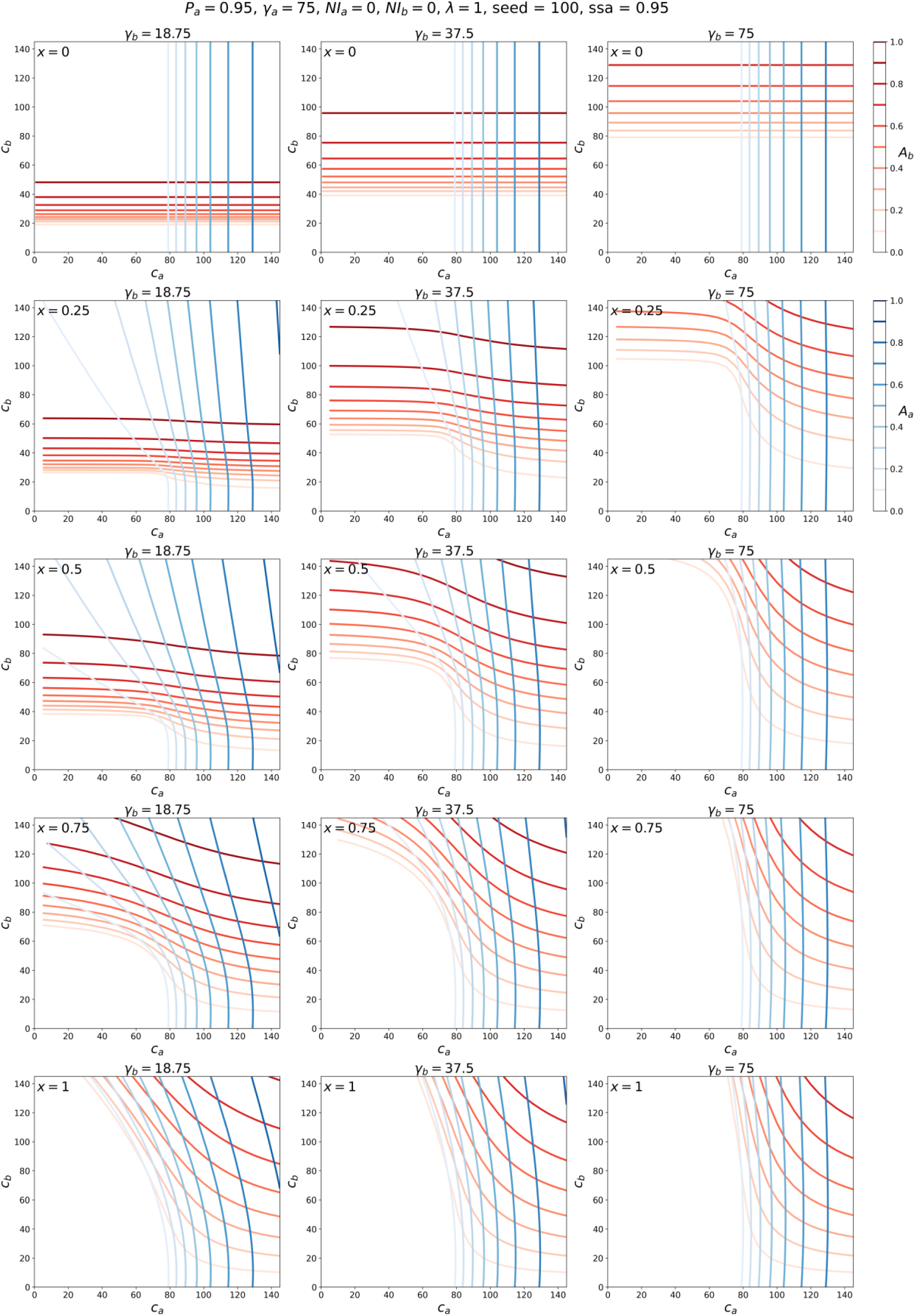
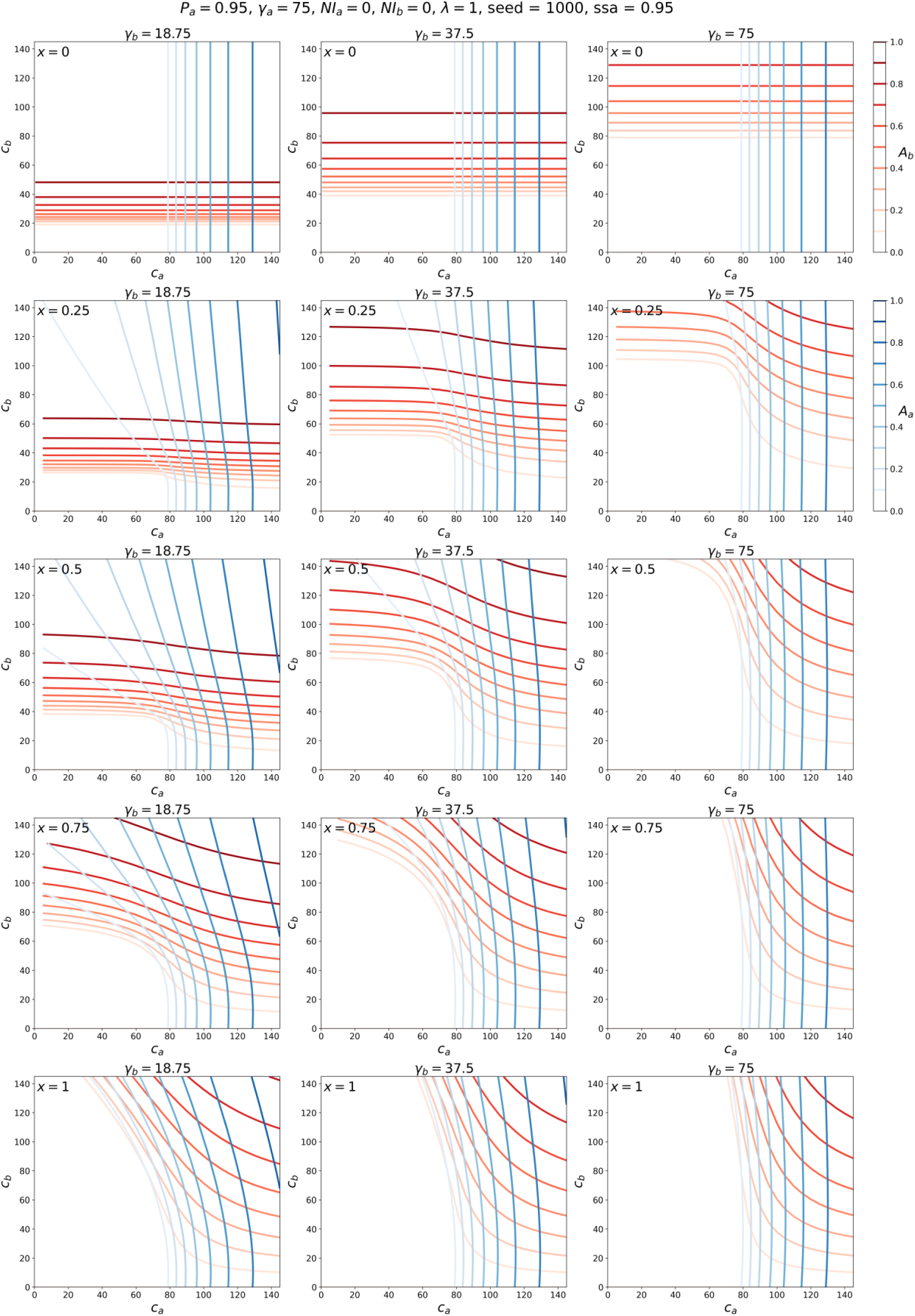
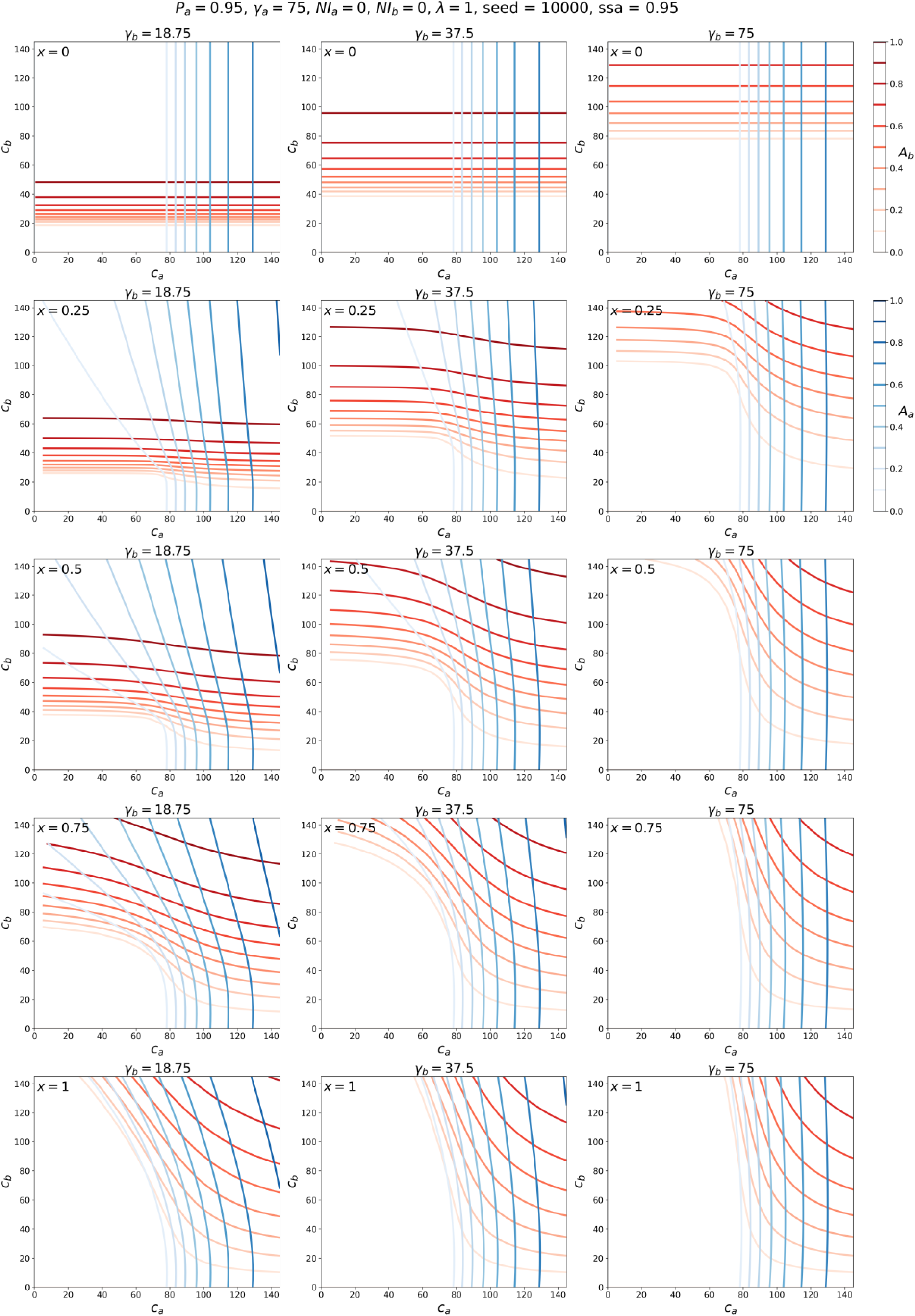

